# Genetic Variants in Recurrent Euploid Pregnancy Loss

**DOI:** 10.1101/2025.10.01.25335660

**Authors:** Mona Aminbeidokhti, Michelle Halstead, Marta Rodriquez-Escriba, Christina G. Tise, Jonathan A. Bernstein, Hakan Cakmak, Elizabeth Pollard, Linda C. Giudice, Katrina Merrion, Gary M. Shaw, Alison Edelman, Maureen Baldwin, David K. Stevenson, Mary Stephenson, Michael P. Snyder, Marina Sirota, Ruth B. Lathi, Svetlana A. Yatsenko, Aleksandar Rajkovic

**Author notes:** Correspondence to either: Aleksandar Rajkovic, MD, PhD, Professor, Departments of Pathology, Obstetrics, Gynecology and Reproductive Sciences, University of California San Francisco. 513 Parnassus Ave HSW 518, San Francisco, CA, 94143, USA, Svetlana Yatsenko, MD, Professor, Department of Pathology, Stanford University, School of Medicine, 3375 Hillview Ave., R2810, Palo Alto, CA, 94304, USA. Drs. RB Lathi, SA Yatsenko, and A Rajkovic contributed equally to this article.

## Abstract

Recurrent pregnancy loss (RPL) affects ∼5% of women, yet the genetic basis of euploid losses remains unclear. We performed genome sequencing in 118 families with unexplained euploid RPL, most without fetal anomalies. We identified genomic variants in 30 families (25.4%) across 28 genes. Thirteen genes were previously linked to perinatal lethality, while fifteen were novel. Inherited variants accounted for 83.3% (25/30) of families, including two due to parental germline mosaicism, seven heterozygous variants transmitted from affected or asymptomatic parents, five cases with hemizygous variants, and 11 with biallelic variants. Four additional families (3.4%) had biparental ultra-rare variants in genes not yet associated with any human disease but with plausible roles in prenatal lethality. Transcriptomic analyses implicated roles in hematopoiesis, cardiovascular development, inflammation, and fluid homeostasis. We identified monogenic basis in a quarter of unexplained euploid RPL cases and expanded our understanding of early human lethality, recurrence risk, and inheritance patterns.

---

Miscarriage is the most common adverse pregnancy outcome, occurring in up to 30% of clinically recognized pregnancies.^1,2^ Recurrent pregnancy loss (RPL), defined as two or more spontaneous losses before 20 weeks’ gestation, affects ∼5% of women of reproductive age, with three or more perinatal losses affecting ∼1%.^3^ RPL is associated with an increased incidence of infertility, intrauterine growth delay, preterm birth, preeclampsia, stillbirth, and neonatal death, suggestive of a shared etiology.^4,5^ Beyond its reproductive toll, RPL carries long-term maternal health risks, including cardiovascular disease and psychological complications such as anxiety, depression, and post-traumatic stress.^6–8^

RPL is believed to be multifactorial, with diverse contributors including parental chromosomal abnormalities, maternal age, uterine anomalies, endocrine or autoimmune disorders, thrombophilia, and environmental factors.^9–11^ Sporadic miscarriage in the first trimester is often due to *de novo* aneuploidy or polyploidy, present in ∼50-70% of products of conception (POC),^12^ while the remaining losses have normal karyotype. In contrast to sporadic pregnancy loss, ∼70% of RPL are chromosomally normal (euploid) and only 4–8% of RPL are attributable to inherited structural chromosomal rearrangements.^13^ Large scale genomic repositories, such as UK biobank and “All of Us”, do not have genomic information on POC.^14,15^ Most RPL cases remain unexplained. Recent next generation sequencing studies in selected families with pregnancy loss and stillbirths point to Mendelian disorders as significant contributors of RPL.^16–21^ Using a bioinformatic approach, we recently identified 138 candidate genes, in which biallelic deleterious variants may potentially lead to prenatal and perinatal loss.^22^ Moreover, it is further estimated that ∼3,400 genes may be essential for human development based on murine and cellular lethality data, far exceeding the ∼600 currently linked to human perinatal lethality,^22,23^ indicating a substantial number of undiscovered disease genes. We hypothesized that many unexplained RPL cases result from variants in genes essential for early development, including genes not previously linked to human disease. In this study, we performed genome sequencing to determine the spectrum of variants in 118 families with unexplained, euploid RPL.

## Results

### Study cohort

Between 2022–2024, 176 families with ≥2 pregnancy losses with no identified cause were enrolled in the study. Exclusion criteria comprised known etiologies such as uterine anomalies, cervical insufficiency, uncontrolled endocrine/autoimmune disease, or fetal infection, and miscarriages with genomic imbalances due to parental chromosome rearrangements. Chorionic villi, amniotic fluid, and fetal organ tissues from the most recent and prior (when available) miscarriages, as well as samples from adverse pregnancy outcomes such as stillbirth and neonatal deaths were collected for genetic analysis. All POC samples underwent karyotype or chromosomal microarray analysis for clinical or research reasons. Short tandem repeat (STR) genotyping confirmed fetal–parental relatedness and assessed maternal cell contamination (MCC). Samples with >20% MCC, aneuploidy, polyploidy, or poor DNA quality were excluded. Ultimately, 118 RPL families had at least one euploid sample from a nonviable conception and parental sample(s) suitable for sequencing (Figure 1A). Clinical data were collected via chart review and questionnaires (Table S1, Figure S1, Figure S2).

**Figure 1.**
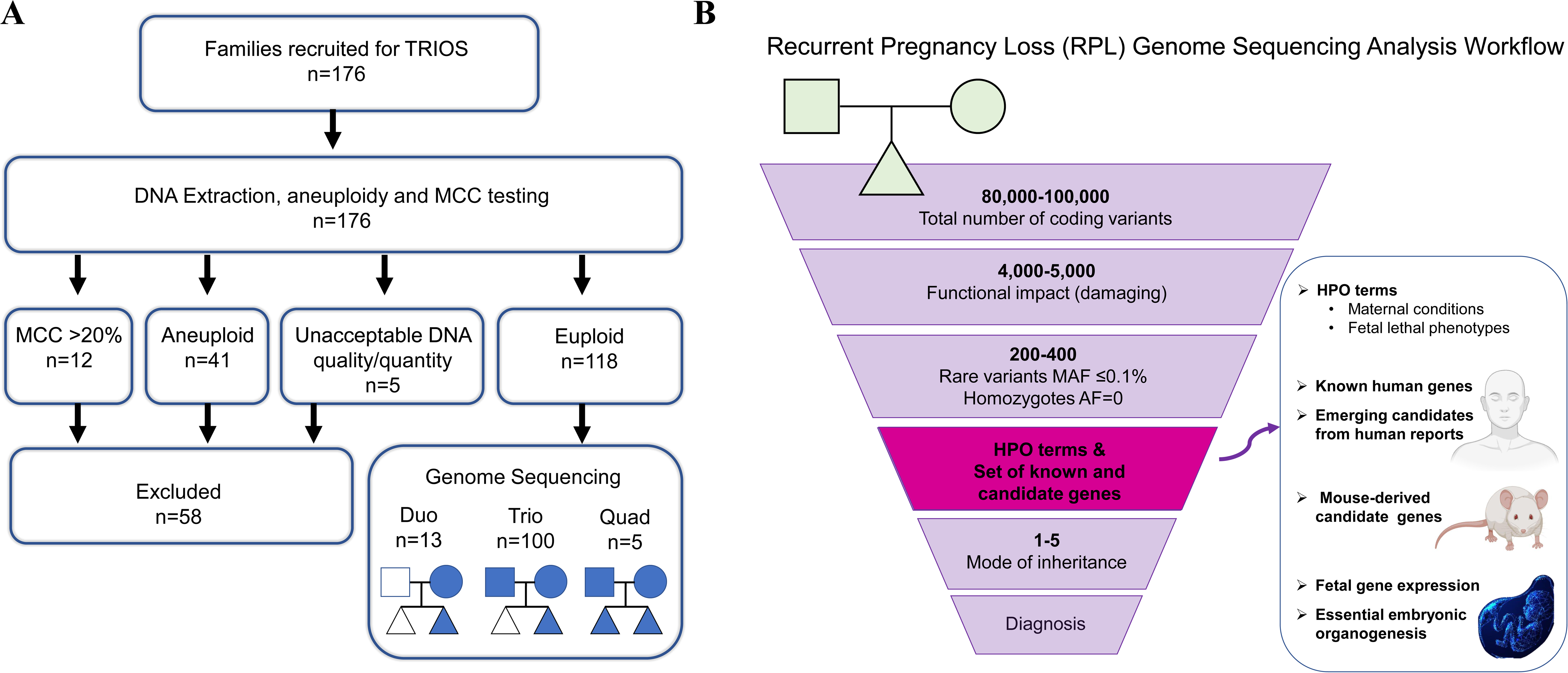
Study Enrollment and Variant Interpretation Workflow. **Panel A** shows the flow of participant recruitment and sample selection. After exclusion of samples with chromosomal abnormalities, maternal cell contamination greater than 20%, or insufficient DNA quality/quantity, 118 families with euploid pregnancy losses proceeded to genome sequencing, including duos, trios, and quads. **Panel B** shows the variant interpretation pipeline, incorporating filtering based on the variant’s functional impact, allele frequency, inheritance pattern, and clinical phenotype. Prioritization incorporated Human Phenotype Ontology (HPO) terms, known and emerging human lethal genes, mouse-derived candidate genes, and developmental transcriptomic data.

### Discovery of genes associated with pregnancy loss

Custom pipelines were used for variant calling and interpretation (Figure 1B). Maternal genomes were screened for variants in RPL-associated genes (Table S2) using Human Phenotype Ontology (HPO)-based terms (Table S3). Fetal variants were prioritized via family-based analysis, integrating: embryonic/perinatal lethality HPO terms (Table S4), lists of 624 known lethal genes, pathogenic variants in which cause fetal demise and neonatal death; 312 genes with emerging evidence for embryonic and fetal lethality ((rpldb.org/intolerome/), and 2,871 mouse-derived candidate genes linked to prenatal lethality in mouse models (Table S5). Strong variants in other genes (with lethality evidence/potential) that are not included in the above categories were considered as findings in novel RPL-plausible genes. Strong candidate variants were defined as those that showed: gene-disease links to prenatal lethality, key developmental roles, fetal expression, constraint metrics, absence from control databases (gnomAD v4.1.0), and consistent inheritance patterns (e.g., *de novo*, recessive, X-linked). Further details are provided in the Supplementary Methods.

### Maternal genes and predisposition to RPL

We first analyzed maternal genomes for variants in 14 genes previously known to be associated with RPL due to thrombophilia, endometrial dysfunction, or hormonal dysregulation (Table S2). Among the 118 women in our study, no P/LP variants were detected in the *AMN, ANXA5*, *F2*, *F5*, *NOS3,* or *SYCP3* gene, which have been shown to increase susceptibility to RPL. Additionally, we did not identify any P/LP variants in *BUB1, BUB1B, KASH5, KHDC3L, MEI1, NLRP7, REC114*, and *TOP6BL* genes, which are implicated in conditions that cause rare syndromic forms of RPL such as recurrent hydatidiform mole, oocyte/zygote/embryo maturation arrest, and mosaic variegated aneuploidy syndrome.

### Genetic Variants in Miscarriage Tissues

A total of 123 samples from 118 RPL families, which included 79 cases of embryonic loss (≤10 weeks’ gestation), 37 cases of fetal demise (11–20 weeks’ gestation), and 6 stillbirths (21-28 weeks’ gestation) and 1 neonatal death underwent genome sequencing and analysis. Most losses (97/123, 78.8%) occurred during the first trimester (Figure 2A), characterized by rapid growth and the formation of major organ systems (Figure 2B). Notably, early processes such as primitive hematopoiesis, the development of the heart, vasculature, and lymphatic system are critical for embryo survival. In our cohort, 95% of cases showed no detectable structural abnormalities by prenatal ultrasonography (Figure 2C). Due to early gestational ages and the negative ultrasound findings, variant interpretation relied on the multifaceted pipeline (Figure 1B) rather than a traditional phenotype-driven approach.

**Figure 2.**
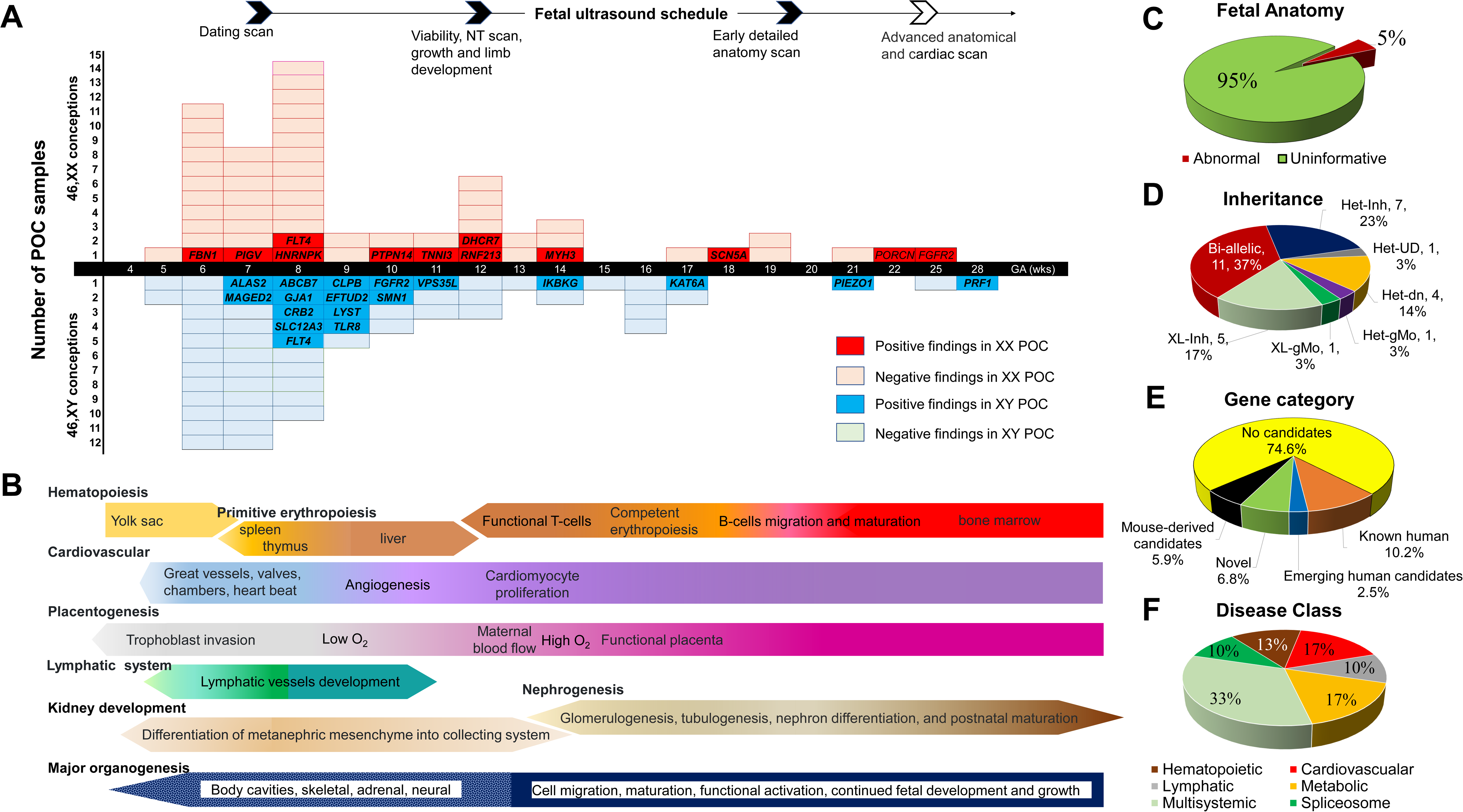
Clinical and Genomic Findings in Recurrent Pregnancy Loss. **Panel A**: Distribution of the miscarriage samples in the study by gestational age (GA) and fetal sex. The y-axis indicates the number of POC samples. Conceptions with 46,XX and 46,XY chromosome complements designated in pink and light blue blocks, respectively. The top of the panel indicates typical timing of prenatal ultrasound evaluations (dating scan in the first trimester, nuchal translucency (NT) scan around 12 weeks, anatomy scans later). Most losses in our series occurred before detailed anatomy scans. Genes identified in positive 46,XX and 46,XY conceptions are shown in the red and dark blue blocks, respectively. **Panel B**: Schematic timeline of human embryonic development and organogenesis. Key early developmental processes are highlighted, including primitive hematopoiesis (yolk sac through fetal liver), cardiovascular development (heart and great vessels, onset of heartbeat), placental development (trophoblast invasion and maternal blood flow), lymphatic system formation, nephrogenesis, and other major organogenesis milestones. **Panel C**: Outcome of prenatal ultrasounds in the RPL cases prior to miscarriage. In 95% of cases, prenatal imaging showed no abnormal findings “uninformative”, whereas ∼5% had abnormal findings. **Panel D**: Inheritance patterns of the likely causative variants. Het: Heterozygous, Inh: inherited, dn: *de novo*, UD: undetermined, gMo: germline mosaicism, XL: X-linked. **Panel E**: Categories of genes identified in positive cases (n=30). Approximately half of the cases had variants in genes previously associated with perinatal lethality (“Known/Emerging” human lethal genes), while the others had variants in candidate lethal genes derived from mouse models or entirely novel genes not previously linked to lethality. **Panel F**: Classification of the disorders identified in the RPL cases by major organ system or disease category.

We identified 39 genomic variants in 28 genes, providing a potential genetic explanation for pregnancy loss in 30 (30/118, 25.4%) families (Table 1). Additional details about these RPL-associated variants and genes are provided in the Supplementary Appendix: Table S6 (variant details), Table S7 (gene categories), Supplementary Results, and Figure S3. Among the 39 variants, 29 were classified as pathogenic (P) or likely pathogenic (LP), and 10 were classified as variants of uncertain significance (VUS), using the ACMG (American College of Medical Genetics and Genomics) criteria.^24^ Each gene was evaluated for lethality evidence from multiple sources, including published human studies of pregnancy loss or perinatal lethal disease, phenotype data from knockout mouse models, the gene’s involvement in essential developmental pathways, and expression in relevant embryonic or fetal tissues (Table 2).

**Table 1.**
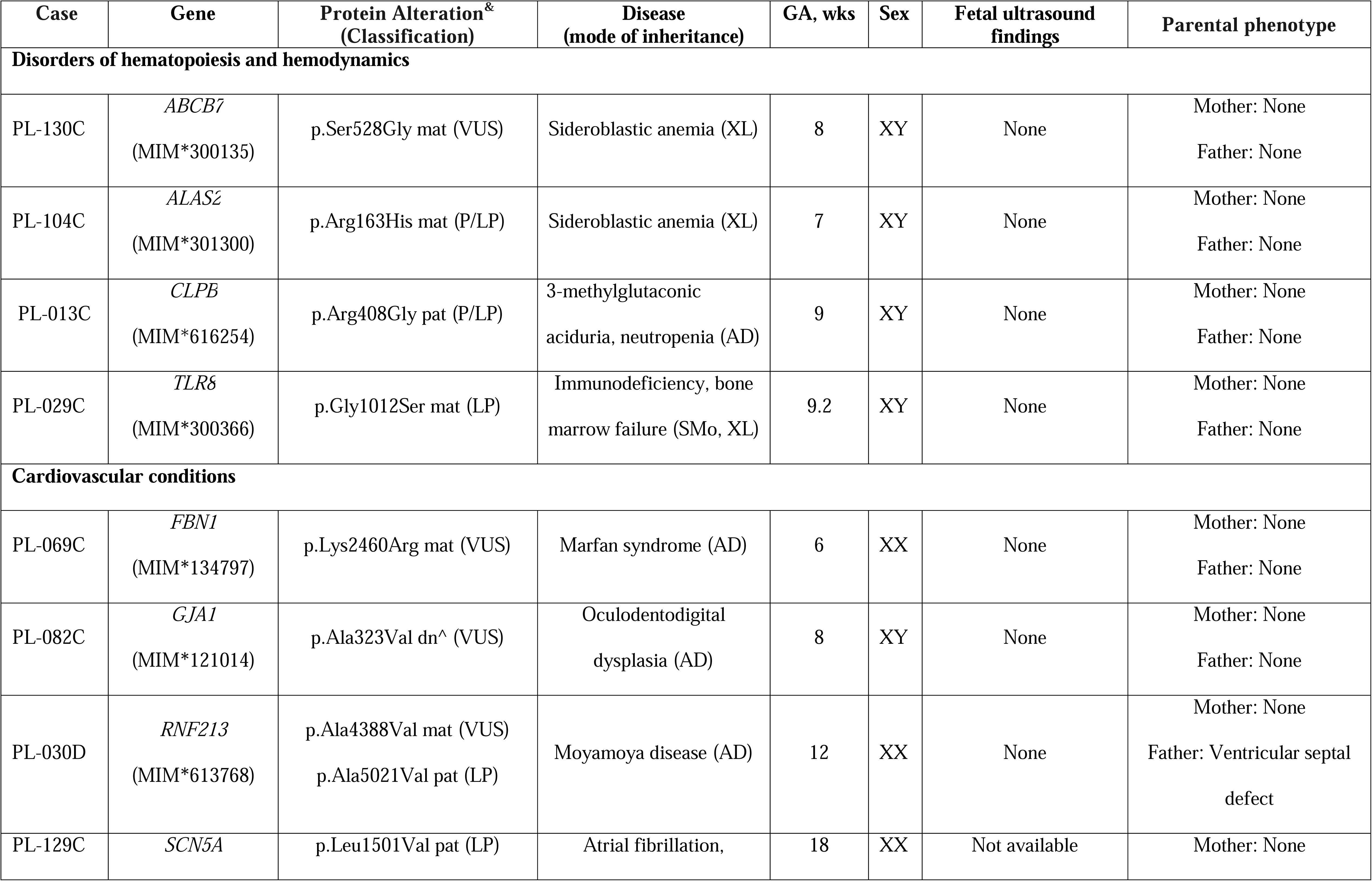
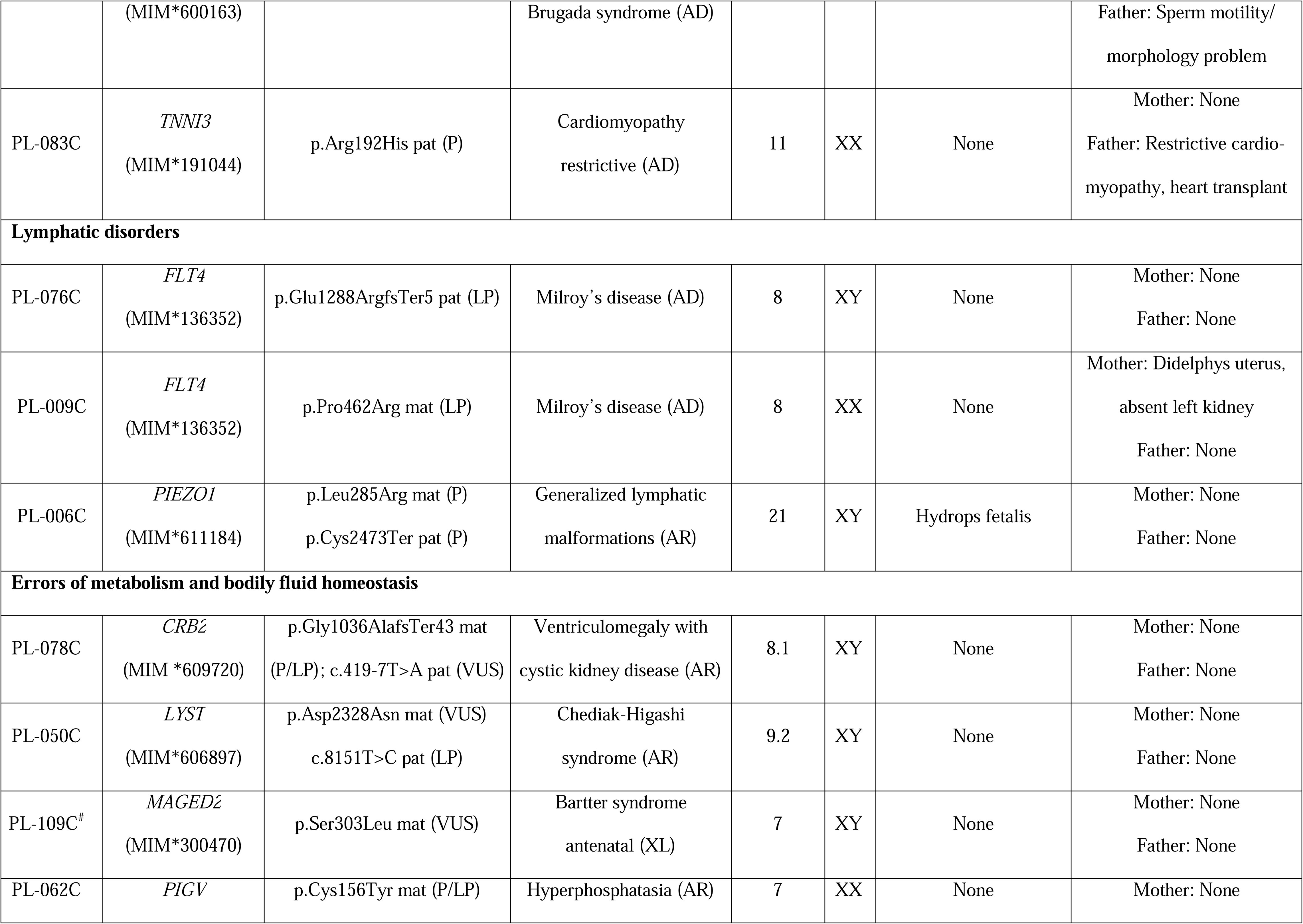
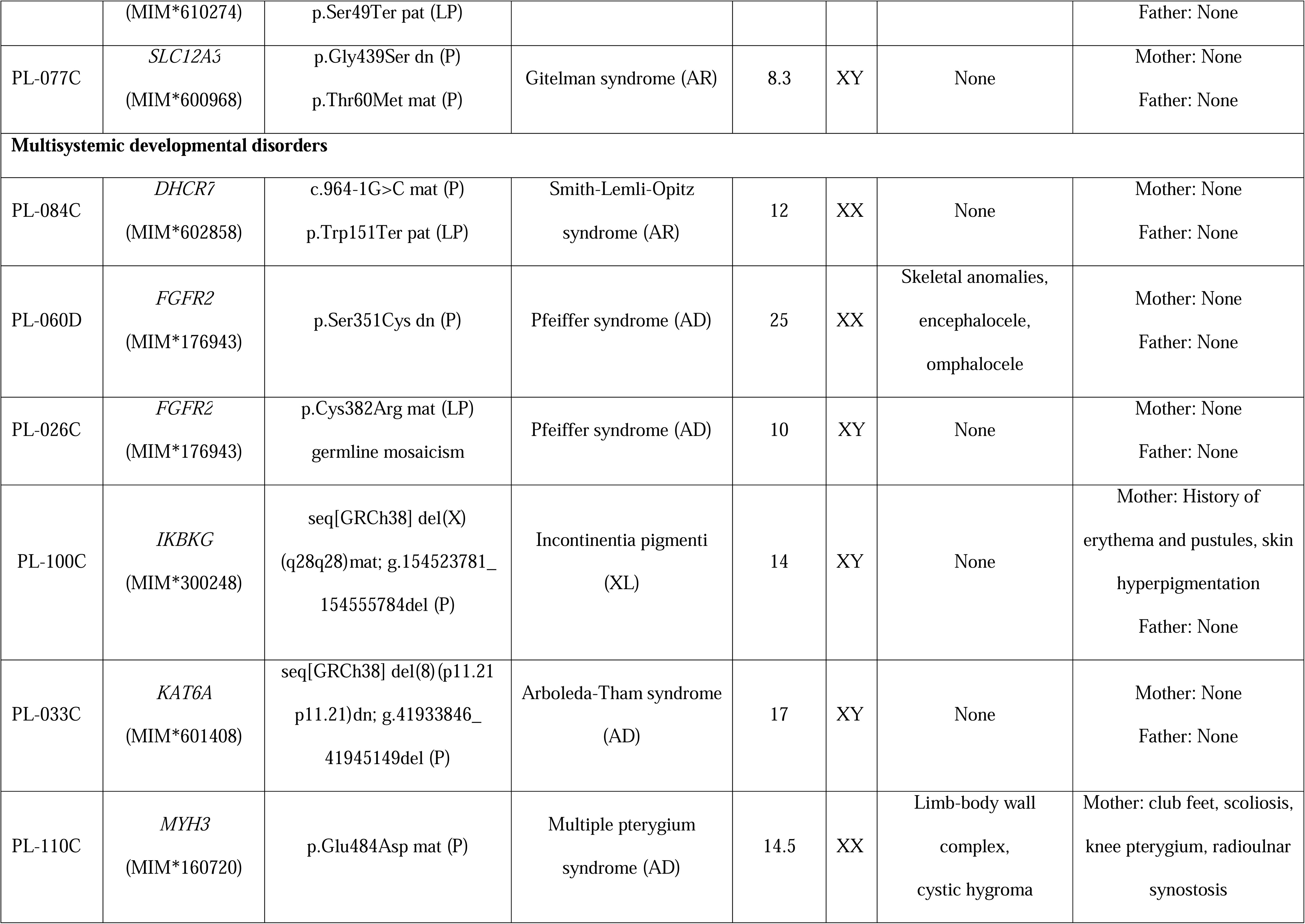
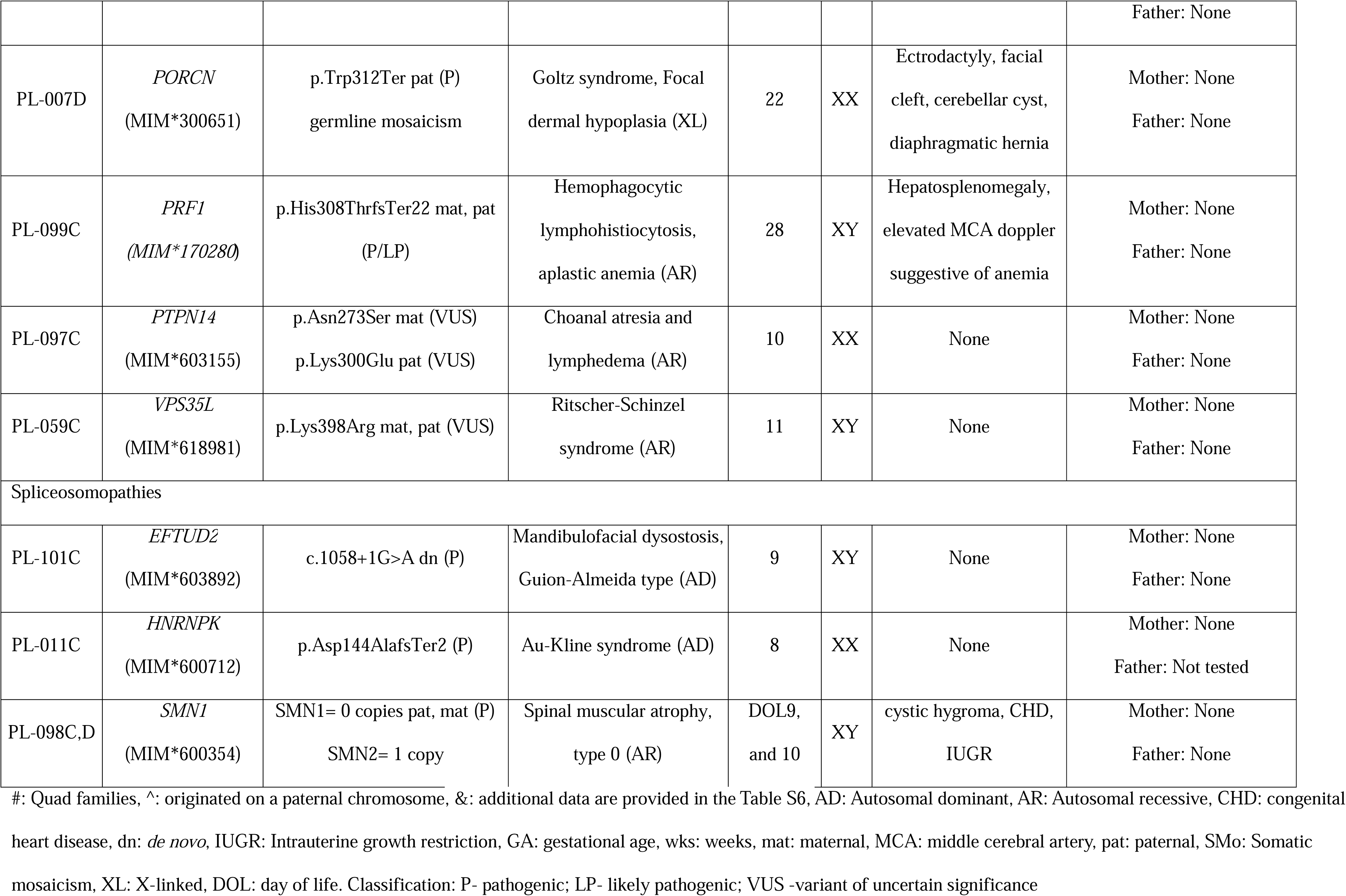
Findings in pregnancy loss samples.

**Table 2.** Genetic contributors to pregnancy loss.

| **Gene** | **Biological function, pathways** | **Expression in fetal tissues/**<br>**human essential organogenesis** | **Human phenotype, evidence for lethality** | **Mouse phenotype,**<br>**evidence in lethality (MGI & IMPC)** |
| --- | --- | --- | --- | --- |
| *ABCB7* | ABC transporter,<br>mitochondrial iron-sulfur<br>cluster biogenesis | Placenta, liver, erythroblasts,<br>thymus/ Intracellular iron ion<br>homeostasis in cardiomyocytes | Anemia, neurodevelopmental problems, brain<br>anomalies | Hemizygous male and heterozygous female<br>mice display prenatal lethality |
| *ALAS2* | Heme synthesis defects and<br>porphyrias | Liver, placenta, spleen, hemato-<br>poietic stem cells/Hematopoiesis | Sideroblastic anemia with variable onset from in<br>utero to adulthood | Embryonic lethality and severe anemia due<br>to arrest of fetal hematopoiesis |
| *CLPB* | DNA replication, protein<br>degradation, chromatin<br>remodeling, granulocyte<br>differentiation | Erythroblasts, thymus, intestine,<br>microglia/ Regulation of<br>hematopoiesis | Severe congenital neutropenia, myeloid matura-<br>tion arrest, polyhydramnios, fetal contractures,<br>IUGR, ovarian insufficiency, infertility and<br>frequent miscarriages, incomplete penetrance. | No lethality in KO mice |
| *CRB2* | Notch signaling, regulation<br>of epithelial to mesenchy-<br>mal transition, establish-<br>ment of cell polarity | Cerebellum, retina, kidney,<br>mesothelial cells, somitogenesis,<br>mesoderm formation/<br>Circulatory system development | Glomerulosclerosis, ventriculomegaly, cystic<br>kidney, hydrocephalus, neonatal demise | Prewaning lethality, severe gastrulation<br>defects, impaired organogenesis, embryonic<br>death by E12.5 |
| *DHCR7* | Cholesterol homeostasis,<br>multicellular organismal<br>processes, Hippo, WNT,<br>and SHH pathways | Widely expressed. Abundant in<br>adrenal glands, lever, brain,<br>/Differentiation of embryonic<br>cells | Impaired brain, limb, and genital development,<br>prenatal and neonatal demise, disease<br>prevalence is lower than expected from<br>observed carrier frequency | Homozygous KO mice show preweaning<br>lethality, abnormal cholesterol homeostasis<br>with reduced tissue cholesterol levels and<br>total sterol levels. |
| *EFTUD2* | Elongation factor, Ribonucleoprotein, mRNA splicing via spliceosome | Widely expressed. Abundant in kidney, spleen, and thymus | Microcephaly, choanal atresia, germline mosaicism for loss-of-function variants reported in recurrent pregnancy loss | Prewaning lethality, embryo arrest before implantation, transient developmental delay between E8.5-E9.5 in heterozygotes. |
| *FBN1* | Integrin Pathway, ERK signaling, calcium ion binding, extracellular matrix structural constituent | Placenta (extra-villous trophoblasts), muscle, perivascular macrophages, nephron progenitor cells | Intrauterine growth restriction, neonatal mortality, cardiovascular involvement, obstetrics complications, aortic dissection, hemorrhage. Women with Marfan syndrome have a higher rate of early spontaneous abortion. | Prewaning lethality with incomplete penetrance (embryonic death to 4 months). Abnormalities include vascular defects, excess bone growth, connective tissue hyperplasia, and lung emphysema. |
| *FGFR2* | RAS, MAPK1/ERK2, MAPK3/ERK1, MAP kinase and AKT1 signaling pathways | Widely expressed. Abundant in lung, kidney. /Regulation of cell proliferation, differentiation, embryonic patterning, trophoblast function, osteogenesis. | Ectrodactyly, congenital cystic adenomatoid malformation of the lung, abnormal facial shape, syndactyly, oligodactyly, pulmonary hypoplasia, abnormal lung lobation, prenatal and neonatal demise | Lethality ranges from embryonic stage to 20 days after birth due to defects in multiple organs and tissues. |
| *FLT4* | MAPK1/ERK2, MAPK3 /ERK1, and AKT1 signaling pathways | Placenta, vascular endothelial cells, spleen / development of the vascular network and the cardiovascular system | Lymphedema (Milroy disease), nonimmune hydrops fetalis, Tetralogy of Fallot, CHD | Growth retardation, vascular anomalies, anemia, death from cardiovascular failure by E9.5. Heterozygotes show abdominal chylous ascites, abnormal lymphatic vessels, and lymphedema. |
| *GJA1* | Gap junction trafficking, mitotic spindle orientation, signal transduction, cell-cell | Adrenal gland, cerebellum, heart/ Osteoblast differentiation, neuron migration, heart looping | Arrhythmogenic right ventricular cardiomyopathy, hypoplastic left heart syndrome, atrioventricular septal defect, sudden | Homozygotes have heart and outflow tract malformations, eye lens anomalies, and male germ cell deficiency. Neonatal lethality. |
|  | signaling | and conduction development | infant death, microphthalmia, syndactyly |  |
| *HNRNPK* | mRNA processing mRNA splicing, via spliceosome, signal transduction | Fetal brain, heart, lung | Hypotonia, global developmental delay, congenital heart defects, genitourinary, skeletal abnormalities, neonatal death | Embryonic lethality of homozygous KO mice |
| *IKBKG* | AKT Signaling, NF-κB Signaling, Toll Like Receptor 7/8 (TLR7/8) Cascade | Heart, brain, placenta, kidney, liver, lung, skeletal muscle/ Formation of ectodermal tissues, global transcriptional regulation | Incontinentia pigmenti, hypohidrotic ectodermal dysplasia, immunodeficiency, male lethality, first trimester miscarriage | Embryonic lethality by E13.5 in males, apoptotic liver damage. Heterozygous females show patchy skin lesions with granulocyte infiltration, growth retardation, and shortened lifespan. |
| *KAT6A* | Gene expression (transcription) and chromatin organization | Kidney, thymus, bipolar cells, eye/ regulation of hemopoiesis, chromosome organization, cellular senescence | Microcephaly, cardiac anomalies, intellectual disabilities, gastrointestinal complications, moderate to severe neutropenia | Perinatal lethality, cyanosis, depletion of hematopoietic progenitor cells, severely impaired spleen and thymus development. Heterozygotes display fertility issues. |
| *LYST* | Intracellular protein trafficking in endosomes, phagocytosis | Abundantly expressed in fetal thymus, peripheral blood leukocytes, bone marrow, cerebellum. | Severe immunologic deficiency, a bleeding tendency, neurologic abnormalities, abnormal intracellular transport to and from the lysosome. Most patients die at an early age | Lysosomal and immune cell dysfunction, increased susceptibility to infections, decreased cytotoxic activity. Prewaning lethality with incomplete penetrance. |
| *MAGED2* | Response to elevated platelet cytosolic $\text{Ca}^{2+}$, regulates the function of the sodium chloride cotransporters | Embryonic kidney, endocardial cells, skeletal muscle, lung, placenta/ renal sodium ion absorption, control of fetal blood | Severe polyhydramnios, preterm delivery, stillbirth, nephrocalcinosis, excessive loss of sodium and potassium | Lethality in KO mice is not reported. |
|  | SLC12A1 and SLC12A3 | pressure and fluid balance |  |  |
| *MYH3* | Rho GTPases and PAK pathways, cytoskeleton regulation and remodeling | Skeletal muscle, eye, perivascular macrophages/ skeletal development | Multiple pterygium syndrome, skeletal anomalies, growth delay, nonimmune hydrops fetalis, prenatal and neonatal demise | Embryonic lethality, incomplete penetrance. Defective differentiation and development of myogenic precursor cells and myoblasts. |
| *PIEZO1* | Ion transmembrane transport, regulation of membrane potential | Lymphatic endothelial cells, perivascular macrophages, liver, erythroblast, spleen / heart failure, fluid accumulation | Lymphedema, nonimmune hydrops fetalis, lymphatic dysplasia, dehydrated hereditary stomatocytosis, hemolytic anemia, prenatal and neonatal demise | Embryonic growth retardation, pericardial effusion, and vascular remodeling defects in the yolk sac and the embryo. Prewaning lethality with complete penetrance. |
| *PIGV* | Synthesis of GPI-anchored proteins, posttranslational modification | Adrenal glands, pancreas, ciliated epithelial cells | Hyperphosphatasia, elevated serum alkaline phosphatase, seizures, hypotonia, facial dysmorphism, and brachytelephalangy | Complex congenital heart disease associated with heterotaxy, thymus hypoplasia, craniofacial and kidney defects |
| *PORCN* | Glycoprotein metabolism, WNT signaling, protein palmitoylation | Hippocampus, cerebellum, kidney, retina, placenta | Skin defects, skeletal abnormalities, ocular anomalies, focal dermal hypoplasia, prenatal and neonatal demise | Dermal atrophy, sternum hypoplasia, cleft palate, tail hypoplasia, absent autopod, and perinatal lethality. |
| *PRF1* | Granzyme A mediated apoptosis pathway, CTL mediated immune response against target cells | Placenta, thymus/ calcium ion binding and wide pore channel activity, apoptotic process, cellular defense response | Generalized edema, hepatosplenomegaly, liver dysfunction, neurologic impairment, pancytopenia, coagulation abnormalities, neonatal demise | Homozygous null mice exhibit increased susceptibility to viral infection and defective cytotoxic T cell cytolysis and NK cell cytolysis. |
| *PTPN14* | Cytokine signaling in immune system and interleukin-1 family | Kidney, placenta/ Regulation of lymph-angiogenesis, cell-cell and cell-matrix adhesion, cell | Choanal atresia, skeletal anomalies, lymphedema | Postnatal growth retardation, decreased body weight, periorbital and limb edema, and lymphatic vessel hyperplasia. Embryonic |
|  | signaling | migration, growth and epithelial-mesenchymal transition |  | lethality prior to organogenesis at E9.5 and preweaning lethality, incomplete penetrance. |
| *RNF213* | Protein ubiquitination, regulation of lipid metabolic process, ATP binding and hydrolysis | Brain, spleen, thymus/<br>Angiogenesis | Vascular occlusion, intracranial hemorrhage, congenital microcephaly, infantile epileptic encephalopathy, coronary artery disease<br>profound developmental delay | Decreased body weight, low insulin and leptin plasma levels. Lethality in KO mice is not reported. |
| *SCN5A* | Sodium ion transmembrane transport, signaling in cardiomyocytes, expressed in human sperm, regulates sperm motility and capacitation | Brain, heart/ Hemodynamic compromise | Atrial fibrillation, ventricular fibrillation, cardiomyopathy, Brugada syndrome, sudden infant death | Homozygous KO mice show prenatal lethality during organogenesis, decreased embryo size, abnormal cardiovascular physiology. Heterozygous mice display abnormal heartbeats and defects in the impulse conduction system. |
| *SLC12A3* | Transport of inorganic cations, anions and amino acids, oligopeptides, disorder of transmembrane transporters | Spleen, thymus, kidney, meta-nephric cells /Transporter and sodium chloride symporter activity, electrolyte homeostasis /Cell volume homeostasis | Hypotension due to intravascular volume depletion, polyuria, renal potassium and magnesium wasting | Homozygous knock-in mice exhibit hypotension, high plasma renin activity, high aldosterone levels, hypokalemia, hypomagnesemia, hypocalciuria, and metabolic alkalosis. |
| *SMN1* | Spliceosomal complex assembly, mRNA processing | Widely expressed. Abundant in brain, kidney, spleen, placenta /mRNA splicing, neurogenesis | Degeneration of the anterior horn cells of the spinal cord, hypotonia, muscle atrophy, cardiac malformations, neonatal death | Homozygotes exhibit cell loss and peri-implantation lethality |
| *TLR8* | Toll Like Receptor 7/8 | Liver, placenta/ Immunity, | Autoimmune hemolytic anemia, | Mice homozygous for a knock-out allele |
|  | (TLR7/8) cascade,<br>canonical NF-kappaB signal<br>transduction | inflammatory response | hyperproduction of inflammatory cytokines,<br>lymphoproliferation, B-cell defects, neutropenia<br>and bone marrow failure. Germline mosaicism<br>in affected males with gain of function variants | exhibit increased anti-nuclear antigen<br>antibodies, altered immunoglobulin levels,<br>decreased marginal zone, B-1a, and B-1b<br>cells, splenomegaly, and glomerulonephritis |
| *TNNI3* | Ion homeostasis, striated<br>muscle contraction, cardiac<br>conduction | Heart, adrenal, perivascular<br>macrophages /Heart<br>development and function | Dilated cardiomyopathy, heart failure, ischemic<br>heart disease, edema, mitral, and tricuspid<br>insufficiency, myocardial fibrosis, sudden death.<br>Variable severity and onset (neonatal-adult). | Ca<sup>2+</sup> dependent control of myofilament<br>activity, reduced germline transmission.<br>Homozygous KO mice die due to acute heart<br>failure around 18-20 days of age. |
| *VPS35L* | Endosomal membrane<br>protein recycling, regulates<br>cell migration and adhesion | Hematopoietic stem cells,<br>placenta, thymus, microglia<br>/Hematopoiesis, cell homeostasis | Cranio-cerebello-cardiac dysplasia, neonatal<br>death | Embryonic lethality between E7.5 and E10.5<br>with complete penetrance. |

MGI: Mouse Genome Informatics (www.informatics.jax.org), IMPC: International Mouse Phenotyping Consortium (www.mousephenotype.org),
CHD: congenital heart disease, E: embryonic day, IUGR: Intrauterine growth restriction, KO: knockout

### Biallelic variants in autosomal genes

Biallelic variants (Figure 2D) were found in POCs from 11 families (37%, 11/30 of all positive cases or 9.3%, 11/118 of total cases). Homozygous and compound heterozygous variants were detected in *CRB2*, *DHCR7*, *LYST*, *PIEZO1*, *PIGV*, *PRF1*, *PTPN14*, *RNF213*, *SLC12A3*, *SMN1*, and *VPS35L* genes, 10 of which are associated with autosomal recessive disorders. In 5 families, both parents carried a P/LP variant in either *DHCR7*, *PIEZO1*, *PIGV*, *PRF1*, or *SMN1* gene. In PL-077C, two pathogenic variants in the *SLC12A3* gene were detected, one of which was maternally inherited, and one occur *de novo* on a paternal allele. In two couples with variants in *CRB2* and *LYST*, one partner carried a P/LP variant while the other had a variant classified as VUS. In one POC, we identified compound heterozygous LP and VUS missense variants in the *RNF213* gene. This gene is associated with autosomal dominant Moyamoya disease in heterozygous carriers, but the biallelic variants have been associated with early-onset severe systemic vasculopathy.^25^ In two families, PL-097 and PL-059, with PTPN14 and VPS35L variants, respectively, both parents were carriers of novel variants not observed in the homozygous state; both variants were classified as VUS (Table 1).

### Heterozygous variants in autosomal genes

In 13 families, heterozygous variants in 11 genes (Figure 2D) were present in 13 POCs (43% of positive cases (13/30) or 11% (13/118) of total cases). Seven inherited variants (six P/LP and one VUS) were observed in the *CLPB*, *FBN1*, *SCN5A*, *TNNI3*, *FLT4* (two cases), and *MYH3* genes, associated with autosomal dominant (AD) conditions with variable expressivity and incomplete penetrance. Only two parents with pathogenic variants in the *TNNI3* and *MYH3* gene, had clinical manifestations consistent with cardiomyopathy and multiple pterygium syndrome, respectively (Table 1). In five remaining families, the parent carrying the variant had little or no clinical manifestations. These findings may imply variable penetrance for certain gene variants. In additional five POC samples, heterozygous variants in *GJA1*, *FGFR2* (two cases)*, KAT6A,* and *EFTUD2* genes appeared to be *de novo*, however in one family with a LP variant in the *FGFR2* gene, germline mosaicism was detected in the maternal blood and urine samples (Supplementary Appendix, Figure S3-8). For one POC (PL-011C) with only mother available for sequencing, a pathogenic heterozygous variant in *HNRNPK* gene was not present in the mother.

### **X-** linked gene variants

Six POCs (20%, 6/30 of positive cases or 5.1%, 6/118 of total cases) had X-linked variants in *ABCB7*, *ALAS2*, *MAGED2*, *IKBKG*, *TLR8*, and *PORCN* genes (Figure 2D), of which four were classified as P/LP variants and two as VUS (Table 1, Table S6). Five hemizygous variants detected in 46,XY POCs were inherited from their heterozygous carrier mothers. Hemizygous variants in *ABCB7*, *ALAS2*, and *MAGED2*, associated with X-linked recessive disorders (sideroblastic anemia and antenatal Bartter syndrome), can manifest with hydrops or anemia in utero in affected males. Hemizygous pathogenic variants in *IKBKG* and *TLR8* are known to cause male-lethal conditions, while heterozygous females and males with germline mosaicism are variably affected. Lastly, in one family, we identified a pathogenic variant in the *PORCN* gene in two 46,XX miscarriages. *PORCN* variants cause an X-linked dominant focal dermal hypoplasia (Goltz syndrome), which is usually male-lethal, while females are variably affected. In this family, the father was found to have ∼3% germline mosaicism (Figure S3-19) upon examination of multiple tissues, explaining the recurrence.

Overall, inherited variants were responsible for the majority of genetic findings in our RPL cohort (83.3%, 25/30 of positive cases or 21.2%, 25/118 of the entire cohort), including 1.7% (2/118) of families with a germline mosaicism.

### Known and novel candidate genes

Fifteen out of the 30 families with positive findings had variants in 13 genes (*CLPB, CRB2, DHCR7, FBN1, FGFR2, IKBKG, MAGED2, PIEZO1, PIGV, PORCN, SMN1, MYH3, FLT4*) that have previously been linked to human fetal demise or neonatal death (known and emerging human lethal genes in the Table S7). Deleterious variants in these genes are known to cause severe congenital syndromes that can manifest prenatally. The remaining 15 families carried genetic variants in genes that, to our knowledge, have not previously been directly linked to pregnancy loss. In seven families, variants were discovered in the “mouse-derived” candidate genes (knockouts of the *EFTUD2, GJA1, KAT6A, LYST, PTPN14, SCN5A, TNNI3* genes showed lethal phenotypes in mouse models, Table S7). In eight families, variants in entirely novel candidate genes (*ABCB7, ALAS2, HNRNPK, PRF1, RNF213, SLC12A3, TLR8, VPS35L* (Figure 2E) were found. Each of these genes is associated with a known severe condition in humans, however this is a first time these genes were linked to pregnancy loss.

In addition to 28 genes described above, we also uncovered strong candidate variants that did not yet meet pathogenicity criteria. In four families (3.3%, 4/118), we identified biallelic deleterious variants in seven genes (*FHL2*, *ADAMTS16*, *NCAPG*, *RIIAD1*, *NCKIPSD*, *SRRD*, and *IRX2*) not linked to any known human phenotype (Table S8). All genes are expressed in embryonic/fetal tissues and the detected variants are absent in a homozygous form in the general population database (gnomAD v4.1.0). These included genes involved in signaling and cell structure, and while plausible contributors to lethality, were conservatively classified as variants of uncertain significance due to limited evidence.

Thus, the genetic findings in a total of 35 genes, 15 known and 20 candidate genes were implicated in pregnancy loss.

### Developmental pathways

The genes identified in our RPL cohort span key developmental pathways and organ systems (Table 1), many of which are critical during the first trimester of pregnancy (Figure 2B). At least 75% of these genes are involved in hematopoiesis, angiogenesis, cardiovascular development, inflammation, and fluid homeostasis (Table 2). Approximately 17% of the genes identified in our cohort associate with cardiovascular disorders, while another 17% associate with metabolic or homeostatic disorders affecting kidneys or endocrine organs (Figure 2F). Hematopoietic disorders accounted for 13% of the genes in our cohort. Three genes (∼10%) are vital for lymphatic development, where disruption can lead to hydrops fetalis. Notably, 43% of cases involved genes underlying complex multisystem disorders and spliceosome components *EFTUD2*, *HNRNPK*, and *SMN1,* illustrating the importance of RNA processing in early embryogenesis. These pathways underscore how embryonic viability relies on genes that govern circulatory function, hematologic balance, placentation, and core cellular processes.

Functional and cluster analysis of 35 detected genes using STRING bioinformatics tools (Figure S4) revealed a statistically significant enrichment in protein-protein interactions (*p* value = 0.0393). Ten genes (*CRB2*, *FBN1*, *FGFR2*, *FHL2*, *FLT4*, *GJA1*, *PTPN14*, *RNF213*, *SCN5A*, and *TNNI3*) participate in circulatory system function and development. Network analysis identified three enriched clusters: Cluster 1 (*IKBKG*, *LYST*, *PRF1*, *TLR8*, and *FHL2*) was enriched for lymphocyte-mediated immunity; Cluster 2 (*GJA1*, *MYH3*, *SCN5A*, and *TNNI3*) for cardiac muscle tissue development and contraction; and Cluster 3 (*FGFR2*, *FLT4*, and *PTPN14*) for lymphangiogenesis. In addition, three specific gene–gene interactions were noted: (*ABCB7*– *ALAS2*) associated with heme biosynthesis, (*MAGED2*–*SLC12A3*) with sodium chloride homeostasis, and (*EFTUD2*–*HNRNPK*) with spliceosome activity. Several of the novel RPL genes (*PRF1* and *TLR8,* for example) and mouse-derived genes (*LYST, PTPN14*) cluster with the known developmental regulators in humans (Figure S4).

To support gene-disease associations, we analyzed single-cell transcriptomic data from human development, spanning from preimplantation through organogenesis and into the second trimester.^26–31^ Expression profiles of novel RPL genes supported developmental relevance (Figures S5, S6, S7). For example, *HNRNPK* and *GJA1* showed broad expression across early tissues, while *PRF1* and others had more restricted, later-stage expression patterns (Figure 3). The expression pattern suggests that defects in broadly expressed genes such *HNRNPK* are more likely to result in early lethality, while variants in genes with restricted expression (e.g., *PRF1*) are more likely to be observed in later losses or postnatal disorders.

**Figure 3.**
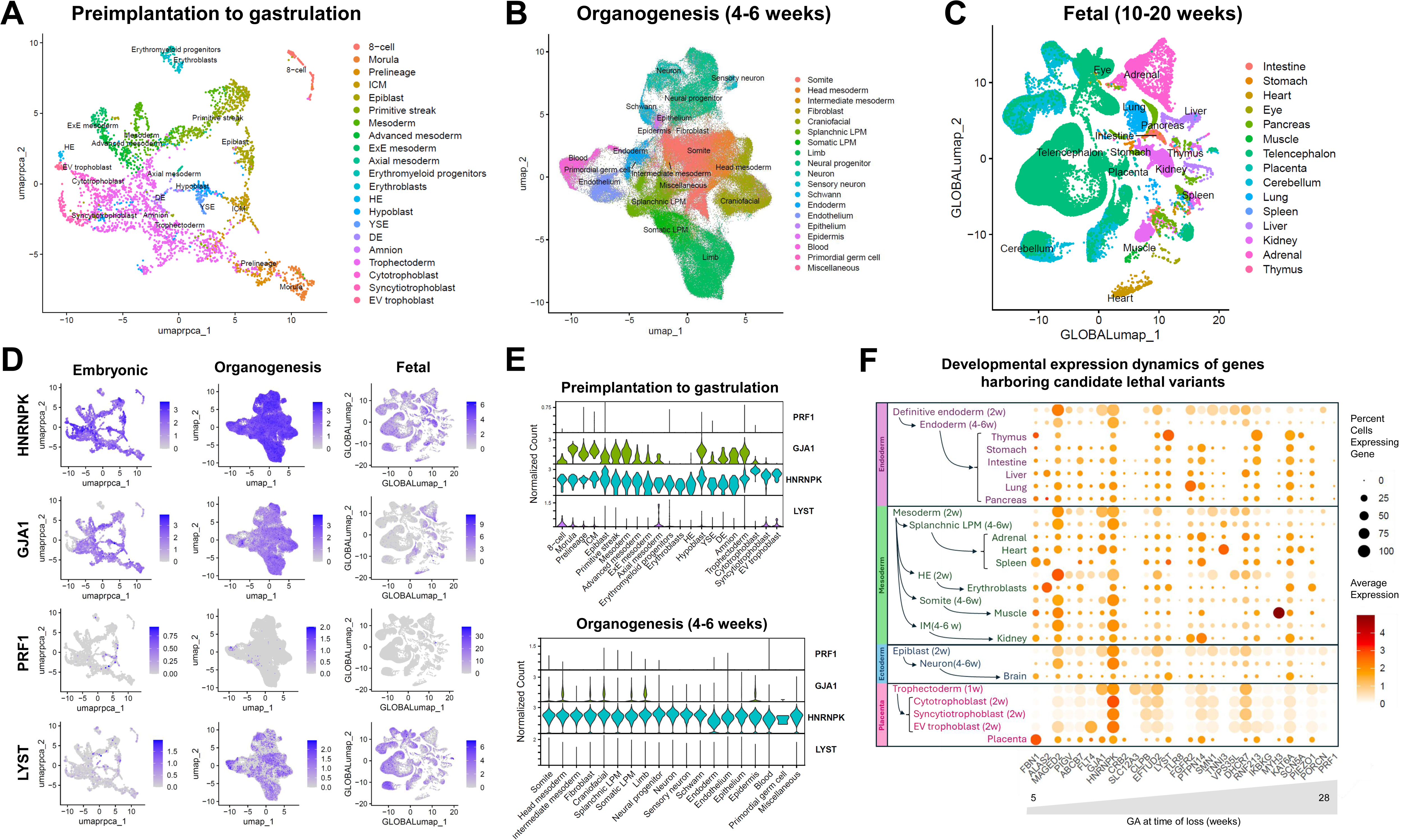
Temporal and Spatial Transcriptomic Expression of Genes Identified in Pregnancy Loss. **Panels A-C**: Uniform Manifold Approximation and Projection (UMAP) visualization of single-cell transcriptomic data from published datasets, spanning the preimplantation to gastrulation stages (1-2 weeks’ gestation),^26–29^ organogenesis (4-6 weeks),^30^ and fetal stages (10-20 weeks).^31^ **Panels D-E**: Log-normalized expression of selected genes, highlighting broad early expression of *HNRNPK* and *GJA1* and more restricted expression of *PRF1* and *LYST*. **Panel F**: Summary of gene expression dynamics from preimplantation through 20 weeks’ gestation for all genes with deleterious variants detected in our cohort. For each timepoint and cell type or organ system, dot size reflects the percentage of expressing cells, and color intensity indicates average log-normalized expression. Genes ordered by increasing gestational age at time of loss for the corresponding POC. LPM: lateral plate mesoderm, IM: intermediate mesoderm. Unless otherwise noted, cell types correspond to the 10–20 week fetal stage. Deleterious variants in genes broadly expressed during early organogenesis (e.g., *HNRNPK)* are more likely to result in embryonic lethality, whereas genes with later or lineage-restricted expression (e.g., *PRF1*) are associated with losses occurring later in gestation or with postnatal metabolic disorders.

## Discussion

We applied trio genome sequencing and a lethality-focused analysis to a cohort of 118 families with unexplained euploid pregnancy losses and identified 39 genomic variants in 28 genes as likely genetic causes of RPL across 30 families (25.4%), including 21 families (17.8%) with P/LP variants in 19 genes, three families (2.5%) with a combination of VUS and P/LP variants in three genes, and six families (5.1%) with VUS in six genes. Among 123 sequenced POC samples, genome sequencing yielded positive results in 21 out of 97 (21.6%) first trimester losses, 6 out of 22 (27.7%) second trimester losses, and in three out of four POCs of the third trimester/neonatal deaths cases. Genes previously associated with neonatal or prenatal/perinatal lethality, such as *CLPB*, *CRB2*, *DHCR7*, *FBN1*, *FGFR2*, *FLT4*, *MAGED2*, *PIGV*, *SMN1* were also identified in the first-trimester miscarriages in our study.^23,32–36^ The *ABCB7, ALAS2, EFTUD2,* and *HNRNPK* genes have not been previously linked to the first trimester miscarriages, although variants in these genes have been identified in case reports of families with pregnancy losses.^18,37–39^ Our study also discovered single nucleotide variants in eight additional genes not previously associated with the first trimester miscarriages, including *GJA1, LYST, PTPN14, RNF213, SLC12A3, TLR8, TNNI3,* and *VPS35L* genes. These findings highlight a spectrum of lethal gene effects spanning multiple stages of human development. The genetic architecture of RPL overlaps with pediatric genetic disorders and preterm birth,^40^ and extends to additional genes essential for embryo and early fetal development. Variants were also identified in genes that have not previously been associated with human disease - *FHL2*, *ADAMTS16*, *NCAPG*, *RIIAD1*, *NCKIPSD*, *SRRD*, *IRX2* (Table S8), demonstrating the value of genome-wide approach in uncovering novel candidate embryonic lethal variants.

Inherited variants predominated in our cohort: in ∼83% of losses diagnostic findings involved variants carried by a parent, including 37% POCs with biallelic variants. This supports the concept that individuals harbor lethal recessive alleles that only manifest in the biallelic state during embryogenesis and aligns with predictions that each person carries 1–2 recessive lethal variants.^22^ As seen in Smith-Lemli-Opitz syndrome (*DHCR7* gene), expected disease incidence based on carrier frequency exceeds the observed birth rates, likely due to unrecognized embryonic demise. Our study confirms this directly, identifying pathogenic *DHCR7* compound heterozygotes among miscarried fetuses. Copy number variants in the *SMN1/SMN2* locus, which cause spinal muscular atrophy, may also lead to first trimester miscarriages as found in our study (PL-098D, Table 1), however further investigation is required to establish this novel gene-disease association. It is interesting to note that both *DHCR7* and *SMN1* show the highest frequency rates among genes associated with lethality, but their involvement in first trimester losses is rarely documented.^22^

We identified germline parental mosaicism in two families with variants in the *FGFR2* and *PORCN* genes, which were initially classified as *de novo* (Table 1). Parental mosaicism has been reported in 1–10% of families with children affected by conditions caused by *de novo* variants^41,42^ and may similarly contribute to a substantial proportion of unexplained pregnancy losses. Recognizing this possibility is important for accurate recurrence risk counseling. In many of our cases, only a single pregnancy loss sample was available for analysis. It is likely that gonadal mosaicism is more prevalent in recurrent loss, but confirming its presence, particularly in maternal gonads, is challenging due to limited accessibility of ovarian tissue for testing.

We also found that autosomal dominant conditions caused by variants in *CLPB*, *FBN1*, *FLT4*, *MYH3*, *TNNI3*, and *SCN5A* genes may present with embryonic lethality. Some variants, typically linked to conditions with a late-onset or variable expressivity (e.g., in *FBN1*, *FLT4, SCN5A*), were inherited from healthy parents but can lead to lethality and/or placentation problems during early embryo development.^35,43–46^ Notably, variable expressivity and incomplete penetrance have been documented for *FLT4* and *SCN5A.*^43,44^ Similarly, families with *FBN1* variants have shown a wide range of outcomes, from stillbirth to neonatal death, further underlining the spectrum of severity in these dominant conditions.^35,45,46^ This highlights how the phenotypic effects of specific variants can be more severe prenatally than postnatally. Previous studies were often based on sequencing the POC alone, lacking parental inheritance data or excluding inherited variants in genes associated with autosomal dominant conditions, which can hinder the discovery of imprinting disorders, sex-specific or parent-of-origin effects for specific genetic variants.^17,35,47–49^

In our cohort, clinical manifestations beyond demise were absent in 95% of cases. Cross-species data, developmental transcriptomics, and protein–protein interaction (PPI) analysis strengthened variant interpretation in the absence of overt fetal anomalies. Ten genes (*ALAS2, CRB2, FGFR2, FLT4, GJA1, HNRNPK, IKBKG, MYH3, PTPN14*, and *VPS35L)* with variants in our cohort are essential for murine embryonic survival, with expression patterns closely aligned with the timing of pregnancy loss (Table 2, Figures S5, S6). Additionally, 13 genes (*ABCB7, CRB2, DHCR7, EFTUD2, FBN1, FGFR2, KAT6A, LYST, PIEZO1, PTPN14, SCN5A, SMN1*, and *TNNI3*) are essential for murine preweaning or pre/postnatal survival, with human developmental expression patterns corresponding to the gestational age at loss (Table 2, Figure S7). For example, a *PRF1* homozygous variant, affecting a gene with minimal expression during early embryogenesis but higher expression later in gestation, was identified in a 28-week loss, consistent with its known neonatal-onset phenotype (Figure 3). By contrast, *GJA1* is expressed from the morula stage and was implicated in an 8-week loss, consistent with mouse data showing perinatal lethality.

Next-generation sequencing has been applied to pregnancy loss, maternal predisposition to RPL,^50^ and stillbirth through various study designs, yielding a wide range of diagnostic outcomes.^16,17,34,36,51^ Stillbirth studies (typically losses ≥20 weeks) have reported relatively low yields. For instance, exome sequencing in stillbirth cases with normal karyotype and microarray finds causative variants in only 6.1% of cases.^17^ This low yield underscores that many single-gene disorders leading to stillbirths in the absence of obvious fetal anomalies are unknown. In contrast, trio exome studies on euploid fetuses with structural anomalies detected by ultrasound uncover pathogenic variants in ∼30% of cases.^16,34,51^ Prior studies of pregnancy-loss families frequently used small cohorts enriched with chromosomally abnormal products of conception, elective terminations, second-trimester losses with prenatal ultrasound anomalies or abnormal autopsy findings, and consanguineous families, which may have contributed to higher diagnostic yields.^16,34,51–54^

The genetic basis of first-trimester, structurally normal, euploid RPL remains largely unexplored. Only a few sequencing studies have specifically focused on recurrent euploid miscarriages, and collectively, have only analyzed a few dozen of POC samples.^47,55–57^ Notably, trio exome sequencing studies examining miscarriages from consanguineous couples with RPL or a specific phenotype such as nonimmune hydrops fetalis found potential P/LP disease-causing variants in ∼37-45% of cases.^53,54^ Other RPL studies analyzing four to eight families have focused on disease-causing genes associated with autosomal recessive conditions and reported P/LP variants in 0-57% of euploid POCs.^18,47,55,56^ An aggregated analysis of ∼60 POCs across RPL studies suggested P/LP variants in over half of cases, however the association of the reported genes with miscarriage is not evident.^18,47,51,53,55–57^ Among these studies, inherited variants in two genes *FLT4* and *PIEZO1* were observed more than once, while other miscarriages with positive results inherited biparental variants in different genes.^18,53^

By applying a comprehensive lethality-centric approach, incorporating variant segregation, gene intolerance metrics, phenotype databases, and developmental expression, we demonstrated a diagnostic yield comparable to anomaly-driven prenatal studies.^36^ We identified P/LP variants in at least 17.8% of families, all of which had normal karyotypes and were mostly the first-trimester losses. This yield is substantially higher than the ∼6–12% yield reported in unexplained stillbirth series,^17,49^ but lower than in cohorts of fetuses with recognizable phenotypes.^16,34,35^ This is expected, since our cohort consisted mostly of structurally normal miscarriages, cases that inherently less likely to have recognizable Mendelian lethal phenotypes. Beyond the overall yield, our study expanded the spectrum of genes implicated in early embryonic and fetal lethality. We detected P/LP variants in 19 genes and identified variants of uncertain significance in 16 genes among our cases. Importantly, 11 genes (*ALAS2, CLPB, DHCR7, EFTUD2, HNRNPK, PIGV, SMN1, FLT4, SLC12A3, TLR8,* and *TNNI3)* with P/LP variants in our cohort have not been previously linked to first-trimester miscarriages and represent novel associations with human embryonic demise.

Pregnancy loss is a highly heterogeneous condition, presenting a greater challenge to genetic diagnosis. A comprehensive review of literature and the Online Mendelian Inheritance in Man (*OMIM*) database queries identified 934 genes (rpldb.org/intolerome/pldb) associated with prenatal, perinatal, or infantile death. In addition, 2,871 candidate lethal genes were suggested by mouse knockout models.^22,23^ It is not surprising that only a few genes consistently reported among the POC studies such as *PIEZO1*, *FLT4, KAT6A, HNRNPK, SCN5A, FGFR2,* and *FBN1,*^18,33–36,53^ whereas other genes are unique. Each affected family may harbor a pathogenic variant in a different gene, and in cases when partners carry lethal variants in multiple genes, the cause of miscarriage might be different for each POC. Analysis of both POC and liveborn children in families with RPL may uncover novel and rare genetic causes and help in a systematic accumulation of data about lethal variants.

In summary, our work demonstrates the diagnostic yield of genome sequencing for RPL in similar range to other diseases and supports its practical utility. We show that even in the absence of fetal structural anomalies, genomic analysis can identify underlying lethal variants in a substantial fraction of recurrent losses. By integrating fetal–parent trio sequencing with cross-disciplinary developmental data, we were able to pinpoint both known disease genes acting in a new context (early pregnancy) and completely novel genes essential for human viability. These findings expand the set of genes known to cause early embryonic demise and provide insights into the diverse biological pathways whose disruption leads to miscarriage. Equally important, our study supports expanding current diagnostic approaches for RPL beyond conventional karyotyping and microarrays to include exome and genome sequencing. An expanded genomic approach in RPL can identify genetic etiology for couples’ losses, which in turn can guide clinical management – for example, informing the use of preimplantation genetic testing (PGT) to select embryos without the lethal variant and enabling targeted preconception carrier screening for at-risk couples. Ultimately, better genomic characterization of RPL will improve our ability to counsel families, implement preventive strategies, and provide more personalized care to those experiencing the heartbreak of recurrent miscarriage.

## Supporting information

ONLINE METHODS - Genetic Variants in Recurrent Euploid Pregnancy Loss

Response to Medrxiv reviewers comments

## Data Availability

All data produced in the present study are available upon reasonable request to the authors.

