## Supplementary material for "Genetic Variants in Recurrent Euploid Pregnancy Loss": ONLINE METHODS - Genetic Variants in Recurrent Euploid Pregnancy Loss

**Study design and participants**

We conducted a multicenter “Trio analysis of Recurrent pregnancy loss Integrated bioinformatics GenOmics Study” (TRIOS) at five academic centers (Stanford University, University of California San Francisco, University of Pittsburgh, Oregon Health Sciences University, and University of Illinois at Chicago). This study received ethical approval from the Institutional Review Boards (IRBs) of UCSF (IRB#21-35237), Stanford (IRB#60431), UPMC (IRB#STUDY20050099), OHSU (IRB#23569), UIC (IRB#2013-0029-MOD002) and a central IRB from the Center for IRB Intelligence (CIRBI, Advarra, Columbia, MD) (protocol ID: Pro00054839) for prospective recruitment and inclusion of the previously enrolled participants. Natera, Inc. provided recruitment support by identifying potential participants through their miscarriage testing for patients with RPL and euploid results on products of conception (POC). All participants gave written informed consent. Inclusion required ≥2 pregnancy losses with no identified cause. Exclusion criteria included known etiologies of pregnancy loss such as parental structural chromosome rearrangements, untreated hormonal and endocrine conditions, uterine anomalies and cervical insufficiency, diagnosis of an autoimmune disease, such as antiphospholipid syndrome, fetal infections. All couples underwent screening for known RPL causes, and any with identified causes were excluded from the study (Figure 1). Clinical data were collected via chart review and questionnaires and managed in REDCap (Table S1, Figure S1, Figure S2). Recruited couples had between two to 12 miscarriages lost pre/perinatally, and a history of zero to three liveborn (Table S1 and Table S6).

**Library Preparation and Genome Sequencing**

Genomic DNA from POC and parental samples (blood, saliva, urine, semen) was extracted using QIAamp kits (Qiagen). Genome sequencing libraries were prepared using the Kapa EvoPlus V2 library preparation kit (Roche). Genome sequencing was done at a CLIA-certified lab via NGS technology using NovaSeq 6000 sequencer (Illumina) (30× average depth, >90% bases ≥20×). Reads were aligned to GRCh38, and quality metrics met clinical standards. Alignment and variant calling were performed using the Illumina DRAGEN v.4.0.3 (The Dynamic Read Analysis for GENomics) pipeline to assess for single nucleotide variants (SNVs), short insertion and deletion variants (indels), mitochondrial genome variants, structural (SVs) and copy number variants (CNV) involving genomic regions with unique DNA sequences.

**Variant analysis and interpretation**

Variant calling and interpretation used MOON (Invitae, San Francisco, CA) and custom pipelines (Figure 1B). MOON employs a prioritization and filtering approach based on Human Phenotype Ontology (HPO) terms. The variant interpretation is based on relevant HPO terms. We specifically selected the most relevant HPO terms and genes associated with pregnancy loss and pre/postnatal lethality, encompassing various conditions in both the mother (Tables S2 and S3) and POC (Table S4.)

**Pipeline for analysis of variants in pregnancy loss samples**

Germline single nucleotide variants (SNVs) and structural variants (SVs) were identified for each sample. Variants with MAF ≤0.1% were retained, curated, and classified using the ACMG guidelines. For SNVs, we included all variants with a minimal depth of 8X, genotype quality equal to or more than 40. We applied rigorous filters, excluding variants with an allele fraction below 20% and reference allele coverage less than 10X. Notably, all low-quality variants were excluded from the subsequent analysis, ensuring a reliable examination of the identified genetic variations. Furthermore, to analyze *de novo* heterozygous variants, we implemented one more filter based on the gnomADv4 database, rejecting variants for which frequency is not 0. Different modes of inheritance, including autosomal dominant, autosomal recessive, and X-linked, were investigated. Variants were classified as pathogenic (P), likely pathogenic (LP), or variants of uncertain significance (VUS), and designated as RPL-relevant if consistent with pregnancy loss etiology.

To streamline variant interpretation, we implemented a tier-wised approach applying: 1) the Human Phenotype Ontology (HPO)-based analysis using a set of virtual human phenotype ontology terms relevant to lethal conditions (Table S4); 2) the predesigned virtual gene panels comprising the known human, emerging, and mouse-derived lethal candidate genes; and 3) a subsequent comprehensive genome assessment to identify novel RPL-causing genes.

**Embryonic and fetal transcriptomic profiling**

For transcriptomic analysis of genes harboring candidate lethal variants, published single cell RNA sequencing datasets were downloaded.^5-10^ For the 10-20 weeks expression,^10^ a subset of the original dataset was downloaded from the CZ CELLxGENE webtool (<https://cellxgene.cziscience.com/datasets>) and filtered to retain only cells corresponding to fetuses with euploid karyotype. Preimplantation and gastrulation stage datasets were integrated, embryonic day 3 (E3) and E4 cells were annotated as 8-cell and morula, respectively. Stage E5 epiblast and hypoblast were relabeled as inner cell mass (ICM). Mitochondrial gene expression was regressed out using SCTransform, and then datasets were integrated by the anchor-based Reciprocal PCA (RPCA) method. Then, the integrated preimplantation and gastrulation stage dataset, the organogenesis stage dataset and the fetal stage dataset were filtered to retain cells with a minimum read count of 2000 to account for differences in sequencing depth. For each dataset, UMAP was used to generate reduced-dimensionality representations, and gene expression counts were log-normalized for downstream analysis.
