## SUPPLEMENTARY Appendix for "Genetic Variants in Recurrent Euploid Pregnancy Loss"

**Content**

| INVESTIGATORS …………………………………………………………………………. | Page 2 |
| --- | --- |
| CONTRIBUTORS…………………………………………………………………………... | Page 2 |
| PROVIDERS THAT REFERRED PARTICIPANTS………………………….…………… | Page 2 |
| SUPPLEMENTARY METHODS………………………………………………….. | Page 3 |
| Virtual gene panels for analysis of variants in pregnancy loss samples……………...  Known lethal genes  Emerging human candidate genes  Mouse-derived candidate lethal genes  Novel candidate genes | Page 3 |
| Confirmatory testing………………………………………………………………… | Page 4 |
| SUPPLEMENTARY TABLES | Excel file |
| Supplementary Table S1. Study cohort | Excel file |
| Supplementary Table S2. Relevant HPO terms for variants prioritization in maternal genomes | Excel file |
| Supplementary Table S3. RPL-susceptibility genes | Excel file |
| Supplementary Table S4. HPO terms for POC analysis and variants prioritization | Excel file |
| Supplementary Table S5. Mouse-derived candidate lethal genes | Excel file |
| Supplementary Table S6. Summary of variants detected in POC | Excel file |
| Supplementary Table S7. Gene categories | Excel file |
| Supplementary Table S8. Variants in genes with no disease association | Excel file |
| SUPPLEMENTARY RESULTS…………………………………………………… | Page 4 |
| Summary findings in each family…………………………………………………... | Page 4 |
| SUPPLEMENTARY FIGURES……………………………………………………. | Page 16 |
| Figure S1. Number of miscarriages in the study cohort…………….………………. | Page 16 |
| Figure S2. **Demographic and sample characteristics**……………….……………….. | Page 17 |
| Figure S3-1. Findings in the *ABCB7* gene………………………….………………. | Page 18 |
| Figure S3-2. Findings in the *ALAS2* gene…………………………..………………. | Page 19 |
| Figure S3-3. Findings in the *CLPB* gene…………………………………………….. | Page 20 |
| Figure S3-4. Findings in the *CRB2* gene……………………………………………. | Page 21 |
| Figure S3-5. Findings in the *DHCR7* gene………………………………………….. | Page 22 |
| Figure S3-6. Findings in the *EFTUD2* gene………………………..……………….. | Page 23 |
| Figure S3-7. Findings in the *FBN1* gene……………………………………………. | Page 24 |
| Figure S3-8. Findings in the *FGFR2* gene………………………….………………. | Page 25 |
| Figure S3-9. Findings in the *FLT4* gene…………………………………………….. | Page 26 |
| Figure S3-10. Findings in the *GJA1* gene…………………………..………………. | Page 27 |
| Figure S3-11. Findings in the *HNRNPK* gene……………………...……………….. | Page 28 |
| Figure S3-12. Findings in the *IKBKG* gene…………………….…..………………. | Page 29 |
| Figure S3-13. Findings in the *KAT6A* gene……………...……….…..……………... | Page 30 |
| Figure S3-14. Findings in the *LYST* gene…………………………....…………….... | Page 31 |
| Figure S3-15. Findings in the *MAGED2* gene……………………....…………….... | Page 32 |
| Figure S3-16. Findings in the *MYH3* gene…………………………..…………….... | Page 33 |
| Figure S3-17. Findings in the *PIEZO1* gene……………………..….…………….... | Page 34 |
| Figure S3-18. Findings in the *PIGV* gene…………………………...…………….... | Page 35 |
| Figure S3-19. Findings in the *PORCN* gene………………………...…………….... | Page 36 |
| Figure S3-20. Findings in the *PRF1* gene…………………………...…………….... | Page 37 |
| Figure S3-21. Findings in the *PTPN14* gene………………………..…………….... | Page 38 |
| Figure S3-22. Findings in the *RNF213* gene………………………...…………….... | Page 39 |
| Figure S3-23. Findings in the *SCN5A* gene………………………....…………….... | Page 40 |
| Figure S3-24. Findings in the *SLC12A3* gene……………………….…………….... | Page 41 |
| Figure S3-25. Findings in the *SMN1* gene…………………………..…………….... | Page 42 |
| Figure S3-26. Findings in the *TLR8* gene…………………………...…………….... | Page 43 |
| Figure S3-27. Findings in the *TNNI3* gene………………………….…………….... | Page 44 |
| Figure S3-28. Findings in the *VPS35L* gene………………….……..…………….... | Page 45 |
| Figure S4. Protein-protein interaction between the genes found in a cohort of pregnancy loss samples. …………………………………………………...……….. | Page 46 |
| Figure S5. Transcriptomic profiles of identified genes during preimplantation to gastrulation. ……………………..……………………………….……………......... | Page 47 |
| Figure S6. Transcriptomic profiles of identified genes during organogenesis (4-6 weeks). ……………………..……………………..…………….……………........... | Page 48 |
| Figure S7. Transcriptomic profiles of identified genes during fetal stage (10-20 weeks)..……………………..……………………..………………….. .…………… | Page 49 |
| SUPPLEMENTARY REFERENCES ……………………..……………………….. | Page 50 |

**INVESTIGATORS**

Mona Aminbeidokhti, Michelle Halstead, Marta Rodriquez-Escriba, Christina G. Tise, Jonathan A. Bernstein, Hakan Cakmak, Elizabeth Pollard, Linda C. Giudice, Katrina Merrion, Gary M. Shaw, Alison Edelman, Maureen Baldwin, David K Stevenson, Mary Stephenson, Michael P. Snyder, Marina Sirota, Ruth B. Lathi, Svetlana A. Yatsenko, Aleksandar Rajkovic.

**CONTRIBUTORS**

We are grateful to the study participants and their families involved in this project, the Recurrent Pregnancy Loss Association (rplassociation.org), team members and supporting personnel for technical and bioinformatic assistance at the recruitment sites: University of California San Francisco (pregnancyloss.ucsf.edu) - Amrita Nagasuri, Bianca Howell, Tomiko Oskotsky, Bahar Yilmaz, Pierre-Marie Martin; Stanford University (pregnancylossanswers.org) - Nisha Udupa, Alma Gonzalez, Kevin Tran, Eda Karagoz, Sophia Adelson, Marisol Maturino, Melissa Rodriguez, John Z. Cao, Virginia Winn, Somaz Chamanara, Durga Thota; Oregon Health & Science University - Lily Angima, Kaitlin Schrote, Corinne Wilcox; Magee Womens Research Institute, University of Pittsburgh - Brittany Mcvicar, Jess Rizzo, Staff at the UPMC Clinical Genomics and Cytogenetics Laboratories.

**PROVIDERS THAT REFERRED PARTICIPANTS**

| University of California San Francisco | Yanett Anaya, Marcelle Cedars, Victor Fujimoto, Eleni Greenwood Jaswa, David Huang, Heather Huddleston, Evelyn Mok-Lin, Paulo Rinaudo, Mitchell Rosen, Maren Shapiro |
| --- | --- |
| Stanford University | Lusine Aghajanova, Ruben Alvero, Stephanie Amaya, Katherine Bianco, Michael Eisenberg, Julia Josowitz, Ronit Mazzoni, Amin Milki, Brent Monseur, Steven Nakajima, Greysha Rivera-Cruz, Brindha Saravanabavanandhan, Robyn Service, Kate Shaw, Jade Shorter, Anna Sokalska, Bo Yu, Fiona Wang, LPCH Prenatal Genetic Counselors |
| University of Pittsburgh Medical Center | Anna Binstock, Katherine Bunge, Beatrice Chen, Eric Fackler, John Fisch, Luanne M. Fraer, Roy Handelsman, Renata Hoca, Monica Kao, Elizabeth Krans, Sami Makaroun, Jennifer Makin, Gretchen Reinhart, Erin Rhinehart, Julie Rios, Ira Rock, Joseph Sanfilippo, Elizabeth Sheehan, Mary Travis, Samantha Erin Vilano, Suji Uhm, Glenn Updike, Division of Maternal Fetal Medicine, Division of Reproductive Endocrinology and Infertility, Fetal Diagnosis and Treatment Center |
| Allegheny Health Network | Meredith Snook, Center for Reproductive Medicine |
| Oregon Health & Science University | Generalist Ob/Gyn Division, Maternal Fetal Medicine Division, Complex Family Planning Division, University Fertility Clinic, Hillsboro Medical Center 8^th^ Ave Women's Clinic |

**SUPPLEMENTARY METHODS**

**Virtual gene panels for analysis of variants in pregnancy loss samples**

**Known human lethal genes**

Among the approximately 20,000 human genes, only about 25% are linked to any known phenotype, most of which are related to postnatal manifestations. The OMIM queries reveled 624 genes (3.7%) associated with lethal phenotypes, which can lead to mortality during fetal development, the perinatal period, or childhood.^1,2^ Some affected individuals with pathogenic variants in these 624 lethal genes might survive longer, depending on the type of the variant and severity of the disease.

**Emerging human candidate lethal genes**

Not all genes associated with pregnancy loss, stillbirth, and neonatal death are currently reported as lethal in humans in the OMIM database. Based on a literature search, we identified an additional 312 genes that have been identified by recent studies, which we term emerging human lethal genes. A combined list of known and emerging human lethal genes, which we refer to as the “Intolerome” is available at [rpldb.org/intolerome](https://rpldb.org/intolerome/)/.

**Mouse-derived candidate lethal genes**

The actual number of human genes essential for fetal viability surpasses the currently identified lethal and emerging candidate genes. It has been estimated that approximately 3,400 human genes are essential for embryonic and fetal development.^1^ Based on mouse knockout phenotypes extracted from the Mouse Genome Informatics (MGI) and International Mouse Phenotyping Consortium (IMPC) databases and the essential organogenesis pathways, 3,684 (39.2%) of mouse genes are associated with lethality.^1,3^ In our previous study, we analyzed allele frequency of pathogenic/likely pathogenic and loss-of-function variants in human orthologs corresponding to mouse lethal genes, and generated a list of 2,612 human genes biallelic variants in which may result in human lethality.^4^ In addition, among healthy individuals, no heterozygous loss-of-function variants were observed in 259 genes that might be associated with autosomal dominant lethal conditions in humans. Variants in the set of 2,871 mouse-derived candidate lethal genes that were identified in POC samples (Table S5) have been thoroughly evaluated.

**Novel candidate genes**

Human-specific genes as well as other genes may have essential function during the human embryonic development, that cannot be predicted from phenotype observed in animal models. Other plausible genes (with lethality evidence/potential) that are not included in the above categories were considered as novel RPL-causing genes.

**Confirmatory testing**

Variants in miscarriage tissue and/or parents were corroborated by alternative methodologies, including Sanger sequencing, digital droplet PCR (ddPCR), chromosomal microarray analysis and Optical genome mapping. Sanger sequencing analysis was performed using Geneious Prime software (version 2024.0.4). The possibility of parental germline mosaicism has been raised in cases with *de novo* variants in genes associated with autosomal dominant and X-linked dominant inheritance. Parental DNA from blood and alternative specimens (saliva, urine and semen), were tested using a high coverage (100X+) exome sequencing or ddPCR to rule out mosaicism.

**SUPPLEMENTARY RESULTS**

**Summary findings in each family**

The lymphatic network serves as a transport system passing fluid, cells, and macromolecules from tissue back into the central circulation. Disturbance in the development or function in the lymphatic system can lead to local or generalized lymphedema, increased hydrodynamic pressure, organ failure, inflammation, and ischemic shock in the affected areas. Multiple genes are known to cause abnormal lymphangiogenesis: ADAMTS3, BRAF, CCBE1, EPHB4, FAT4, FLT4, FOXC2, GATA2, GJA1, GJC2, HGF, IKBKG, KIF11, MAP2K1, MAP2K2, PIEZO1, PIK3CA, *PRF1*, PTPN11, PTPN14, RASA, RASA1, RIT1, SOS1, SOX18, TSC1, TSC2, and *VEGF.^11^* In this study, in 7/30 (23%) POC samples variants were present in 6 lymphedema-causing genes: *FLT4* (Figure S3-9)*,* GJA1 (Figure S3-10), IKBKG (Figure S3-12), PIEZO1 (Figure S3-17), *PRF1* (Figure S3-20)*,* and *PTPN14* (Figure S3-21)*.*

**Findings in the *ABCB7* and *ALAS2* genes (Families PL-130 and PL-104)**

X-linked congenital sideroblastic anemia is a rare condition caused by pathogenic variants in genes involved in heme biosynthesis pathway and iron/ sulfur cluster formation, including *ABCB7* (ATP binding cassette subfamily B member 7; OMIM*300135) and *ALAS2* (5'-aminolevulinate synthase 2, OMIM*301300) genes. During embryonic and fetal stages of development, genes involved in heme synthesis are mostly expressed in erythroid cells, hepatocytes and spleen (Supplemental Figures S5, S6, S7). Affected individuals, typically males, have the pathological accumulation of iron in the mitochondria of erythroblasts, inefficient erythropoiesis and microcytic anemia of variable severity.

We identified two POC samples (PL-130C and PL-104C) with maternally inherited hemizygous variants in the *ABCB7* and *ALAS2* genes, respectively (Figures S3-1 and S3-2). The *ABCB7* gene is a member of the adenosine triphosphate-binding cassette transporter superfamily that plays a central role in the transport of heme from the mitochondria to the cytosol and the maturation of cytosolic iron/sulfur cluster-containing proteins. *ALAS2*, which encodes an erythroid-specific enzyme that localizes to mitochondria, catalyzes the first step in the heme biosynthetic pathway. Male lethality and pregnancy losses have been documented in families affected by X-linked sideroblastic anemia.^12-14^ Studies of murine models with ABCB7 deficiency demonstrated early embryonic lethality of male mice as well as female mice with maternally inherited *Abcb7* deletion. In mice, ABCB7 was found to be essential in extraembryonic tissues where the maternal X chromosome is preferentially active.^15^ This could suggest that defects in placental function could also underly embryonic lethality in these cases.

**Findings in the *CLPB* gene (Family PL-013)**

The *CLPB* (Caseinolytic peptidase B family mitochondrial disaggregase; OMIM*616254) gene belongs to the superfamily of the AAA+ proteins, which form ring-shaped homohexamers with highly conserved ATPase domains. In the PL-013 family (Figure S3-3), we found a pathogenic, paternally inherited, missense variant in the POC, that previously was reported in babies with neurologic problems and neutropenia.^16^ Variants in the *CLPB* gene have been associated with both autosomal dominant and autosomal recessive types of 3-methylglutaconic aciduria with severe congenital neutropenia. It has been shown that missense variants affecting the ATP-binding domain act in a dominant-negative manner.^17^ The CLPB deficiency is characterized by significant clinical variability; manifestations range from severe to mild and asymptomatic forms.^18^ Severe forms are associated with early childhood death secondary to impaired mitochondrial function and defective granulocyte differentiation.^19,20^ Prenatal presentations also include brain atrophy and movement disorder.^17^ It is unclear if severity of CLPB deficiency is modulated by additional factors or if this condition is associated with incomplete penetrance.^21^

**Findings in the *CRB2* gene (Family PL-078)**

The *CRB2* (crumbs cell polarity complex component 2; OMIM*609720) gene produces a polarity complex protein that is part of the Crumbs family and plays a fundamental role in regulating apical polarity during development. Homozygous or compound heterozygous variants in *CRB2* are associated with both steroid resistant nephrotic syndrome and prenatal onset ventriculomegaly with cystic kidney disease, manifesting with early onset in-utero dilated cerebral ventricles and cysts in renal tubules.^22^ In prenatal cases, combinations of frameshift and missense variants or two missense variants in *CRB2* have been reported.^22-25^ Postnatal death at 7 months of age was reported in an infant with a homozygous missense variant in *CRB2*.^22^ Prenatal onset ventriculomegaly with cystic kidney disease also has been reported in a stillborn baby at 22 weeks’ gestation, with a history of pregnancy losses in the family, none of which were tested for *CRB2* variants.^26^

In PL-078 family (Figure S3-4), two *CRB2* variants were identified in the POC, consisting of a maternal frameshift variant and a paternal splice region variant. The maternal variant has been previously reported in individuals with steroid-resistant nephrotic syndrome^27^ and in affected siblings with prenatal-onset ventriculomegaly and kidney disease, where both pregnancies were terminated due to severe congenital hydrocephalus and urinary tract anomalies.^28^ The paternal splice region variant is rare, with an allele frequency of 0.0001 in gnomADv4, and has been predicted by in silico tools such as SpliceAI to introduce a new splice acceptor site (acceptor gain **Δ score = 0.96),** leading to a premature stop codon in exon 3 and loss of protein function^29^ (https://spliceailookup. broadinstitute.org/). This combination of two loss-of-function variants has not been reported before in this gene, and this case represents the first report of early pregnancy loss under these circumstances. The severity of the variants may explain why the pregnancy ended at an earlier stage (8 weeks’ gestation) compared to previously documented cases, which typically showed ultrasound abnormalities in later stages of gestation and elective termination. Findings from animal models further support the essential role of this gene in early embryonic development. In mouse studies, knockout of *Crb2* resulted in disturbed polarity of epiblast cells at the primitive streak, severe defects in gastrulation, impaired organ formation, and embryonic lethality by embryonic day 12.5.^30^ These observations may suggest that biallelic loss of function in this gene may be incompatible with human development and could contribute to early pregnancy loss.

**Prenatal lethality due to biallelic variants in the *DHCR7* gene (Family PL-084)**

Smith–Lemli–Opitz syndrome (SLOS) is a rare autosomal recessive disorder caused by variants in the *DHCR7* (OMIM*602858) gene, which encodes the enzyme 7-dehydrocholesterol reductase, resulting in cholesterol deficiency and accumulation of its precursors 7- and 8-dehydrocholesterol in tissues and body fluids. *DHCR7* is expressed during early stages of embryogenesis (Supplemental Figures S5-6), as cholesterol plays a vital role during early development. SLOS exhibits a wide spectrum of phenotypes, ranging from severe manifestations leading to prenatal or postnatal death. Major features include intrauterine growth restriction; microcephaly; variable malformations of brain, limbs, craniofacial abnormalities, cardiac defects, renal and genital anomalies.^31,32^ Cholesterol deficiency affects the sonic hedgehog (SHH) signaling cascade involved in patterning of the limbs, heart, and midline structures of the brain, leading to holoprosencephaly and other malformations in SLOS.

In multiple ethnic populations, the carrier frequency of *DHCR7* variants ranges from 1/40 to 1/140,^33^ yet the average SLOS prevalence of 1 per 50,000 is significantly lower than anticipated.^34,35^ The difference between the *DHCR7* mutation carrier rate and the incidence of SLOS is thought to reflect prenatal loss of conceptions with homozygous or compound heterozygous null mutations.^36^ In our previous study, we also showed that the cumulative frequency of variants in *DHCR7* gene is ≥ 0.5% in the general population which is fairly high.^4^ This condition is also included in a comprehensive preconception carrier screening panel,^37^ however until laboratory standards consider reporting only pathogenic and likely pathogenic variants, alterations in the *DHCR7* gene causing early fetal lethality will remain unrecognized. In the PL-084C, we found compound heterozygous variants inherited from both parents (Figure S3-5).

**Loss-of-function variants in *EFTUD2* cause the first trimester miscarriage (Family PL-101)**

The *EFTUD2* (elongation factor Tu GTP binding domain containing 2; OMIM*603892) gene encodes a GTPase, a component of the spliceosome. The spliceosome is an RNA-protein complex responsible for the removal of non-coding introns from pre-mRNAs and alternative splicing. In embryos, *EFTUD2* is strongly expressed during the first two weeks after fertilization (Figure S5), continued to at least up to 6 weeks of embryonic development in the head, brain, somites, epidermis, and primordial germ cell (Figure S6).

Heterozygous loss-of-function variants and deletions in *EFTUD2* are associated with mandibulofacial dysostosis, an autosomal dominant condition characterized by microcephaly, craniofacial anomalies, neurodevelopmental problems, esophageal atresia, and congenital heart defects. Studies of knockout animal models showed that *EFTUD2* plays a role in maintaining normal osteoblast and chondrocyte proliferation.^38^ Neural crest cell-specific conditional *Eftud2*^-/-^ knockout results in pre-implantation arrest in mice. RNA sequencing revealed that loss of *EFTUD2* has a strong effect on the TP53 signaling pathway activating genes involved in the cell cycle arrest and apoptosis. In addition, the expression of *EFTUD2* was found to correlate with known immune modulatory proteins like pregnancy zone protein (PZP) and human chorionic gonadotropin and is significantly upregulated in the syncytiotrophoblast samples obtained from spontaneous miscarriage.^39^

In the PL-101 family, we found a *de novo* splice donor variant in the POC in intron 12 (Figure S3-6). Recurrent pregnancy loss of three fetuses with a nonsense variant in the exon 12 of the *EFTUD2* gene have been reported previously,^40^ resulted from paternal germline mosaicism. Similarly to our case, spontaneous miscarriages in the affected family occurred before 10 weeks of gestation. The cumulative evidence points to the *EFTUD2* as a plausible cause of pregnancy loss.

**Findings in the *FBN1* gene (Family PL-069)**

The *FBN1* (fibrillin 1; OMIM*134797) gene encodes an extracellular matrix glycoprotein that plays a crucial role in forming calcium-binding microfibrils. These microfibrils provide mechanical support and contribute to the structural stability of both elastic and nonelastic connective tissues throughout the body. Pathogenic variants in *FBN1* are the cause of Marfan syndrome, which involves skeletal, ophthalmologic, pulmonary, and cardiovascular abnormalities often leading to life-threatening complications such as aortic aneurysm. This condition is characterized by high degree of clinical variability, ranging from mild to a severe neonatal disease with multiorgan involvement. *FBN1* variants have been documented in fetal demise at 22-35 weeks’ gestation,^41,42^ and shortly after birth.^43^ Neonatal Marfan syndrome is the most severe form of the disease, and frequently results in fatal outcomes.^44-46^ In mouse models, knockout of *Fbn1* gene causes embryonic and early death.^47-49^

In the PL-069 family (Figure S3-7), we identified a maternally inherited missense variant in the POC, which has been previously reported in individuals with Marfan syndrome.^50,51^ Marfan syndrome exhibits considerable phenotypic variability, both among individuals within the same family and across different families.^52^ Additionally, women with Marfan syndrome have been shown to have a higher rate of early spontaneous abortion compared to controls.^53^ It is possible that the presence of a variant in the mother contributed to an increased risk of early pregnancy loss. Other genes may play a role in modifying the clinical phenotype and risk of pregnancy loss in individuals with *FBN1* variants on chromosome 15. Notably, the male partner in this family carries a balanced inversion involving chromosome 5 confirmed by karyotype. The miscarriage sample ruled to be euploid and negative for the chromosome 5 inversion.

**Pathogenic variants in the *FGFR2* gene (Family PL-026 and PL-060)**

*FGFR2* encodes fibroblast growth factor receptor 2 (OMIM*176943), which plays a crucial role during embryonic development, regulating cell proliferation, apoptosis and differentiation of various types of cells, angiogenesis, organogenesis and bone development. Heterozygous pathogenic variants in the *FGFR2* gene result in diverse phenotypes, such as Apert syndrome, Beare-Stevenson cutis gyrata syndrome, Crouzon syndrome, Jackson-Weiss syndrome, Pfeiffer syndrome, Saethre-Chotzen syndrome, and syndromic craniosynostosis. These autosomal dominant conditions are mainly due to sporadic, de novo variants, although parental germline mosaicism has been documented.

We identified missense variants in two unrelated fetuses: PL-026C and PL-060D (Figure S3-8). Previous studies have identified variants in the *FGFR2* gene that were associated with prenatal lethality, with most clustering in the transmembrane (TM) domain and the immunoglobulin-like domain III (Ig III).^54,55^ In our cohort, the variants found in these two POCs were also localized to the Ig III domain. Parental genome and Sanger sequencing suggested de novo occurrences of FGFR2 variants in both fetuses. To exclude germline mosaicism, we performed high coverage exome sequencing and ddPCR on parental DNA samples from blood, urine, and semen. The blood and urine samples from the mother (PL-026A) were found to be mosaic for the same c.1144T>C variant as found in the PL-026C fetus, ranging from 0.3% in urine to 0.8% in blood. No evidence of parental germline mosaicism for the *FGFR2* variant in the other family (PL-060) was observed, suggesting *de novo* change.

**Mechanisms of *FLT4* related pregnancy loss (Family PL-009 and PL-076)**

FLT4 (vascular endothelial growth factor receptor 3; OMIM *136352) is essential for development of the embryonic cardiovascular system and lymphatic vessels. During embryogenesis, *FLT4* is expressed in vascular endothelial cells and becomes restricted to lymphatic vessels.^56^ Heterozygous *Flt4*^-/wt^ mice have abnormal lymphangiogenesis, and complete loss of FLT4 in mice results in embryonic lethality due to cardiovascular failure.

Multiple exome and genome sequencing studies have reported deleterious variants in FLT4 in association with primary hereditary lymphoedema (Milroy disease), non-immune hydrops fetalis, cardiac defects such as Tetralogy of Fallot (TOF), and with recurrent pregnancy loss. The FLT4 variants can lead to two distinct phenotypes: cardiovascular malformations and lymphoedema, depending on the specific location of a genetic variant within a gene. The kinase activity of FLT4 is a key regulator in the development of blood and lymphatic vascular systems, thus heterozygous variants with a dominant negative effect occurring within the Protein tyrosine Kinase domain have been found in patients with Milroy disease and non-immune hydrops fetalis. In contrast, *FLT4* variants that predispose to cardiac malformations (TOF and septal defects) are protein-truncating changes. Incomplete penetrance and variable expressivity have been observed between and within families with the same alterations, thus pathogenetic variants can be transmitted from a parent to offspring as observed in Pl-009 family with recurrent pregnancy loss (Figure S3-9). It is unclear if *FLT4* change in the mother is also the cause of didelphys uterus and absent left kidney or if her carrier status influences the ability to carry an unaffected pregnancy to term.

**Findings in the *GJA1* gene (Family PL-082)**

The *GJA1* (Gap Junction protein Alpha 1; OMIM*121014) gene encodes the transmembrane, gap junction protein connexin 43 (CX43). *GJA1* is expressed during early embryonic development in almost every tissue (Figure S5), forming clusters of intercellular channels, called Gap junctions, that allow ions, metabolites, and small molecules to pass between adjacent cells.

Alterations in *GJA1* can cause a variety of autosomal dominant conditions, including oculodentodigital dysplasia, lymphedema, sudden infant death syndrome, and heart malformations.^57^ In the PL-082 family, we found a *de novo* missense variant in the POC (Figure S3-10) that is located within the carboxyl terminus (COOH-tail), responsible for post-translational modifications and protein-protein interactions.^58^ Variants in the COOH tail are most commonly associated with heart defects, ranging from mild arrhythmias to severe structural malformations. In addition, *GJA1* is strongly expressed in trophectoderm (Figure S5), and plays an important role in placenta development. Decreased connexin 43 expression has been observed in individuals with recurrent pregnancy loss.^59^

**Findings in the *HNRNPK* gene (Family PL-011)**

The *HNRNPK* (heterogeneous nuclear ribonucleoprotein K; OMIM*600712) gene encodes one of the major mRNA-binding proteins that plays an important role in the regulation of chromatin remodeling, RNA stability and splicing, translation and signal transduction.^60^ During embryonic and fetal stage, *HNRNPK* is highly expressed across all tissues (Supplemental Figures S5, S6, S7). Heterozygous loss-of-function variants in the *HNRNPK* gene are the cause of Au-Kline syndrome, also known as Okamoto syndrome. This autosomal dominant disorder is characterized by multiple congenital malformations, global developmental delay, craniofacial anomalies, heart defects, genitourinary and skeletal abnormalities. Prenatal presentations also include increased nuchal translucency, hydronephrosis, cardiomegaly, non-immune fetal hydrops, and pregnancy loss.^61,62^

In the PL-011 family, we found a frameshift variant in the POC (Figure S3-11). This variant was not present in the mother, however the sperm donor cannot be tested. Together, these results demonstrate that loss of one *HNRNPK* allele causes multiple developmental anomalies and can result in in utero lethality.

**Structural variant in the *IKBKG* gene (Family PL-100)**

The *IKBKG* (inhibitor of nuclear factor kappa B kinase regulatory subunit gamma; OMIM*300248), previously known as NEMO, is a known X-linked gene associated with in utero lethality in males. Along with the other 2 subunits, IKBKA (OMIM*600664) and IKBKB (OMIM*603258), IKBKG forms a complex that controls the NF-kappa-B-induced activation of cytokine-associated genes. Inappropriate function of NF-kappa-B (nuclear factor of kappa B) has been linked to aberrant inflammatory responses, autoimmune conditions, cell apoptosis and growth deficiencies, incompetent development of immune cells.

Heterozygous pathogenic variants in *IKBKG* cause X-linked dominant Incontinentia Pigmenti (IP). This condition is typically lethal in males, while heterozygous females survive due to mosaicism or selective skewed X-inactivation of the affected allele.^63,64^ Due to a repetitive nature of DNA sequences surrounding the *IKBKG* gene, the most common alteration seen in ~80% females with IP is a 11.7-kb deletion of exons 4-10, that occurs *de novo* during paternal spermatogenesis. In our cohort, mother PL-100A (Figure S3-12) had a childhood history suggestive of IP, however the common *IKBKG* deletion rearrangement was not detected using the clinically available MLPA assay. A 32-kb deletion was identified in a male fetus (PL-100C) and two subsequent pregnancy losses. Microarray analysis of the mother revealed a loss in copy number in the Xq28 region comprising the *G6PD* gene and exons 1 and 2 of the *IKBKG* gene. These findings support the general consensus regarding the lethality of male fetuses with pathogenic variants in the *IKBKG* gene.

**Variants in *KAT6A* as novel cause of miscarriage (Family PL-033)**

KAT6A (lysine acetyltransferase 6A, OMIM*601408) is a subunit of histone acetyltransferase complex that regulates chromatin remodeling, transcription of multiple genes, protein translation, and cell proliferation. In mouse models, knockout of *Kat6a*^-/-^ is embryonic lethal due to a failure of hematopoiesis. *KAT6A* is highly expressed during preimplantation and early embryo development stages (Supplemental Figures S5, S6), and plays a key role in cell differentiation. During fetal stages, *KAT6A* is ubiquitously expressed across different tissues including intestine, heart, spleen, lung, and thymus (Figure S7). De novo pathogenic variants in *KAT6A* have been identified as a cause of Arboleda-Tham syndrome, an autosomal dominant condition characterized by intellectual disability, microcephaly, cardiac anomalies, and gastrointestinal problems. Genotype–phenotype analysis revealed that protein-truncating genomic variants involving exons 16–17 are prevalent, especially in patients with more severe manifestations.^62^

In the PL-033 family, we found a *de novo* deletion comprising exons 13-17 as well as a *de novo* missense variant at the deletion breakpoint, that presumed to originate along with the deletion (Figure S3-13). Identification of pathogenic variants in the *KAT6A* gene in prenatal and pregnancy loss cases expanded the phenotypic spectrum of Arboleda-Tham syndrome,^65,66^ to include hepatic and craniofacial anomalies, craniosynostosis, thymus dysplasia, additional cardiovascular findings, and severe hypotonia, supporting the role of *KAT6A* as a cause of fetal demise.

**Findings in the *LYST* gene (Family PL-050)**

Chediak–Higashi syndrome is autosomal recessive condition caused by biallelic mutations in the *LYST* (lysosomal trafficking regulator; OMIM*606897) gene. The LYST protein regulates the vesicular transport into and from the lysosomes and late endosomes. Chediak–Higashi syndrome is a lysosomal storage disorder characterized by immunodeficiency, oculocutaneous albinism, and neurodevelopmental abnormalities. Individuals with this condition have immune dysfunction of variable degree manifesting with hyperinflammation, hepatosplenomegaly, cytopenia, bleeding tendency, recurrent infections, and hemophagocytic lymphohistiocytosis, leading to childhood death in more than 80% of patients.

In the PL-050 family, we found a compound heterozygous variant in the POC inherited from both parents (Figure S3-14). Less than 500 patients affected by Chediak-Higashi syndrome have been reported, therefore its prevalence has been estimated to be less than 1 in 1,000,000 individuals. The observed number of loss-of-function variants in *LYST* is less than expected with a LOEUF score of 0.505, suggesting biological constraint. It is probable that variants in the *LYST* gene are associated with severe manifestations during fetal development, leading to pregnancy loss. Additional studies are needed to establish the role of the *LYST* gene in RPL.

**Findings in the *MAGED2* gene (Family PL-109)**

Pathogenic hemizygous variants the *MAGED2* (MAGE family member D2; OMIM*300470) gene are associated with an X-linked form of transient Bartter syndrome, a fetal renal salt-wasting condition manifesting during the antenatal period and at birth. *MAGED2* is strongly expressed almost in all tissues until 6 weeks of gestation (Supplemental Figures S5, S6, S7), and is known to regulate the function of the sodium chloride cotransporters SLC12A1 and SLC12A3. Bartter syndrome is characterized by high amniotic chloride concentration, polyhydramnios, fetal polyuria with a compensatory hyperaldosteronism, hypokalemic alkalosis, and low blood pressure. Approximately 20% of babies with pathogenic hemizygous variants in *MAGED2* die in utero or shortly after birth secondary to intrauterine growth restriction and prematurity.^67^ There is significant variability in the clinical severity of the disease; in some instances, all symptoms resolved in surviving infants.

In the PL-109 family (quad) analysis, we detected a hemizygous maternally inherited missense variant in one of the tested POC (Figure S3-15). This rare variant (MAF-het = 0.000004964; MAF-hmz = 0.000002524) is located at a well-conserved position and predicted as deleterious.

### ***MYH3* - emerging gene associated with fetal lethality (Family PL-110)**

The *MYH3* gene encodes the embryonic myosin heavy chain (OMIM*160720), a member of the myosin family of motor proteins. Heterozygous pathogenic variants in MYH3 are associated with multiple conditions with great variability of isolated and syndromic phenotypes and incomplete penetrance including Freeman-Sheldon syndrome (OMIM#193700), Sheldon-Hall syndrome (OMIM#618436), arthrogryposis, multiple pterygium syndrome, and spondylo-carpostarsal fusion syndrome (OMIM#178110), characterized by congenital contractures and multiple skeletal anomalies. During fetal development, the *MYH3* gene is predominately expressed at 10 to 20 weeks of gestation in skeletal muscle (Figure S7), although it is also expressed at a lower level in heart and several other tissues. Embryonic myosin forms a dimer by intertwining the tail domains and interacts with other proteins to assemble the functional sarcomere: an essential contractile apparatus of skeletal and cardiac muscle cells. It has been proposed that pathogenic variants in MYH3 alter the TGF-β signaling, leading to contractile dysfunction and associated anomalies. In the PL-110 family, we found a heterozygous variant in the POC, inherited from the mother, who presented with multiple pterygium syndrome (Figure S3-16). Deleterious variants in the *MYH3* gene have been reported in patients with MYH3-associated conditions, individuals with atrial septal defects,^68^ fetuses with cystic hygroma, hydrops fetalis, pleural effusion, flexion contracture, club foot and stillborn babies with spectrum skeleto-muscular phenotypes.^69-71^

### ***PIEZO1* mechanisms in pregnancy loss (Family PL-006)**

The *PIEZO1* (piezo type mechanosensitive ion channel component 1; OMIM *611184) gene encodes an ion channel found in the erythrocyte plasma membrane. *PIEZO1* is expressed in erythroid progenitor cells, fetal liver, and bone marrow and plays a crucial role in erythroid differentiation, epithelial cell crowding and division, and development of vascular and lymphatic structures. The conditions caused by pathogenic biallelic variants in the *PIEZO1* gene include non-immune hydrops fetalis, generalized lymphatic dysplasia, lymphatic malformations, manifesting as fetal anemia (dehydrated stomatocytosis), ascites, cystic hygroma pericardial and/or pleural effusion, generalized lymphedema, polyhydramnios, leading to a subsequent pregnancy loss. In the PL-006 family, we found a compound heterozygous variant in the POC inherited from both parents (Figure S3-17). This family elected for preimplantation genetic testing (PGT-M) for selection of an embryo unaffected by the familial *PIEZO1* variants, resulting in the birth of a healthy child.

**Findings in the *PIGV* gene (Family PL-062)**

The PIGV gene (phosphatidylinositol glycan anchor biosynthesis class V; OMIM*610274) encodes glycosylphosphatidylinositol (GPI) mannosyltransferase 2, an enzyme that plays a crucial role in the biosynthesis of GPI-anchored proteins. The synthesis of the highly conserved GPI anchor backbone occurs through a series of at least nine sequential enzymatic reactions, which are catalyzed by a minimum of 18 distinct proteins including PIGA, PIGN, PIGV, and PIGL.^72^ Specifically, PIGV adds the second mannose residue to the elongating GPI anchor during its biosynthetic process. GPI anchors serve as a key post-translational modification, mediating the attachment of proteins to the cell membrane, thereby defining a class of proteins known as GPI-anchored proteins. Pathogenic variants in PIGV lead to aberrant GPI-anchored protein synthesis, which in turn disrupts essential biological processes, particularly those governing the development and function of multiple organ systems, with a pronounced impact on the nervous system.^72,73^ Hyperphosphatasia mental retardation syndrome (HPMRS), also known as Mabry syndrome, is caused by homozygous and compound heterozygous variants in different proteins from the GPI anchor biosynthesis pathway.^74,75^ Most reported PIGV variants are missense mutations that result in partial enzyme activity, allowing viability despite severe phenotypes. A homozygous missense variant *(*c.905T>C (p.Leu302Pro)*)* has been described in a newborn with cardiac defects and cerebellar abnormalities who died shortly after birth, suggesting that some missense variants can contribute to perinatal lethality.^75^ A loss-of-function variant, including an N-terminal stop-gain change in the last exon of PIGV, in conjunction with a missense variant, was reported in a viable newborn exhibiting severe clinical manifestations. The viability of this severely affected patient may be attributed to the residual enzymatic activity of both the missense and N-terminal stop-gain PIGV variants. Notably, the stop-gain mutation, located in the last exon, may not trigger nonsense-mediated decay (NMD) of the transcript, thereby allowing partial gene function to be retained.^75^

In the PL-062 family, we identified compound heterozygous variants in PIGV inherited from both parents, consisting of a stop-gained mutation and a missense variant, both located in exon 3 of the gene (Figure S3-18). The missense variant in Cys156 which is a highly conserved residue previously reported in individuals with HPMRS in combination with other missense variants.^76,77^ Polyphen-2 in silico tool predicted this missense variant as probably damaging to protein structure (Polyphen-2 score= 0.98). The stop gain variant is predicted to induce NMD, leading to reduced or absent protein product from this allele which may explain the more severe phenotype in this POC. This novel combination of loss-of-function and missense variants expands the phenotypic spectrum of PIGV-related disorders, suggesting a possible association with early prenatal lethality. Reports of pregnancy loss, stillbirth, and postnatal death have been documented in other genes involved in the GPI biosynthesis pathway, including PIGN, PIGA, and PIGL.^78,79^ In mouse models, Piga knockout results in embryonic lethality at E9, underscoring the critical role of GPI anchor biosynthesis in mammalian development.^80^

**Fetal lethality due to variants in the *PORCN* gene (Family PL-007)**

*PORCN* (porcupine O-acyltransferase; OMIM *300651) is a member of the evolutionarily conserved endoplasmic reticulum porcupine family proteins. Porcupine proteins contain multiple transmembrane domains and are essential for Wnt signal activation. *PORCN* deficiency is associated with focal dermal hypoplasia also known as Goltz syndrome, an X-linked dominant disorder characterized by abnormal development of the ectodermal and mesodermal tissues, such as skin, limbs, and eyes. Pathogenic variants in *PORCN* in males are associated with midline thoracic and abdominal wall defects, the limb-body wall complex, and in utero lethality, whereas heterozygous female fetuses display variable defects depending on the distribution of cells with active X chromosome containing the *PORCN* variant and the level of X-inactivation. Pregnancy loss of female fetuses due to *PORCN* alterations is rarely documented.^81,82^ In ~95% of fetuses affected by Goltz syndrome, pathogenic variants in *PORCN* occur *de novo* and in ~5% of cases variants might be transmitted from asymptomatic mothers who either showed a skewed X-inactivation or have mosaicism for the *PORCN* variants.

In the PL-007 family, we found a nonsense variant in the female fetus (PL-007D) with abnormal prenatal ultrasound significant for split hand-feet malformation, facial cleft, cerebellar cyst, and diaphragmatic hernia (Figure S3-19). Analysis of a sample available from a prior pregnancy loss (PL-007C) was suspicious for the same variant, although this DNA sample was found to be heavily contaminated by maternal cells (~80% MCC). High depth exome sequencing and ddPCR were performed using blood, saliva, and urine samples to examine the parents for germline mosaicism. Interestingly, 3% germline mosaicism for the same variant was detected in a urine sample obtained from the father. This is the first report of transmission of an X-linked male-lethal condition from an asymptomatic father due to germline mosaicism.

**Loss-of-function variants in the *PRF1* gene is a novel etiology for pregnancy loss (Family PL-099)**

*PRF1* (perforin 1; OMIM*170280) encodes perforin, a toxin utilized by cytotoxic lymphocytes to destroy virus-infected or cancerous cells. Perforin induces pores in the membranes of targeted cells that are essential for intracytoplasmic injection of pro-apoptotic serine proteases, and a subsequent cell apoptosis. Biallelic loss-of-function variants in the *PRF1* gene are the cause of hemophagocytic lymphohistiocytosis (HLH) clinically characterized by aplastic anemia, edema, hepatosplenomegaly, liver dysfunction, pancytopenia, coagulation abnormalities, hypofibrinogenemia, and hypertriglyceridemia with onset in infancy. HLH is associated with hyperactivation and proliferation of T cells and macrophages, increased production of cytokines, reduced activity of cytotoxic T cells, and natural killer (NK) cells that accumulate at the maternal-fetal interface during pregnancy.

In the PL-099 family, a homozygous frameshift variant in the POC was inherited from both heterozygous parents (Figure S3-20). Previous studies have documented multiple instances of a neonatal death in babies with perforin deficiency.^83,84^

**Pathogenic variants in the *PTPN14* gene (Family PL-097)**

*PTPN14* (protein tyrosine phosphatase non-receptor type 14; OMIM*603155) is a member of the protein tyrosine phosphatase (PTP) family genes, which are known to regulate a variety of cellular processes, including lymphangiogenesis, cell growth, migration, and adhesion. *PTPN14* also modulates *TGF-beta* gene expression, promoting epithelial-mesenchymal transition. During prenatal stages of human development, *PTPN14* is expressed in a variety of tissues including stomach, spleen, kidney, lung and placenta (Supplemental Figures S5, S6, S7). **O**nly ∼60% of Ptpn14^-/-^ mice survive to birth, underlining the importance of *PTPN14* for prenatal development.^85^

In the PL-097 family, two heterozygous variants, inherited from both parents, were identified in the POC (Supplemental Figures S3-21). Biallelic pathogenic variants in the *PTPN14* gene have been reported in patients with intrauterine growth restriction, choanal atresia, lymphedema, dysmorphic facial features, and congenital heart defects, suggesting that *PTPN14* pathogenic variants may cause embryonic or neonatal death in humans**.^86^**

**Biallelic variants in the *RNF213* as a possible cause of pregnancy loss (Family PL-030)**

The ring finger protein 213 gene (*RNF213*; OMIM*613768) encodes Mysterin, a protein that contains the AAA+ ATPase and RING E3 domains. Previous studies have shown that RNF213 controls a wide range of biological processes including angiogenesis, vascular remodeling, lipid metabolism, immune responses, protein degradation, transcription, and DNA.^87-93^ RNF213 is widely expressed, particularly at the early stages of embryogenesis (Supplemental Figures S5, S6, S7). Heterozygous variants in the *RNF213* gene are associated with Moyamoya disease, characterized by progressive cerebrovascular lesions, coronary artery disease and pulmonary artery stenosis. Affected children experience ischemic attacks that can lead to cerebral infarction, intracranial hemorrhage, hemiplegia, and epileptic seizures. Prenatal and neonatal presentations may include mitochondrial disorder, acute liver failure, cardiomyopathy and death in infancy.^94^

In the PL-030 family, we found biallelic missense variants in the POC inherited from both parents (Figure S3-22). Homozygous and compound heterozygous variants in *RNF213* are extremely rare and generally seen in neonates with severe presentations. Here we report the first case of a fetal demise in association with biallelic variants in the *RNF213* gene.

**Findings in the *SCN5A* gene (Family PL-129)**

The *SCN5A* (sodium voltage-gated channel alpha subunit 5; OMIM*600163) gene encodes the alpha subunit of the cardiac sodium channel Nav1.5, which plays a critical role in the regulation of cardiac conduction. Heterozygous pathogenic variants in the *SCN5A* gene have been shown to be associated with a spectrum of cardiac conditions, including long QT syndrome, Brugada syndrome, atrial and ventricular fibrillation, and dilated cardiomyopathy. Affected individuals, including infants and fetuses, are predisposed to sudden death.

In the PL-129 family (Figure S3-23), we found a missense, likely pathogenic variant in the POC inherited from an asymptomatic father. A high rate of intrauterine fetal loss, stillbirth and neonatal death has been reported in individuals with *de novo* and familial variants in the *SCN5A* gene.^41,95-99^

### **Biallelic variants in the *SLC12A3* gene (Family PL-077)**

*SLC12A3* (solute carrier family 12 member 3; OMIM*600968) encodes a renal sodium-chloride cotransporter responsible for electrolyte homeostasis. In preimplantation embryos, *SLC12A3* is expressed in the trophectoderm and inner cell mass, whereas at the fetal stage of development it is almost exclusively expressed in the kidney (Figure S5). SLC12A3 plays the fundamental role in sodium, magnesium and calcium reabsorption and in the maintenance of extracellular fluid volume. Biallelic pathogenic variants in *SLC12A3* have been found in patients with Gitelman syndrome, an autosomal recessive salt-wasting disorder, characterized by hypokalemic alkalosis, hypomagnesemia, low urinary calcium, and occasionally more severe presentations such as growth retardation, rhabdomyolysis, seizures, and cardiac arrhythmia. Prenatal manifestations of Gitelman syndrome are mainly unknown, but may include polyhydramnios, cystic kidneys, and fetal demise. In the PL-077 family (Figure S3-24), we found a *de novo* and a maternal missense variant in the POC.

### ***SMN1* and *SMN2* alterations in Family PL-098**

Homozygous loss of the *SMN1* (survival of motor neuron 1, telomeric; OMIM*600354) gene on chromosome 5q13 is the cause of spinal muscular atrophy (SMA). *SMN2* (survival of motor neuron 2, centromeric OMIM*601627) is a gene-modifier of SMA phenotype. SMA is an autosomal recessive condition and is a leading cause of infantile and pediatric death.^100,101^ Prenatal findings in PL-098C suggested a severe presentation of SMA, with fetal growth restriction, cystic hygroma, and polyhydramnios (Supplementary Table S1). The SMN1 protein is a member of the SMN complex, which consists of nine proteins responsible for the assembly of small nuclear ribonucleoproteins essential for pre-mRNA splicing. In addition, SMN1 a multifunction protein that plays a role in translational control, splicing, mRNA transport, ribosome biology, signal transduction crucial cellular homeostasis and cell survival. The *SMN1*x0, *SMN2*x1 genotype detected in pregnancy loss samples in this family is associated with the most severe SMA, type-0, characterized by prenatal onset (Figure S3-25). Pregnancy loss has been reported previously in SMA patients in humans.^102,103^ Mouse knockout models for *SMN1* were not viable and showed preimplantation lethality. Biallelic loss of *SMN1* and reduced expression of *SMN2* is a novel cause for early human lethality. The incidence rate of SMA in live births (1 in 20,000) is lower than expected (1 in 2,500), based on the carrier frequency of *SMN1* (1 in 50), suggestive of embryonic lethality.

**Findings in the *TLR8* gene (Family PL-029)**

*TLR8* encodes an endosomal toll like receptor 8 (OMIM *300366), pathogenic variants in which are associated with X-linked immunodeficiency with autoinflammation (OMIM# 301078). Germline gain-of-function variants have been described in children affected by recurrent infections, neutropenia, bone marrow failure, and lymphoproliferation, leading to severe immune dysregulation, autoinflammation and childhood lethality (Figure 3). Importantly, the majority of affected male patients had germline mosaicism for the *TLR8* missense variant present in 8–26% of cells among the peripheral blood and multiple somatic tissues.^104^ Non-mosaic germline *TLR8* variants have been identified in two affected boys,^105^ and a female patient^106^ with a neonatal onset of anemia, reticulocytopenia, neutropenia, lymphocytopenia and hepatosplenomegaly. These observations indicate a dominant gain-of-function mechanism of TLR8-related immune dysregulation, consistent with an X-linked dominant disorder. As with multiple other X-linked dominant conditions, male individuals are severely affected, whereas carrier females may be asymptomatic or have mild or late onset symptoms, depending on the X chromosome inactivation pattern.

In the PL-029 family, we found a missense variant in the POC with XY sex chromosome complement. The variant was inherited from the mother, who has a long history of infertility (Figure S3-26). In all previously described patients, pathogenic variants were located in the leucine-rich repeats (LRR), which are responsible for recognition of microorganism ligands specific pathogens. In contrast, the variant detected in this family is present in a conserved cytoplasmic Toll/interleukin-1 receptor (TIR) domain, which plays a critical role in the binding of proteins involved in signaling leading to NF-kappa-B activation, secretion of cytokines and the inflammatory response. Interestingly, the *IKBKG* (Inhibitor of NF-kappa-B kinase regulatory subunit gamma) gene is another member of the Toll-like Receptor Signaling Pathway, and pathogenic *IKBKG* variants cause the X-linked systemic autoinflammatory disease and lethality in males. Transcriptomic analysis showed very low expression of *TLR8* during the preimplantation and embryonic stages of normal fetal development. We hypothesize that missense variants in the cytoplasmic domain of *TLR8* could cause activation of a cellular response and autoimmune inflammation, even when no ligand is attached.

### **Findings in the *TNNI3* gene (Family PL-083)**

*TNNI3* (troponin I3; OMIM*191044) encodes cardiac troponin, heterozygous pathogenic variants in which are the cause of hypertrophic, dilated and restrictive cardiomyopathy. In PL-083 family (Figure S3-27), a missense variant in the POC was inherited from the father who had heart transplant due to a childhood onset of restrictive cardiomyopathy. The *TNNI3* variants has been reported in individuals with predominantly childhood onset cardiomyopathy, cardiac insufficiency, pulmonary hypertension, and mitral and tricuspid insufficiency, and neonatal death.^107^

### **Homozygous variant in the *VPS35L* gene (Family PL-059)**

The *VPS35L* (Endosomal protein-sorting factor-like; OMIM*618981) gene is a subunit of the Commander complex, which is required for trafficking and recycling of hundreds of proteins through the endosomal network.^108^ During early embryonic development, the *VPS35L* gene is strongly expressed in many tissues (Supplemental Figures S5), including placenta. Biallelic variants in the *VPS35L* gene are associated with Ritscher-Schinzel syndrome, also known as 3C syndrome, a multisystemic disorder characterized by craniofacial dysmorphic features, cerebellar hypoplasia, and cardiovascular abnormalities. Vps35l^−/−^ mice demonstrated embryonic lethality at an early stage.^109^ Ritscher-Schinzel syndrome due to variants in the *VPS35L* gene is associated with diverse and more severe phenotypes complicated by hypogammaglobulinaemia and intestinal lymphangiectasia.^110^

In the PL-059 family, we found a homozygous missense variant in the POC inherited from unrelated parents (Figure S3-28). This variant occurred within a highly conserved part of the protein which is important for the Commander complex assembly.^111^ VPS35L is necessary for autophagic function and essential for embryo survival.

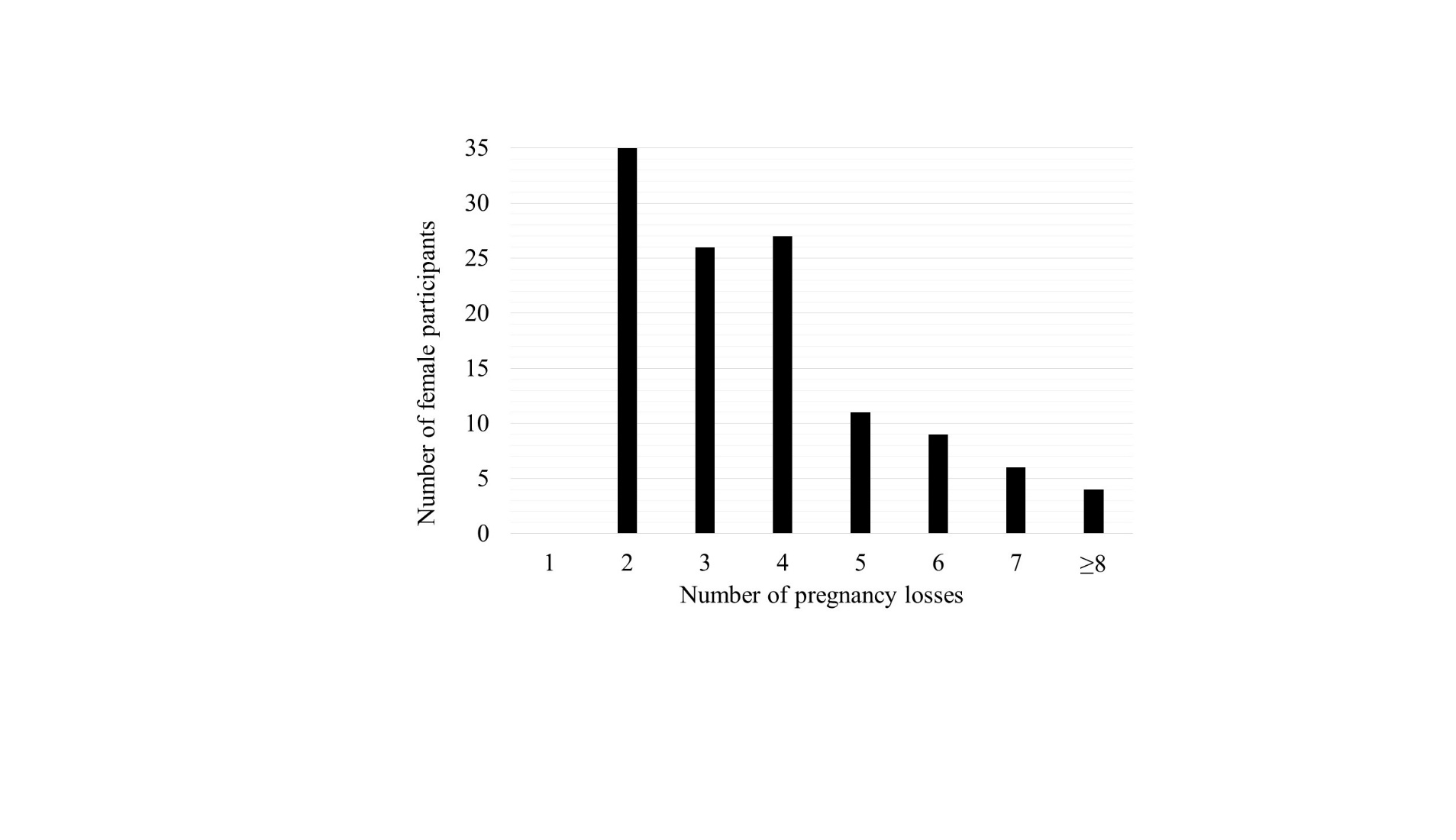

**Figure S1**. **Number of miscarriages in the study cohort**.

In 74.6% of families (88/118) up to 4 losses were documented. Twenty-six couples (21.7%) experienced 5-7 pregnancy losses. Four couples (3.4%) had 8-12 (≥ 8) miscarriages.

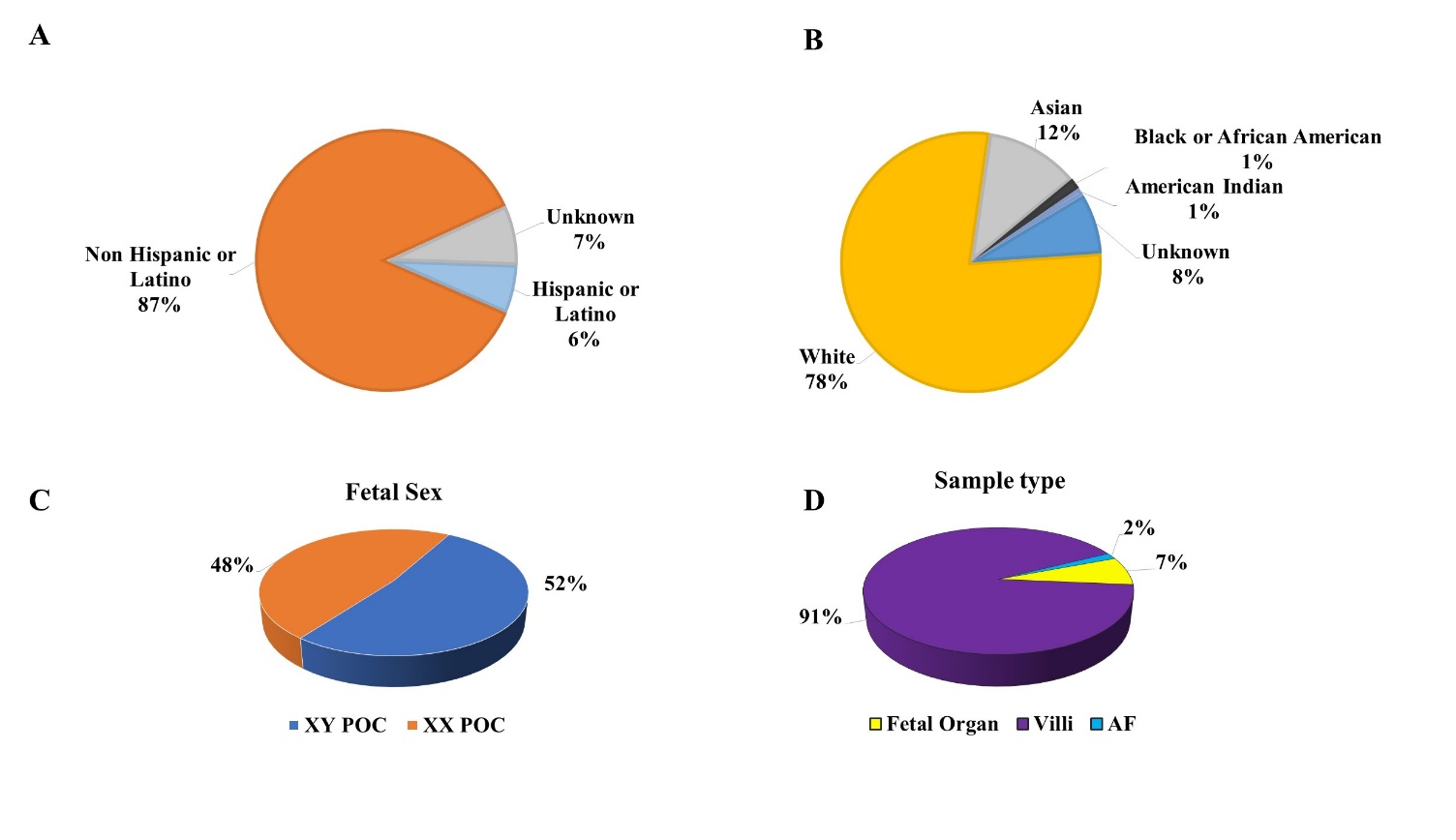

**Figure S2. Demographic and Sample Characteristics**.

**(A) Ethnicity distribution of parental samples. (B) Race distribution of parental samples,** demonstrating that the predominant racial group was White (78%), followed by Asian (12%). The minority of individuals identified as Black or African American (1%) or American Indian (1%). Race in 8% of the participants was categorized as Unknown. **(C) Fetal sex distribution in pregnancy loss samples:** 52% were identified as euploid with XY sex chromosome complement, and 48% had a 46,XX karyotype. **(D)** Tissue source **of pregnancy loss samples**. The majority of collected samples originated from chorionic villi, with a smaller proportion derived from amniotic fluid (AF) and fetal organ tissue (lung, kidney, liver, muscle).

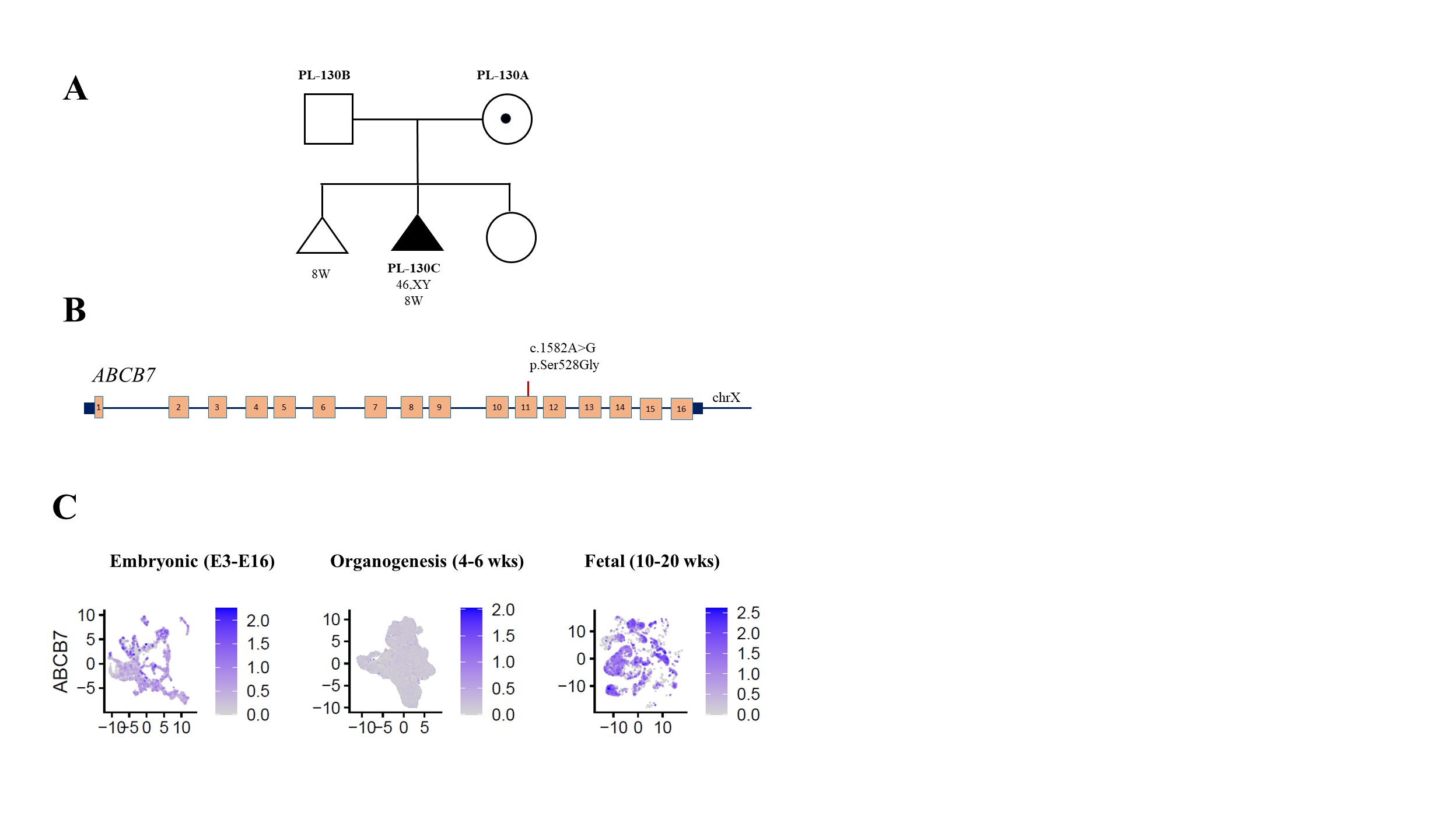

**Figure S3-1.** **Findings in the *ABCB7* gene**.

(**A**) Pedigree of the PL-130 family. Tested POC sample (PL-130C) is shown as a black triangle. (**B**) Schematic representation of the *ABCB7* gene located on chromosome X. Coding exons are denoted by orange blocks. Black boxes indicate non-coding exons. In pregnancy loss sample, a missense hemizygous variant, inherited from the mother, was found in exon 11. (**C**) Log-normalized expression of *ABCB7* gene in three stages: Embryonic E3-E16 (Embryonic day 3 to 16), organogenesis (4-6 weeks of gestation), and fetal (10-20 weeks of gestation) development.

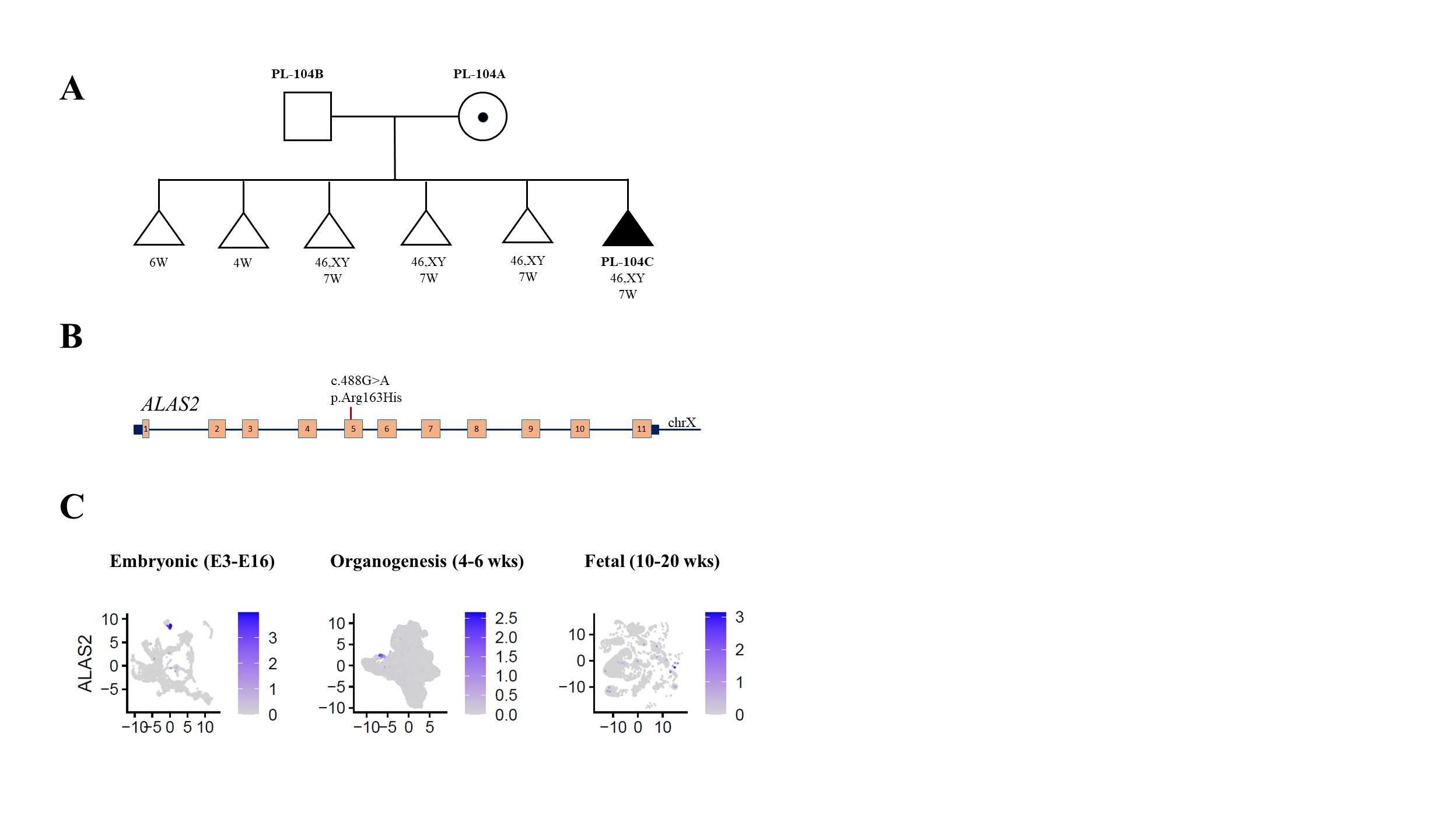

**Figure S3-2. Findings in the *ALAS2* gene.**

(**A**) Pedigree of the PL-104 family. (**B**) Schematic representation of the *ALAS2* gene located on chromosome X. In pregnancy loss sample PL-104C, a maternally inherited missense hemizygous variant was found in exon 5. (**C**) Log-normalized expression of *ALAS2* gene in three stages: Embryonic E3-E16 (Embryonic day 3 to 16), organogenesis (4-6 weeks of gestation), and fetal (10-20 weeks of gestation) development.

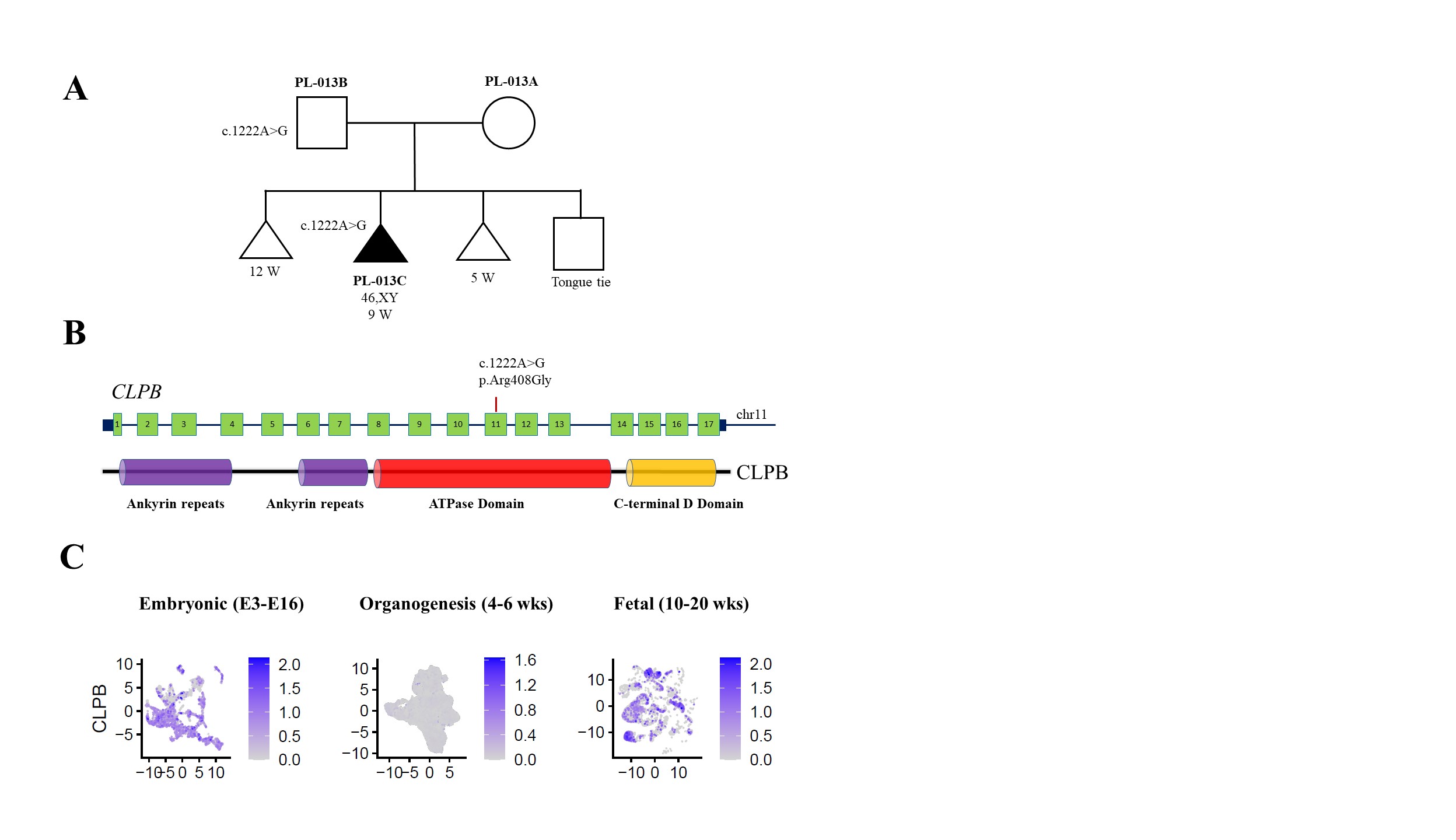

**Figure S3-3. Findings in the *CLPB* gene**.

(**A**) Pedigree of the PL-013 family. (**B**) Schematic representation of the *CLPB* gene located on chromosome 11. Below, the CLPB protein domains. In pregnancy loss sample PL-013C, a paternally inherited missense heterozygous variant was found in exon 11, encoding the ATPase domain. (**C**) Log-normalized expression of *CLPB* gene in three stages: Embryonic E3-E16 (Embryonic day 3 to 16), organogenesis (4-6 weeks of gestation), and fetal (10-20 weeks of gestation) development.

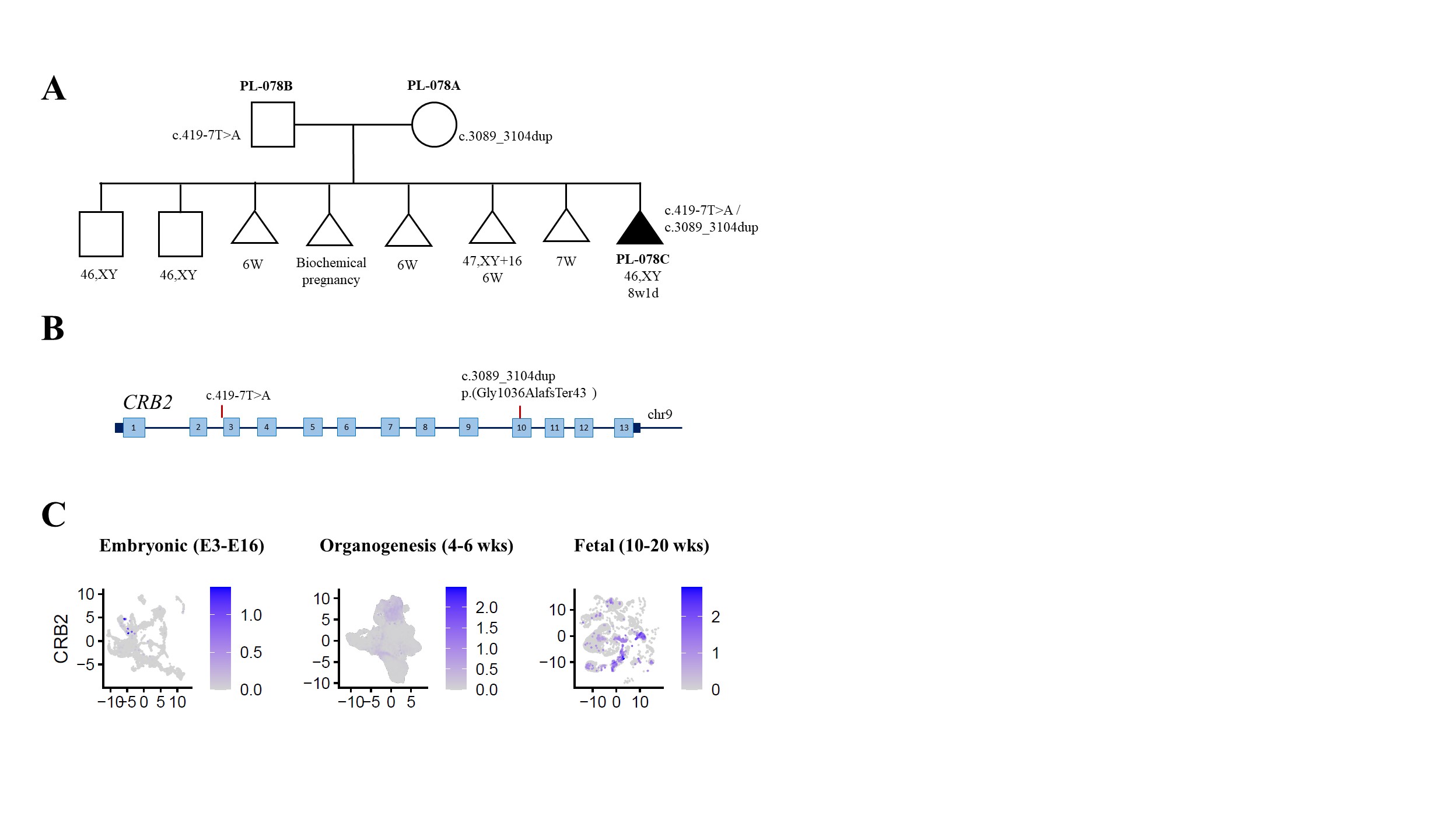

**Figure S3-4. Findings in the *CRB2* gene.**

(**A**) Pedigree of the PL-078 family. (**B**) Schematic representation of the *CRB2* gene located on chromosome 9. In pregnancy loss sample PL-078C, an inherited compound heterozygous variant including a maternally inherited heterozygous frameshift variant in exon 10 and a paternally inherited splice region heterozygous variant in intron 2 was found. (**C**) Log-normalized expression of *CRB2* gene in three stages: Embryonic E3-E16 (Embryonic day 3 to 16), organogenesis (4-6 weeks of gestation), and fetal (10-20 weeks of gestation) development.

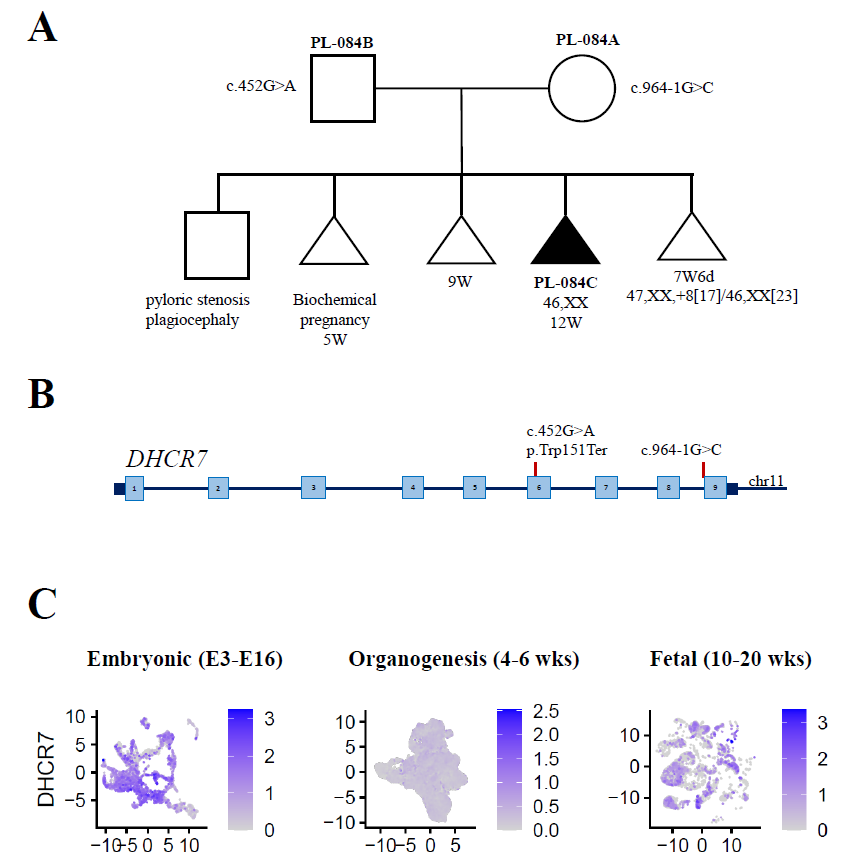

**Figure S3-5. Findings in the *DHCR* gene.**

(**A**) Pedigree of the PL-084 family. (**B**) Schematic representation of the *DHCR7* gene located on chromosome 8. In pregnancy loss sample PL-084C, biallelic variants were found in exon 6 and exon 9. (**C**) Log-normalized expression of *DHCR7* gene in three stages: Embryonic E3-E16 (Embryonic day 3 to 16), organogenesis (4-6 weeks of gestation), and fetal (10-20 weeks of gestation) development.

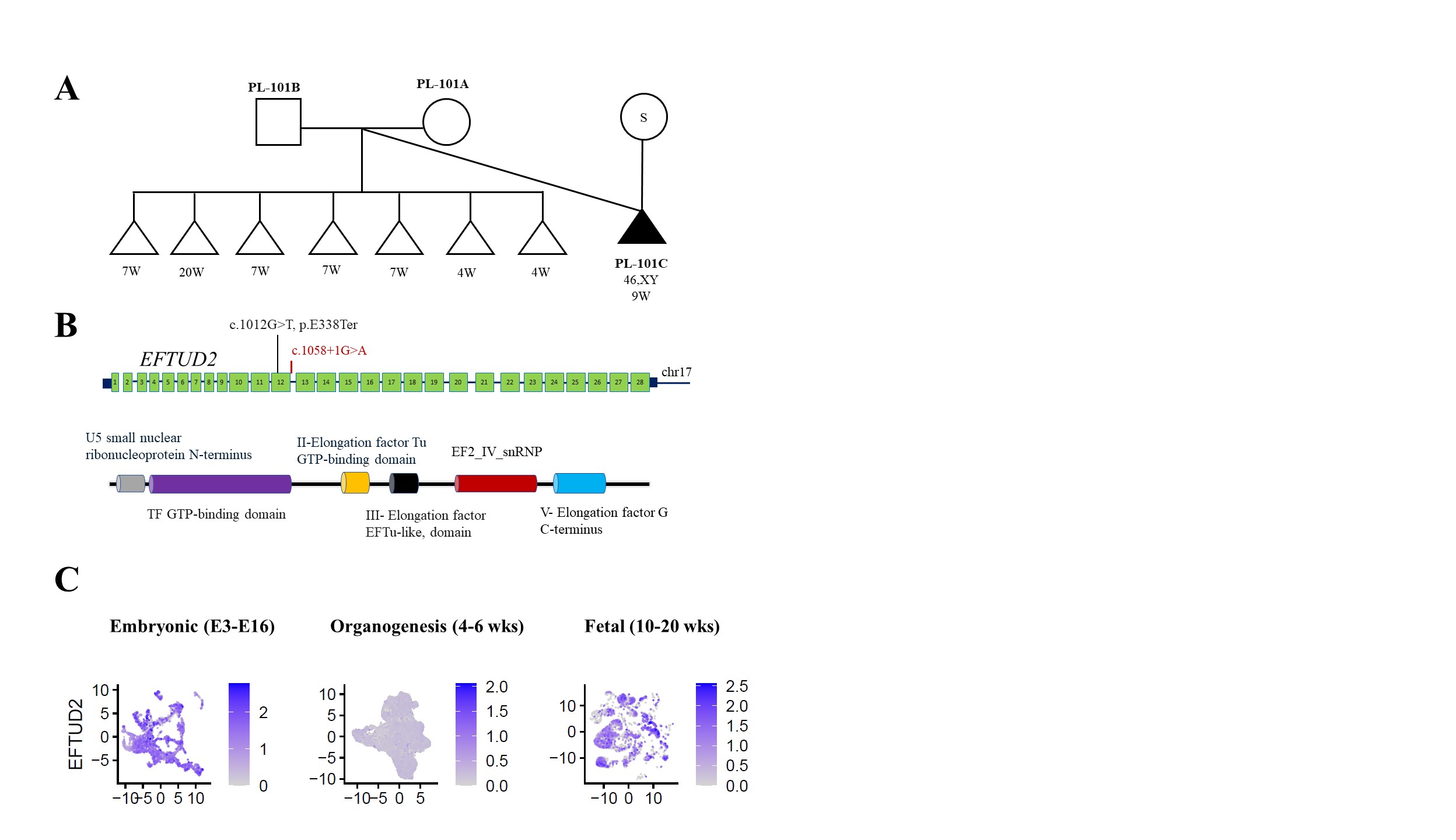

**Figure S3-6. Findings in the *EFTUD2* gene.**

(**A**) Pedigree of the PL-101 family. A surrogate carrier represented by a “S” in a circle.

(**B**) Schematic representation of the *EFTUD2* gene located on chromosome 17. Below, the EFTUD2 protein domains. In red, a variant detected in pregnancy loss sample PL-101C. In black, variant in a pregnancy loss case reported previously.^40^ Both variants are located in the TF GTP-binding domain. (**C**) Log-normalized expression of *EFTUD2* gene in three stages: Embryonic E3-E16 (Embryonic day 3 to 16), organogenesis (4-6 weeks of gestation), and fetal (10-20 weeks of gestation) development.

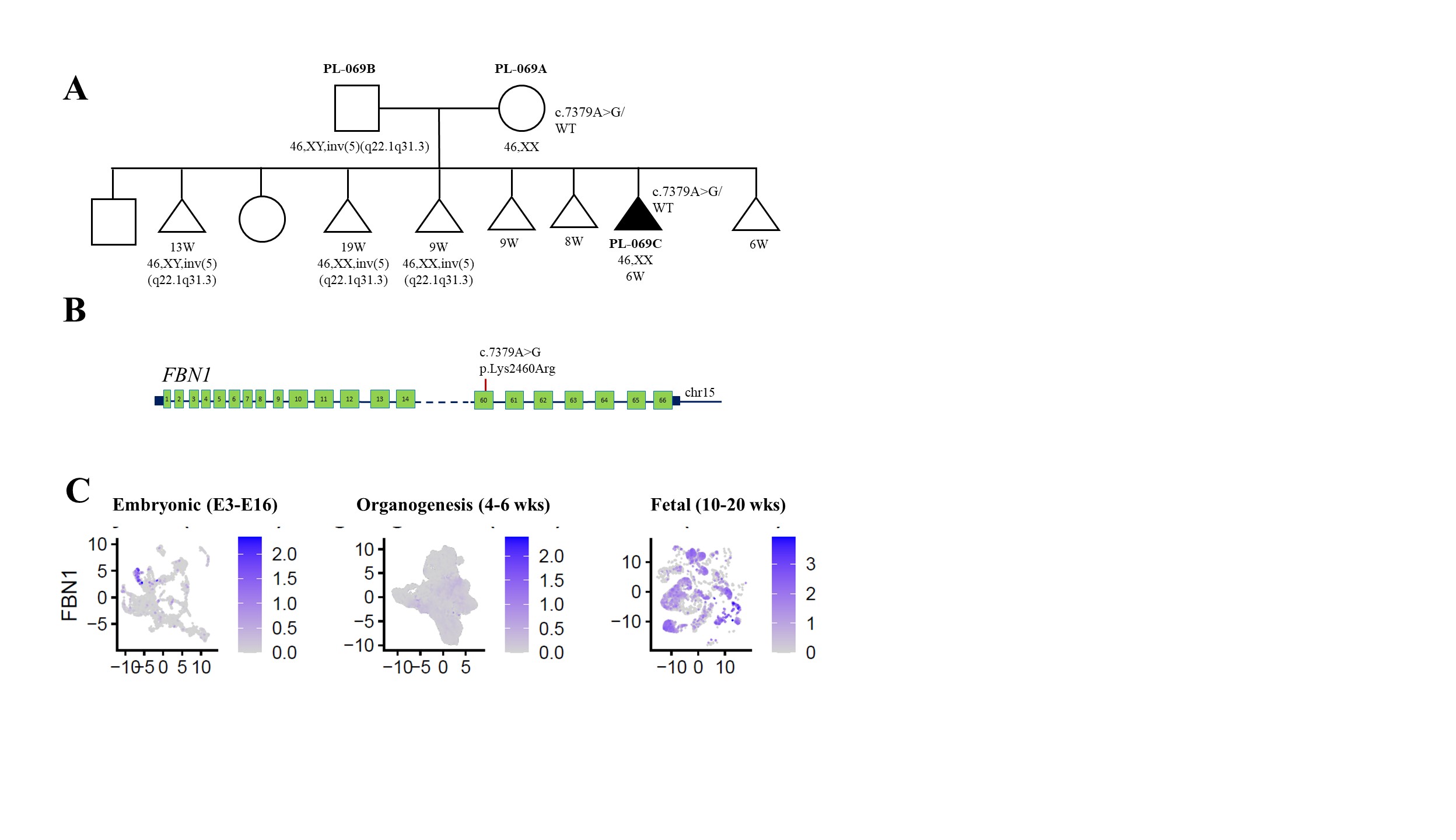

**Figure S3-7. Findings in the *FBN1* gene.**

(**A**) Pedigree of the PL-069 family. (**B**) Schematic representation of the *FBN1* gene located on chromosome 15. In pregnancy loss sample PL-069C, a maternally inherited missense heterozygous variant was found in exon 60. (**C**) Log-normalized expression of *FBN1* gene in three stages: Embryonic E3-E16 (Embryonic day 3 to 16), organogenesis (4-6 weeks of gestation), and fetal (10-20 weeks of gestation) development.

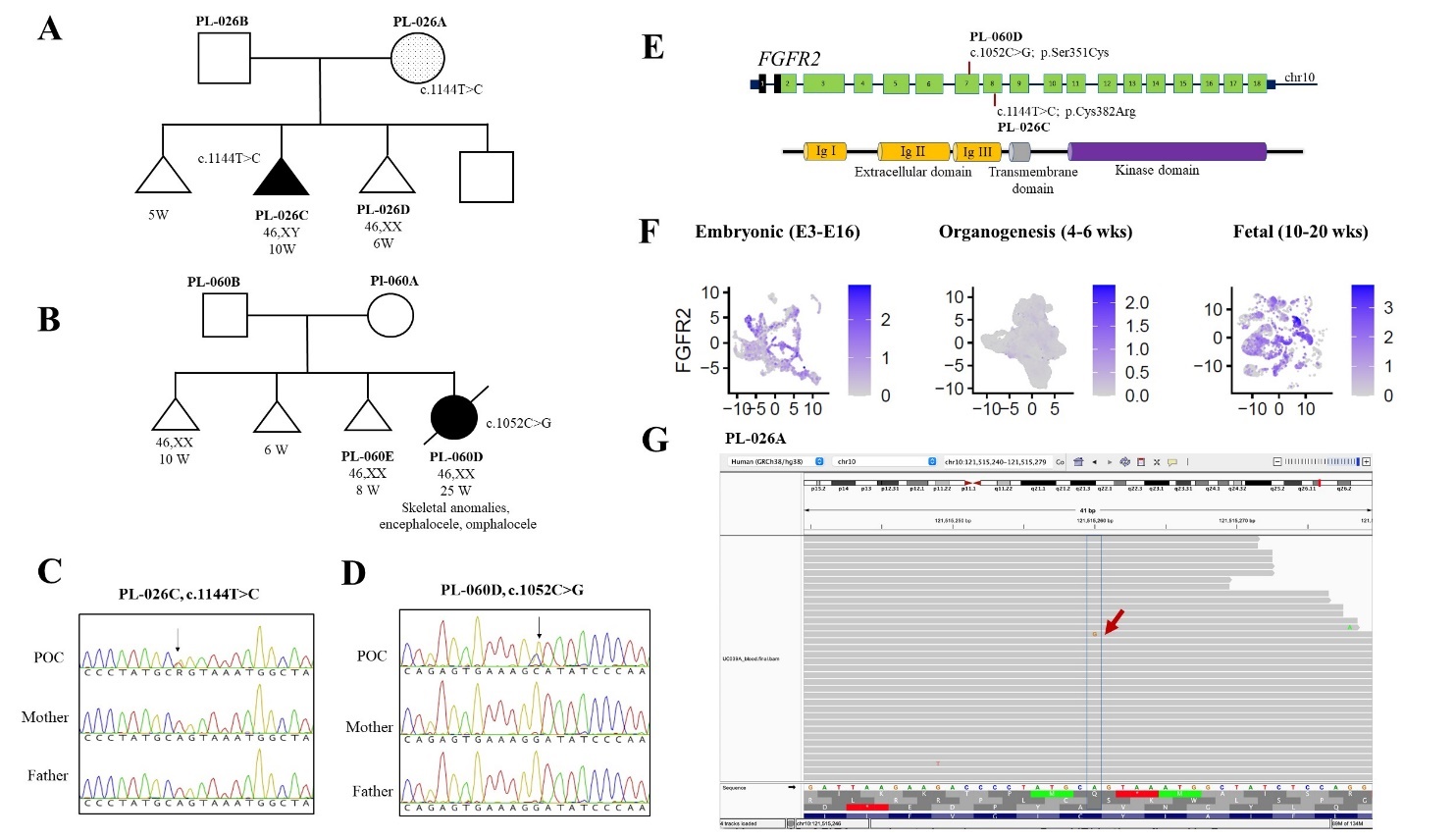

**Figure S3-8. Findings in the *FGFR2* gene.**

(**A**) Pedigree of the PL-026 family and (**B**) PL-060 family. Fetuses with pathogenic variants in the *FGFR2* gene are indicated by filled triangle and circle. Dotted shade indicates a germline mosaicism in the parent. (**C, D**) In pregnancy loss samples, heterozygous variants were confirmed by Sanger sequencing. (**E**) Schematic representation of the *FGFR2* gene located on chromosome 10 and variants found in exon 7 and 8 in affected POC samples. Below, the FGFR2 protein structure, showing extracellular region, composed of three immunoglobulin-like domains (Ig I, Ig II, and Ig III); transmembrane domain; and cytoplasmic tyrosine kinase domain. (**F**) Log-normalized expression of *FGFR2* gene in three stages: Embryonic E3-E16 (Embryonic day 3 to 16), organogenesis (4-6 weeks of gestation), and fetal (10-20 weeks of gestation) development. (**G**) Visualization of sequencing reads on Integrative Genome Viewer showing the mosaic variant (red arrow) in a blood sample from asymptomatic mother PL-026A.

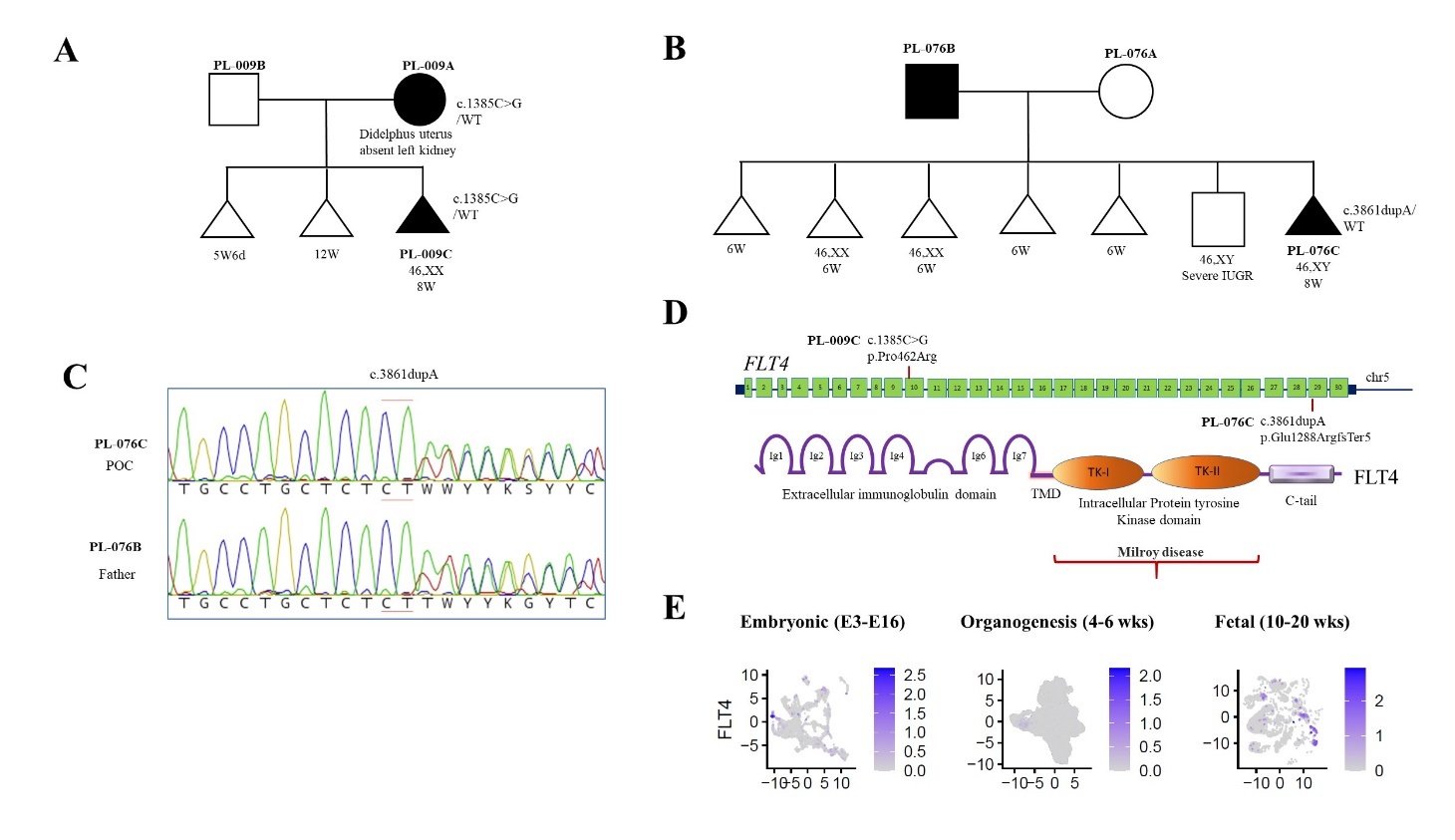

**Figure S3-9. Findings in the *FLT4* gene.**

(**A**) Pedigree of the PL-009 family and (**B**) PL-076 family, confirmed by Sanger sequencing (**C**). (**D**) Schematic representation of the *FLT4* gene located on chromosome 5 and variants found in exon 10 and 29 in affected POC. Below, the FLT4 protein structure, comprising the extracellular domain, organized into immunoglobulin (Ig)-like folds; TMD- transmembrane domain; split tyrosine-kinase domain, interrupted by a 70-amino-acid kinase insert, and a C-terminal tail. Variants in the Kinase domain are generally associated with Milroy disease, while variants in other domains are seen in patients with congenital heart defects. (**E**) Log-normalized expression of *FLT4* gene in three stages: Embryonic E3-E16 (Embryonic day 3 to 16), organogenesis (4-6 weeks of gestation), and fetal (10-20 weeks of gestation) development.

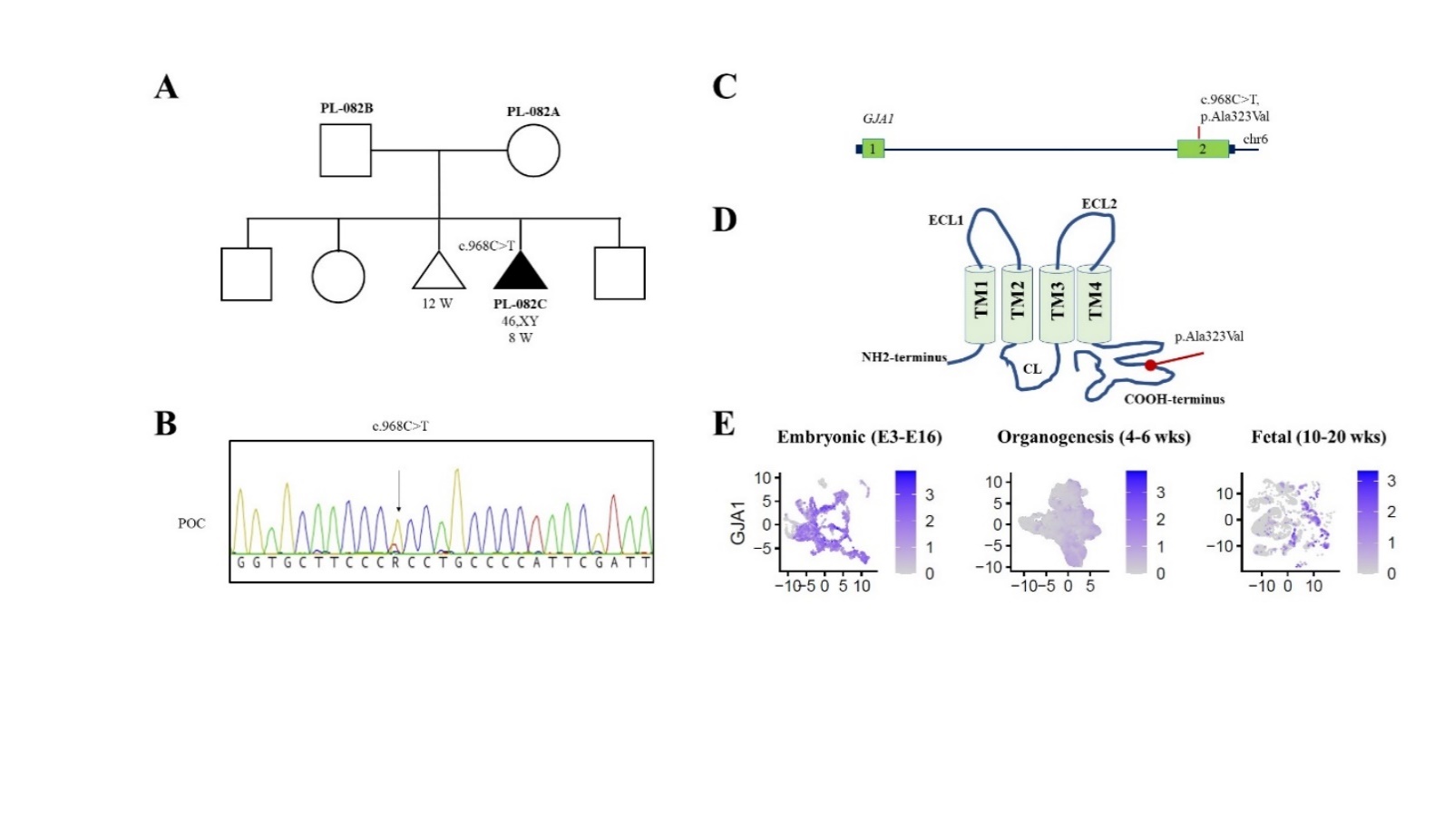

**Figure S3-10. Findings in the *GJA1* gene.**

(**A**) Pedigree of the PL-082 family. In pregnancy loss sample PL-082C, a *de novo* missense heterozygous variant was found by genome sequencing and (**B**) confirmed by Sanger sequencing. (**C**) Schematic representation of the *GJA1* gene located on chromosome 6 and a variant found in exon 2. (**D**) GJA1 protein is composed of four α-helical transmembrane helix domains (TM1-4), two extracellular loops (ECL1-2), a cytoplasmic loop (CL), the cytoplasmic the amino (NH2) and carboxyl (COOH) termini. Alanine to valine substitution is located within the COOH tail (red dot). (**E**) Log-normalized expression of *GJA1* gene in three stages: Embryonic E3-E16 (Embryonic day 3 to 16), organogenesis (4-6 weeks of gestation), and fetal (10-20 weeks of gestation) development.

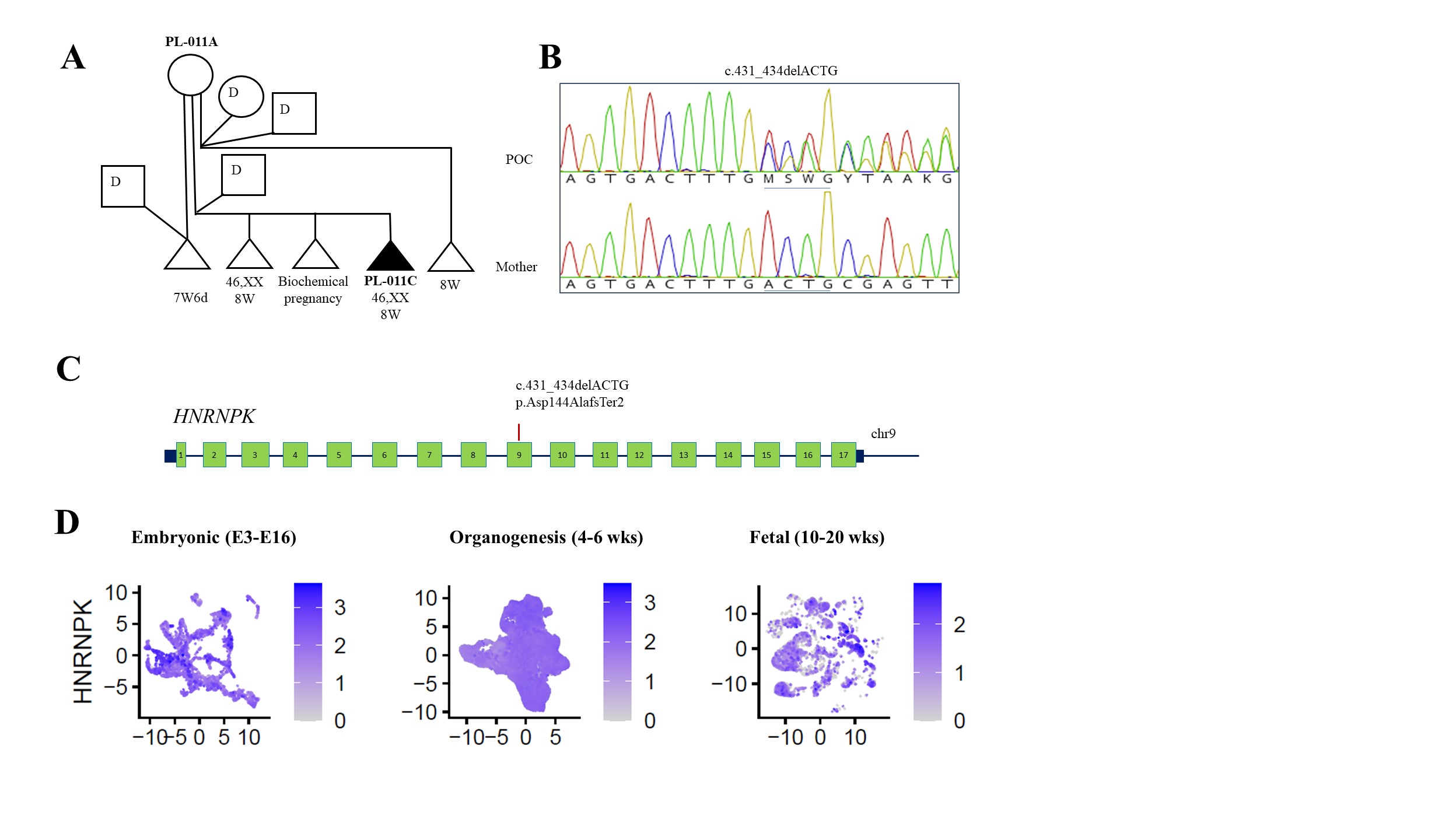

**Figure S3-11. Findings in the *HNRNPK* gene.**

(**A**) Pedigree of the PL-011 family. A sperm/egg donor is indicated by “D”. A total three sperm donors and an egg donor were used for 5 pregnancies. (**B**) In pregnancy loss sample PL-011C, a frameshift heterozygous variant was identified in confirmed by Sanger sequencing. (**C**) Schematic representation of the *HNRNPK* gene located on chromosome 9 and a frameshift variant found in exon 9.

(**D**) Log-normalized expression of *HNRNPK* gene in three stages: Embryonic E3-E16 (Embryonic day 3 to 16), organogenesis (4-6 weeks of gestation), and fetal (10-20 weeks of gestation) development.

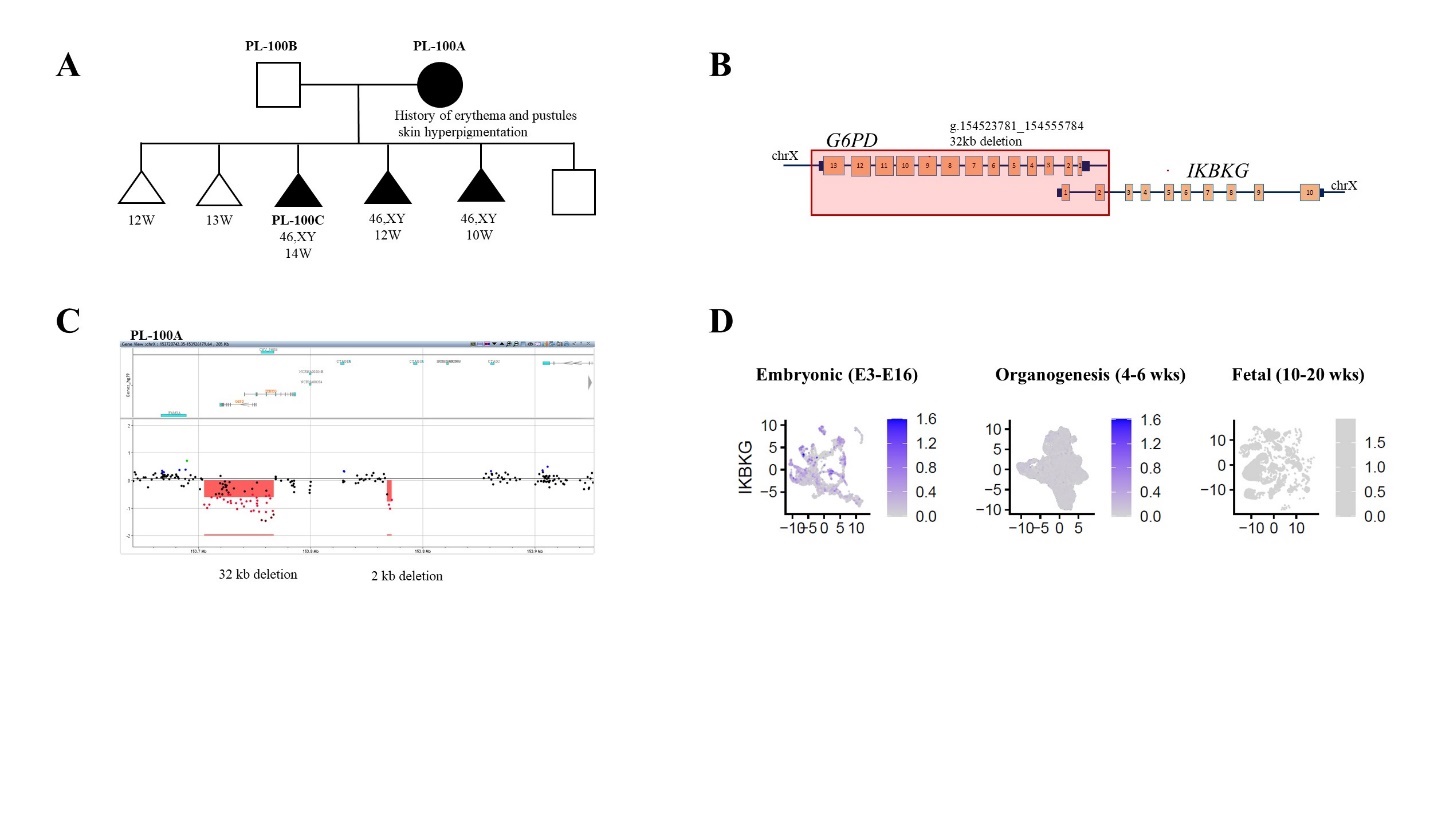

**Figure S3-12. Findings in the *IKBKG* gene.**

(**A**) Pedigree of the PL-100 family. (**B**) Schematic representation of the *IKBKG* gene located on the X chromosome. In pregnancy loss sample PL-100C, a 32 kb maternally inherited deletion (pink shaded rectangle) was found on chromosome X. This deletion encompasses exons 1 and 2 of the *IKBKG* gene and the entire *G6PD* gene. (**C**) The Xq28 deletion in the mother detected by custom X-chromosome high resolution microarray. (**D**) Log-normalized expression of *IKBKG* gene in three stages: Embryonic E3-E16 (Embryonic day 3 to 16), organogenesis (4-6 weeks of gestation), and fetal (10-20 weeks of gestation) development.

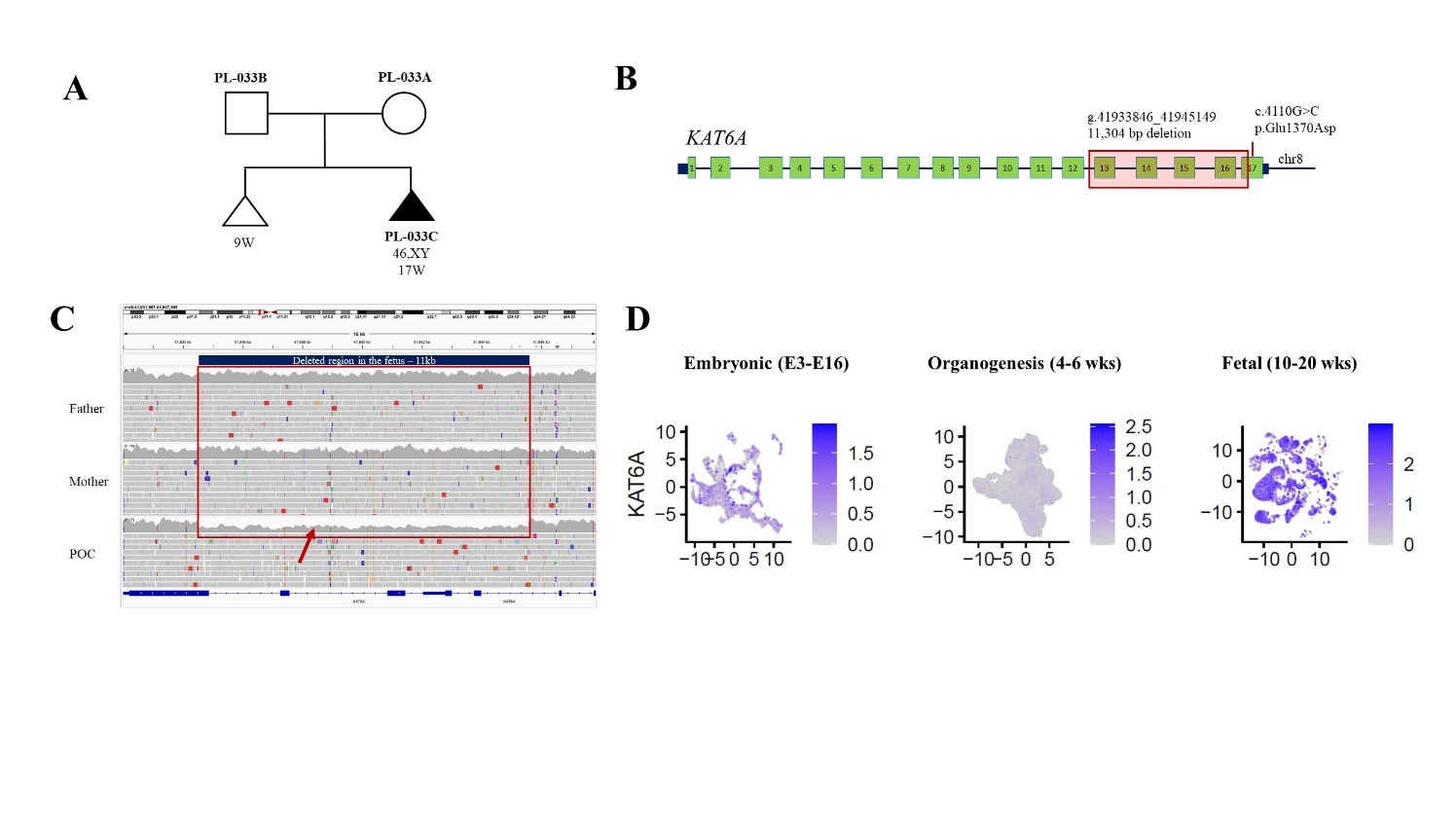

**Figure S3-13. Findings in the *KAT6A* gene.**

(**A**) Pedigree of the PL-033 family. (**B**) Schematic representation of the *KAT6A* gene located on chromosome 8. In pregnancy loss sample PL-033C, a *de novo* 11 kb deletion comprising exons 13-17 is indicated by a pink shaded area. (**C**) IGV display of a detailed coverage track from paternal, maternal and POC BAM files showing a reduced coverage in the POC at the deleted region (red arrow). SNP analysis of the affected segment indicates that the deletion arose *de novo* on a paternal chromosome. (**D**) Localization of *KAT6A* deletion has been shown in IGV (E) Log-normalized expression of *KAT6A* gene in three stages: Embryonic E3-E16 (Embryonic day 3 to 16), organogenesis (4-6 weeks of gestation), and fetal (10-20 weeks of gestation) development.

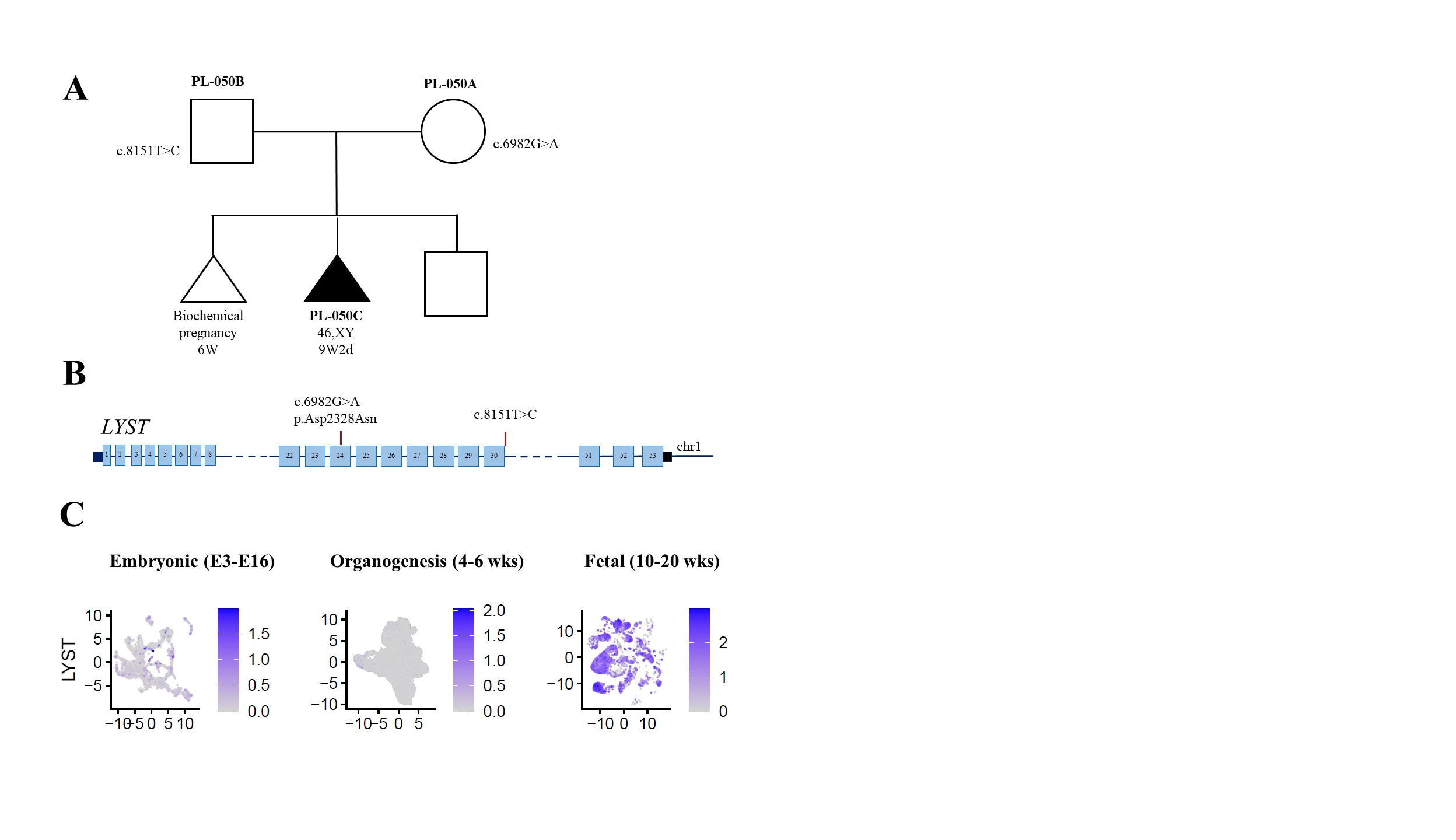

**Figure S3-14. Findings in the *LYST* gene.**

(**A**) Pedigree of the PL-050 family. (**B**) Schematic representation of the *LYST* gene located on chromosome 1. In pregnancy loss sample PL-050C, two variants were detected including a missense variant in exon 24 and a splice region variant in intron 30. (**C**) Log-normalized expression of *LYST* gene in three stages: Embryonic E3-E16 (Embryonic day 3 to 16), organogenesis (4-6 weeks of gestation), and fetal (10-20 weeks of gestation) development.

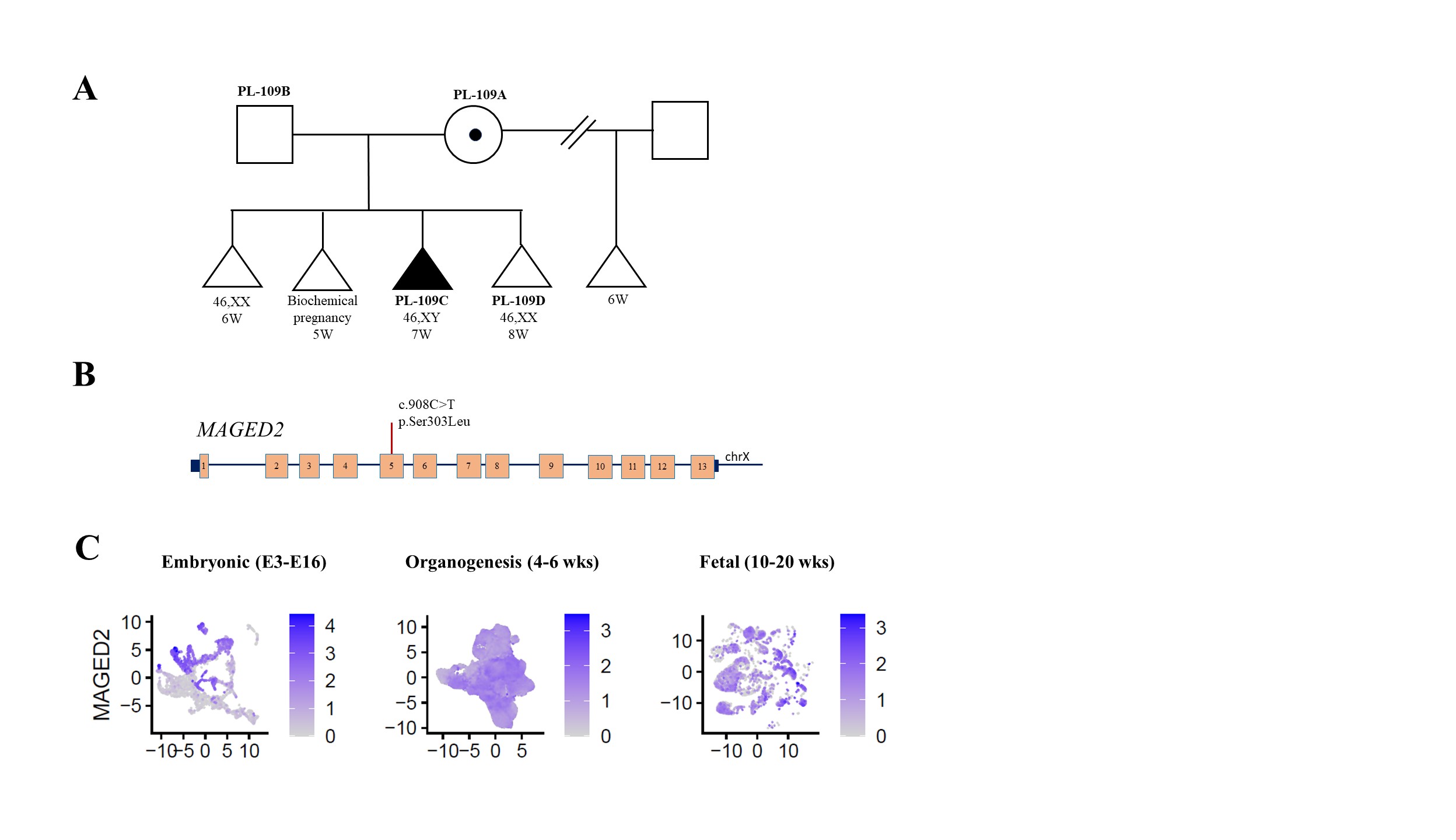

**Figure S3-15. Findings in the *MAGED2* gene.**

(**A**) Pedigree of the PL-109 family. (B) Schematic representation of the *MAGED2* gene located on the X chromosome. In pregnancy loss sample PL-109C, a maternally inherited missense heterozygous variant was found in exon 5. (D) Log-normalized expression of *MAGED2* gene in three stages: Embryonic E3-E16 (Embryonic day 3 to 16), organogenesis (4-6 weeks of gestation), and fetal (10-20 weeks of gestation) development.

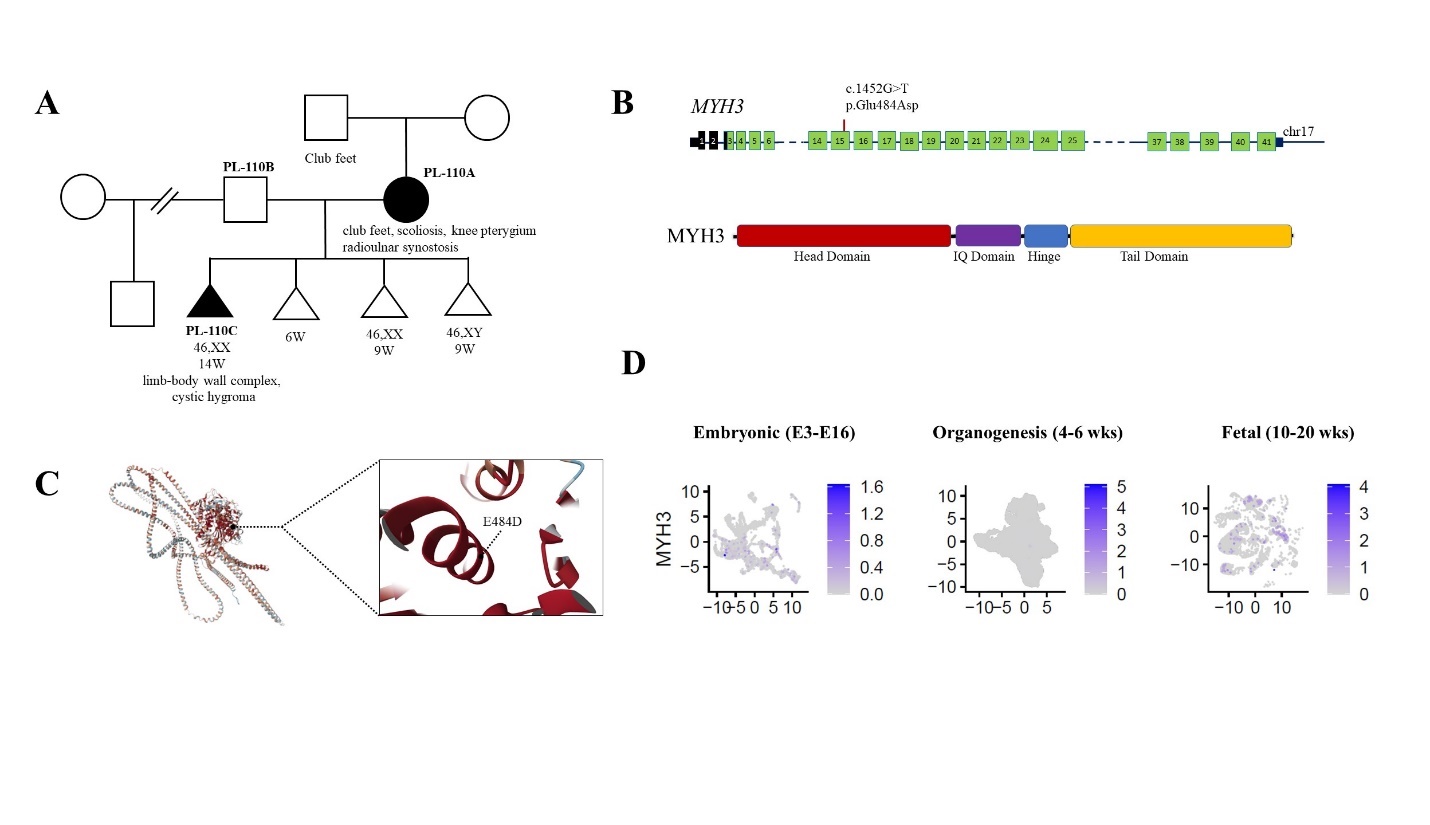

**Figure S3-16. Findings in the *MYH3* gene.**

(**A**) Pedigree of the PL-110 family. Note, the mother (PL-110A) presented with skeletal manifestations (club feet, scoliosis, unilateral knee pterygia) consistent with *MYH3*-associated distal arthrogryposis. Maternal father had club feet. (**B**) Schematic representation of the *MYH3* gene located on chromosome 17. Coding exons are denoted by green blocks. Black boxes indicate non-coding exons. In pregnancy loss sample PL-110C, a heterozygous missense variant was found in exon 15. Below is a structure of the MYH3 protein. This novel variant, located within the head domain of the myosin molecule, was inherited from the mother (**C**) AlphaFold structure of MYH3 with a magnified view of the pathogenic *MYH3* E484D variant identified in the family. (**D**) Log-normalized expression of *MYH3* gene in three stages: Embryonic E3-E16 (Embryonic day 3 to 16), organogenesis (4-6 weeks of gestation), and fetal (10-20 weeks of gestation) development.

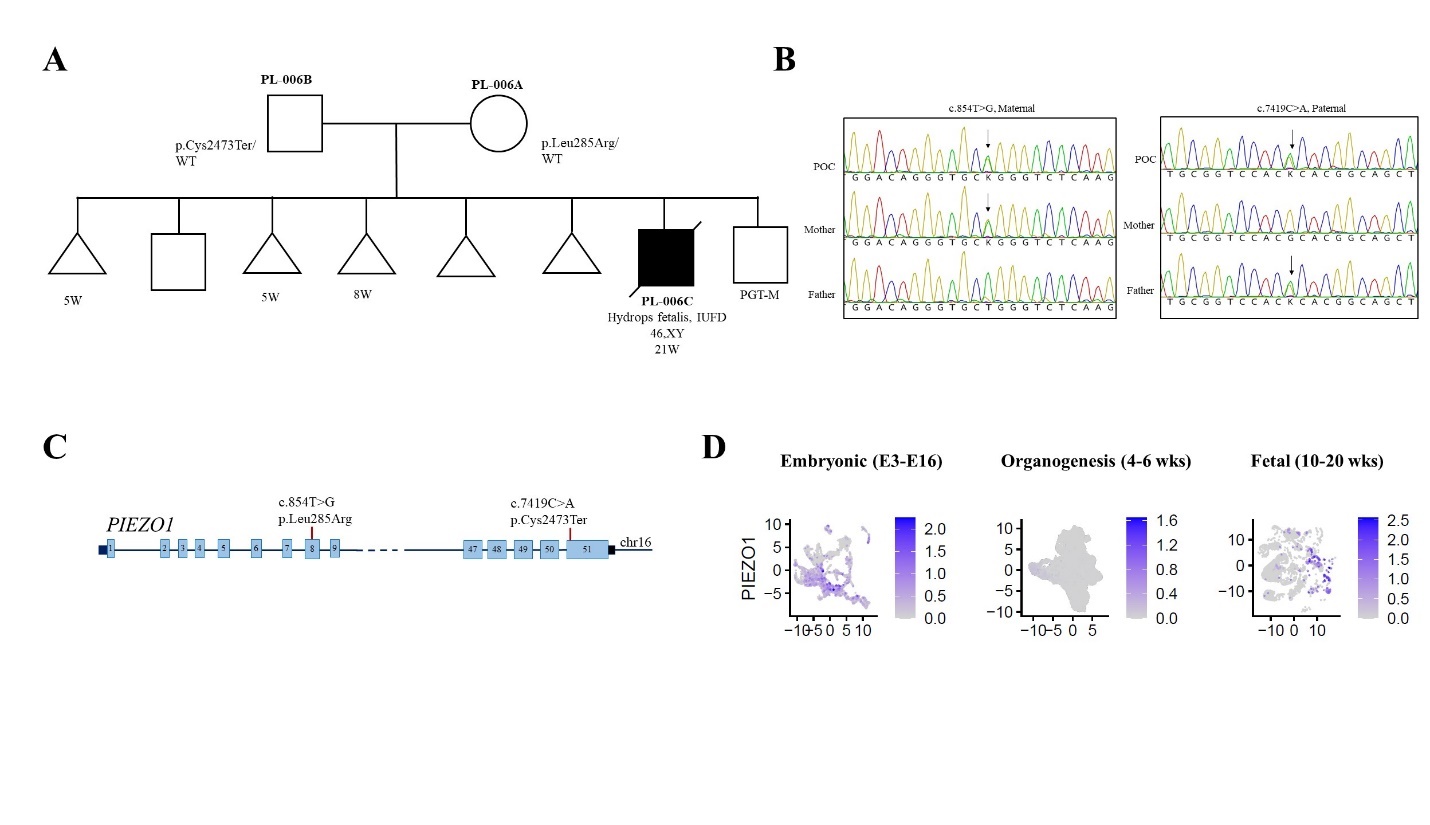

**Figure S3-17. Findings in the *PIEZO1* gene.**

(**A**) Pedigree of the PL-006 family. (**B**) In pregnancy loss sample PL-006C, biparental inheritance of *PIEZO1* variants was confirmed by Sanger sequencing. (**C**) Schematic representation of the *PIEZO1* gene located on chromosome 16 and two variant detected in the POC sample. (**D**) Log-normalized expression of *PIEZO1* gene in three stages: Embryonic E3-E16 (Embryonic day 3 to 16), organogenesis (4-6 weeks of gestation), and fetal (10-20 weeks of gestation) development.

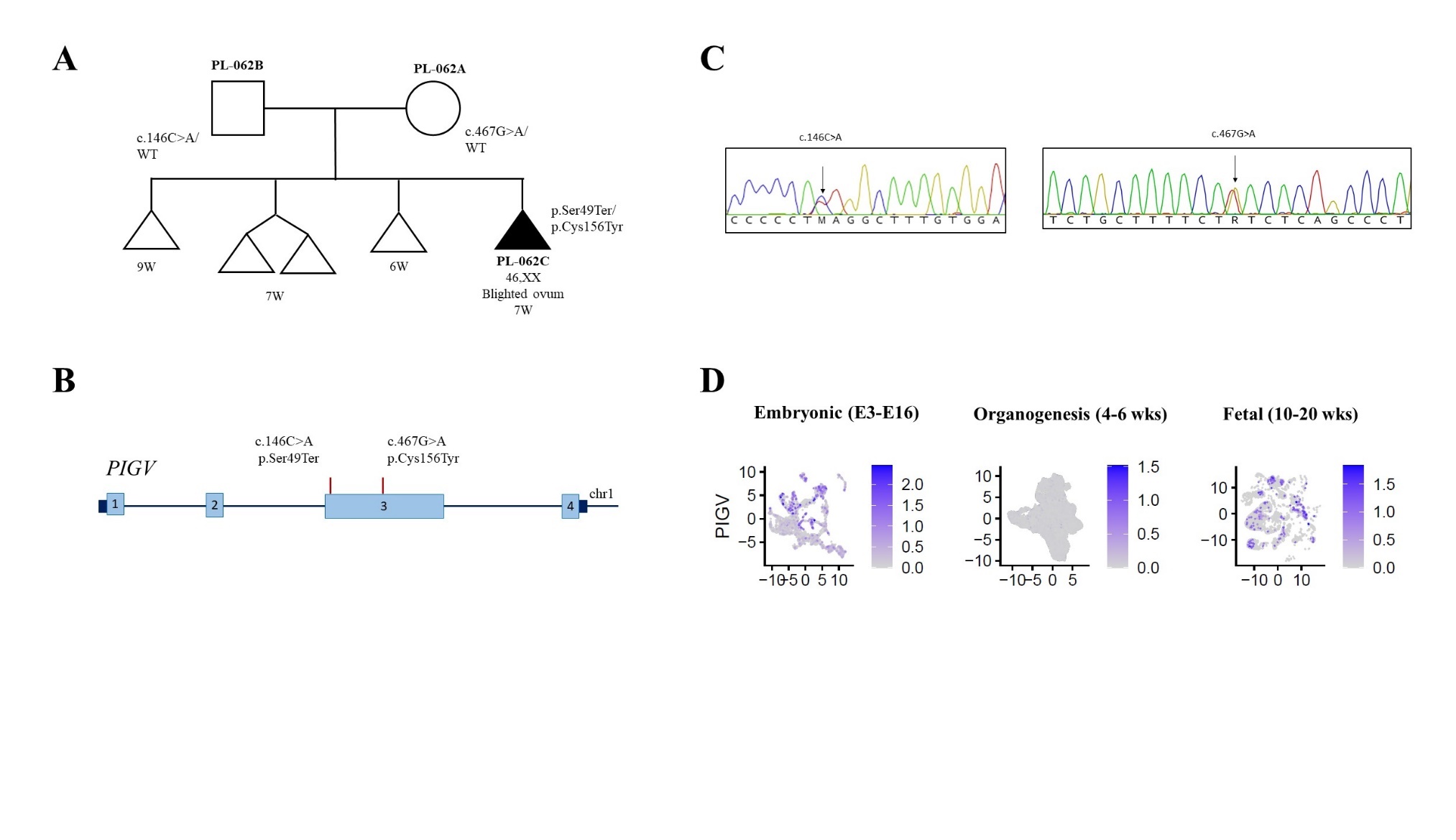

**Figure S3-18. Findings in the *PIGV* gene.**

(**A**) Pedigree of the PL-062 family. (**B**) Schematic representation of the *PIGV* gene located on chromosome 1. In pregnancy loss sample PL-062C, compound heterozygous variant including a maternally inherited missense variant in exon 3 and paternally inherited nonsense variant were found in exon 3 and (**C**) confirmed by Sanger sequencing. (**D**) Log-normalized expression of *PIGV* gene in three stages: Embryonic E3-E16 (Embryonic day 3 to 16), organogenesis (4-6 weeks of gestation), and fetal (10-20 weeks of gestation) development.

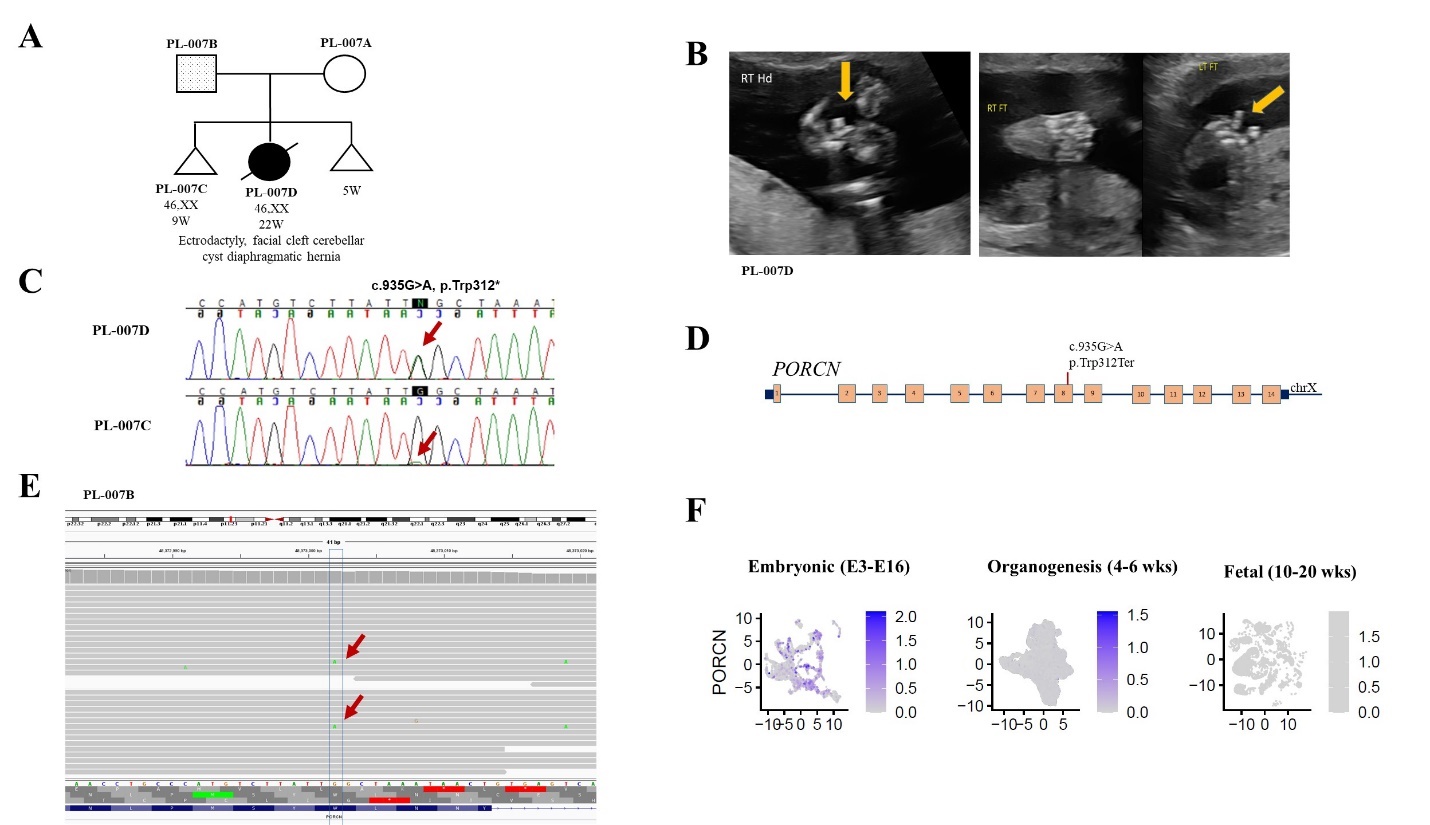

**Figure S3-19. Findings in the *PORCN* gene.**

(**A**) Pedigree of the PL-007 family. (**B**) Prenatal ultrasound in PL-007D showed multiple congenital anomalies and ectrodactyly (yellow arrow) in left (L) foot and right (R) hand. In pregnancy loss sample PL-007D, a stop gained heterozygous variant was found in exon 8 of *PORCN* gene and (**C**) confirmed by Sanger sequencing. A suspicion for the same change (red arrow) was raised after sequencing of a sample from the previous POC (PL-007C). (**D**) Schematic representation of the *PORCN* gene located on the X chromosome. (**E**) Exome sequencing with 100X coverage showed 3% paternal germline mosaicism for the same variant as found in POC samples. (**F**) Log-normalized expression of *PORCN* gene in three stages: Embryonic E3-E16 (Embryonic day 3 to 16), organogenesis (4-6 weeks of gestation), and fetal (10-20 weeks of gestation) development.

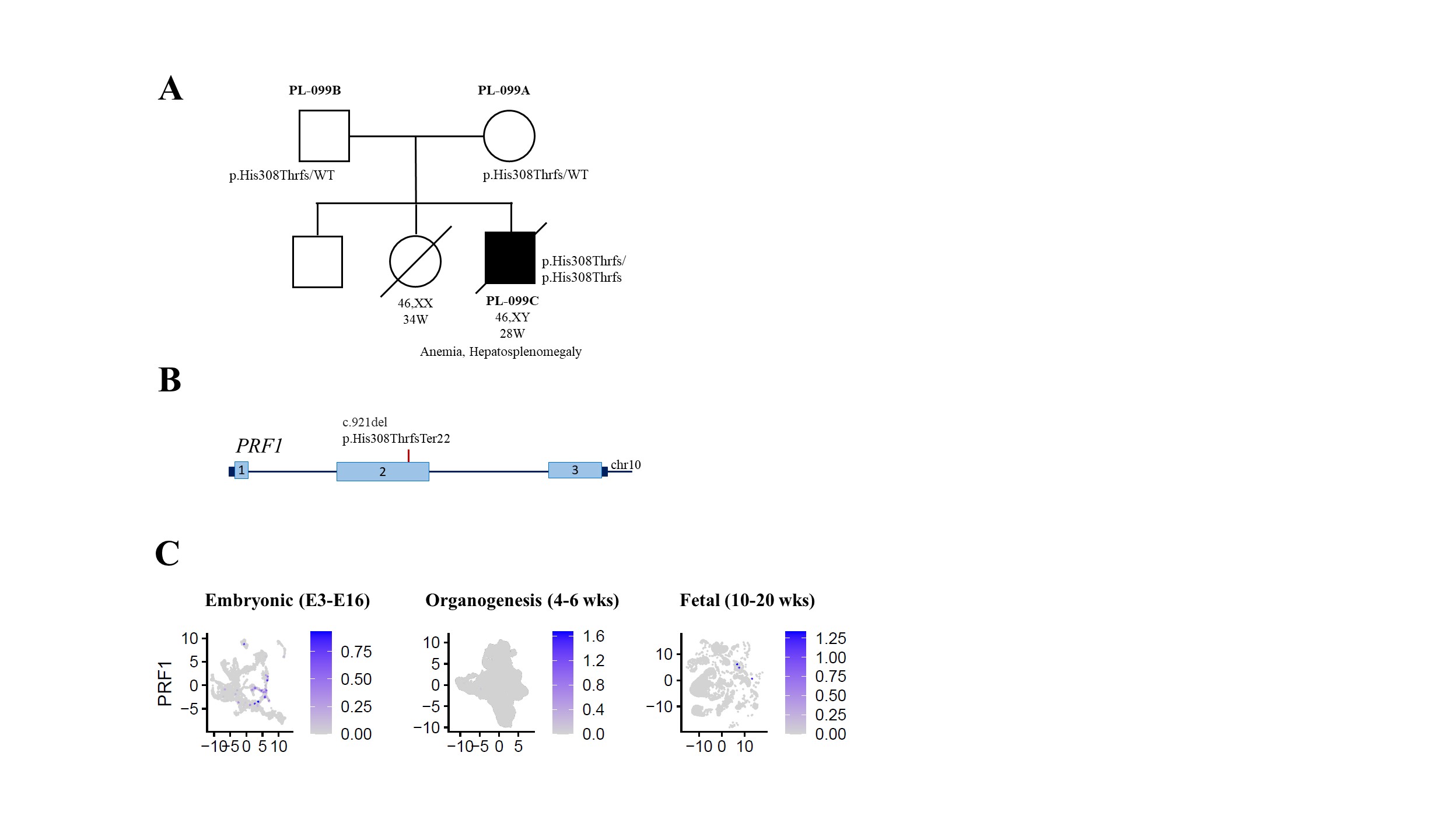

**Figure S3-20. Findings in the *PRF1* gene**.

(**A**) Pedigree of the PL-099 family. (**B**) Schematic representation of the *PRF1* gene located on chromosome 10. In pregnancy loss sample PL-099C, a frameshift homozygous variant was found in exon 2. (**C**) Log-normalized expression of *PRF1* gene in three stages: Embryonic E3-E16 (Embryonic day 3 to 16), organogenesis (4-6 weeks of gestation), and fetal (10-20 weeks of gestation) development.

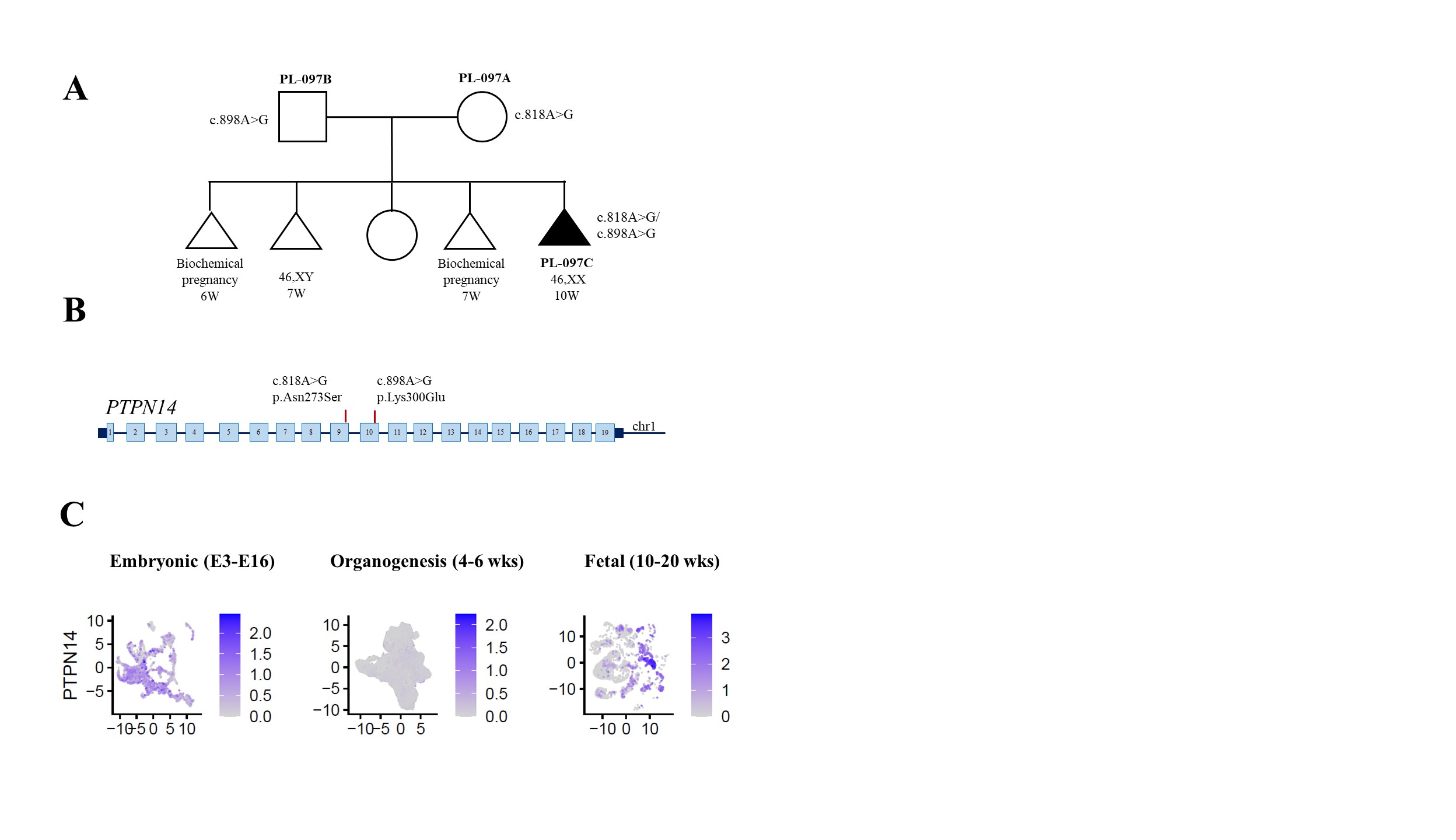

**Figure S3-21. Findings in the *PTPN14* gene.**

(**A**) Pedigree of the PL-097 family. (**B**) Schematic representation of the *PTPN14* gene located on chromosome 1. In pregnancy loss sample PL-097C, two missense variants in exon 9 and exon 10 were identified. (D) Log-normalized expression of *PTPN14* gene in three stages: Embryonic E3-E16 (Embryonic day 3 to 16), organogenesis (4-6 weeks of gestation), and fetal (10-20 weeks of gestation) development.

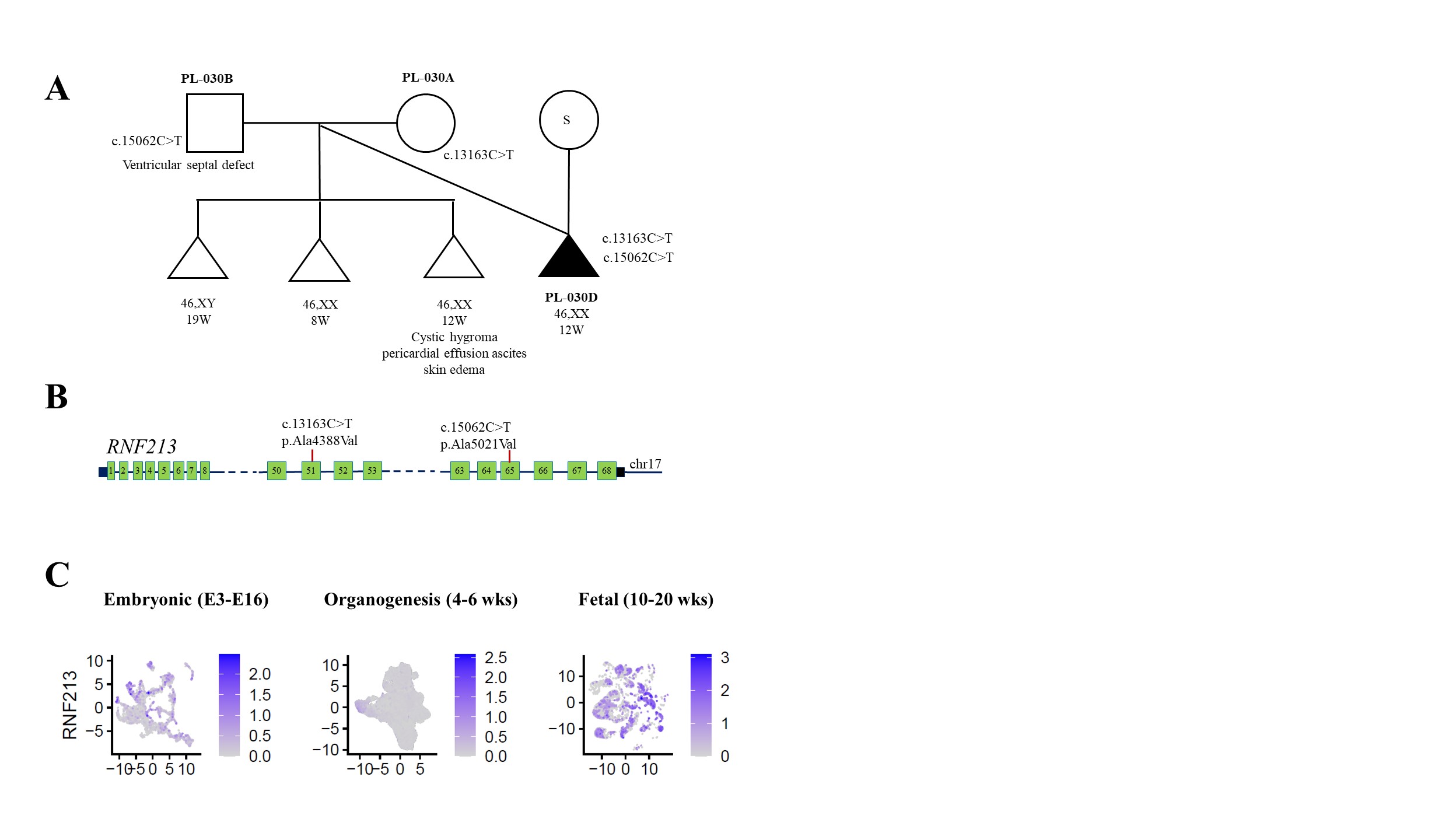

Figure **S3-22. Findings in the *RNF213* gene.**

(**A**) Pedigree of the PL-030 family. A surrogate carrier is indicated by “S” in the circle. (**B**) Schematic representation of the *RNF213* gene located on chromosome 17. In pregnancy loss sample PL-030D, each parent transmitted a variant to the POC. (**C**) Log-normalized expression of *RNF213* gene in three stages: Embryonic E3-E16 (Embryonic day 3 to 16), organogenesis (4-6 weeks of gestation), and fetal (10-20 weeks of gestation) development.

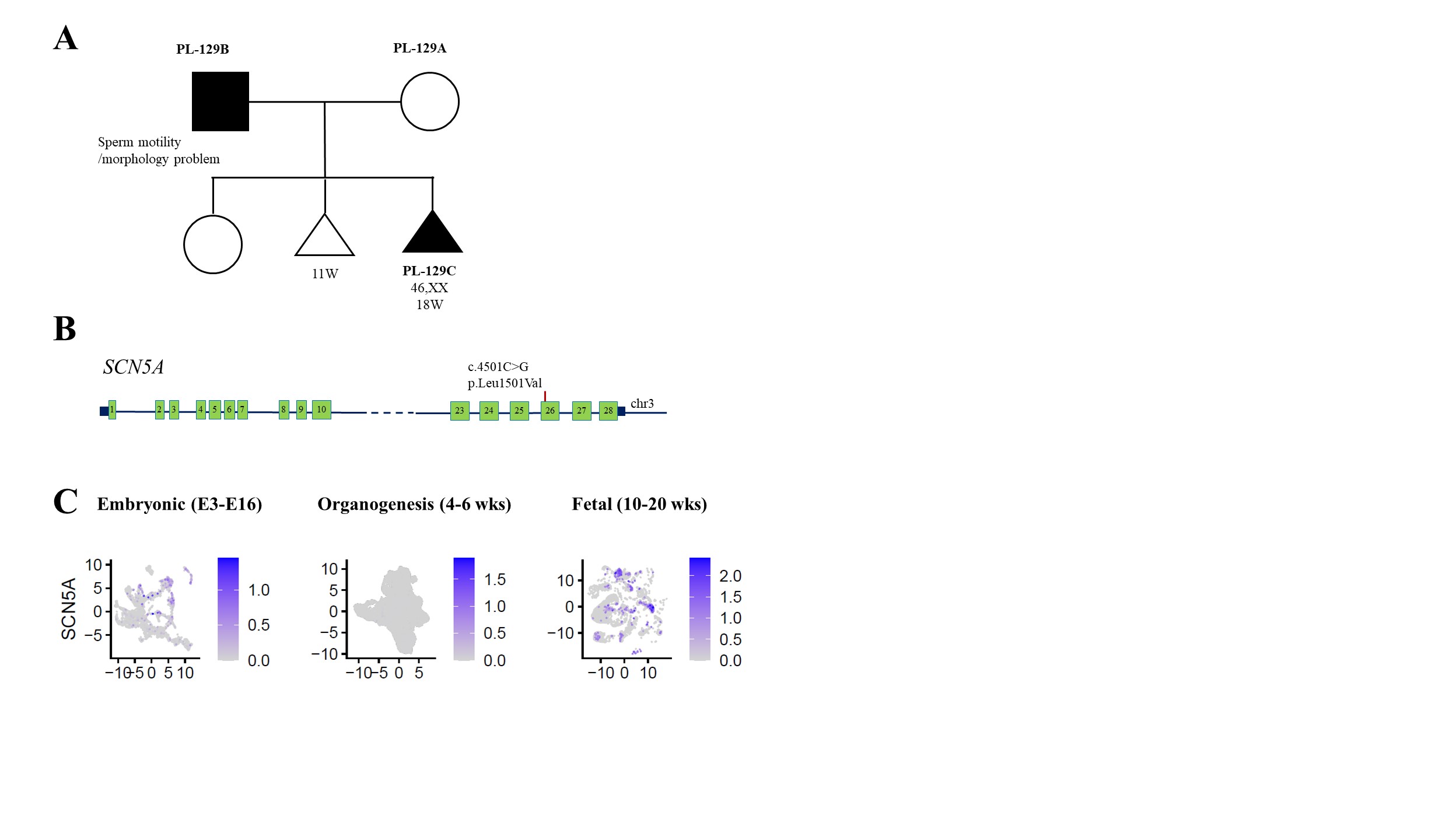

**Figure S3-23. Findings in the *SCN5A* gene.**

(**A**) Pedigree of the PL-129 family. (**B**) Schematic representation of the *SCN5A* gene located on chromosome 3. In pregnancy loss sample PL-129C, a paternally inherited missense heterozygous variant was found in exon 26 and (**C**) confirmed by Sanger sequencing. (**D**) Log-normalized expression of *SCN5A* gene in three stages: Embryonic E3-E16 (Embryonic day 3 to 16), organogenesis (4-6 weeks of gestation), and fetal (10-20 weeks of gestation) development.

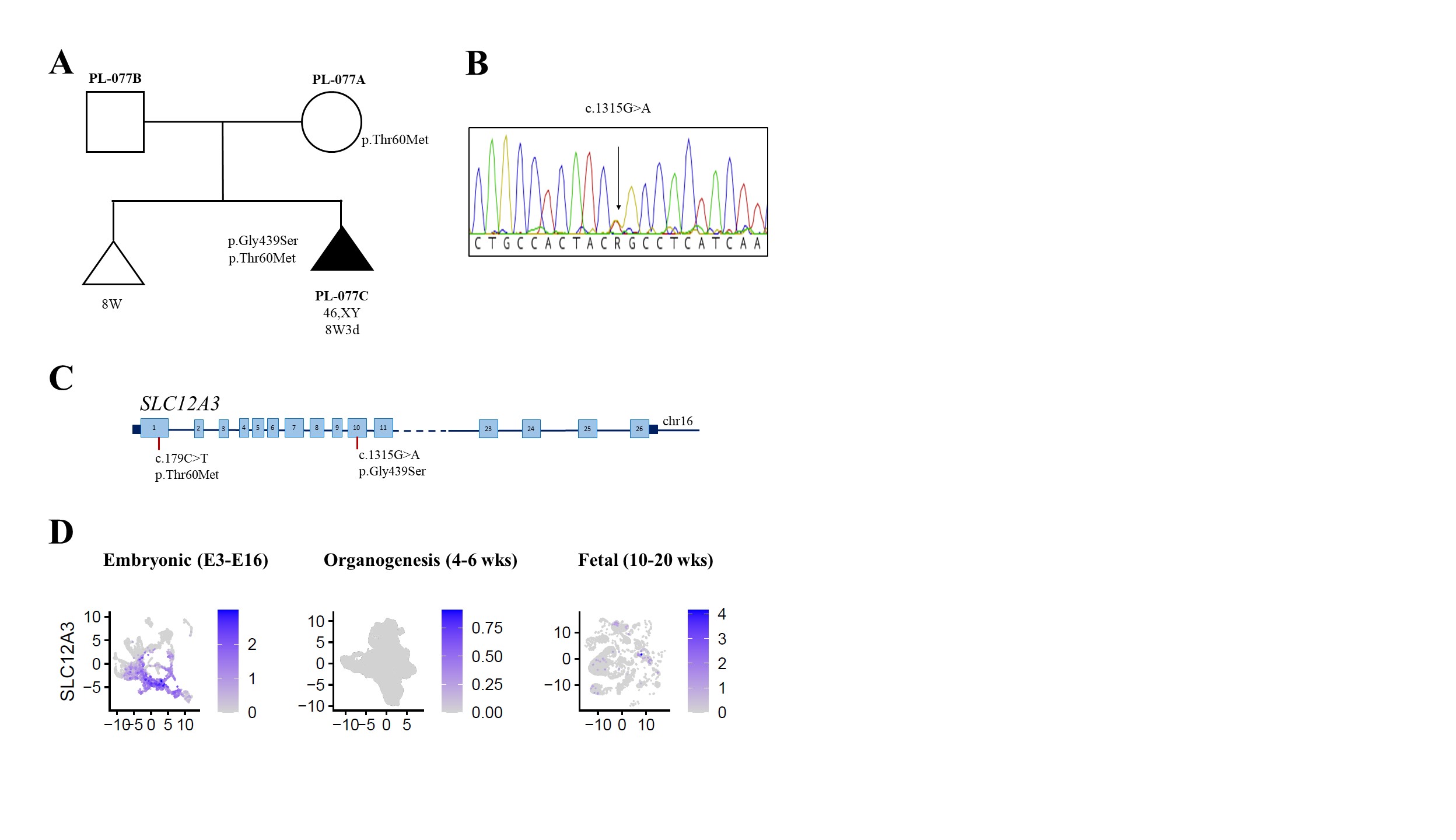

**Figure S3-24. Findings in the *SLC12A3* gene.**

(**A**) Pedigree of PL-077 family. In pregnancy loss sample PL-077C, a de novo missense heterozygous variant c.1315G>A, p.(Gly439Ser) was found in exon 10 and a maternally inherited missense heterozygous variant c.179C>T, p.(Thr60Met) was found in exon 1 of *SLC12A3*. (**B**) Chromatogram showing the de novo pathogenic variant in the POC. (**C**) Schematic representation of the *SLC12A3* gene located on chromosome 16 and variants found in the POC. (**D**) Log-normalized expression of *SLC12A3* gene in three stages: Embryonic E3-E16 (Embryonic day 3 to 16), organogenesis (4-6 weeks of gestation), and fetal (10-20 weeks of gestation) development.

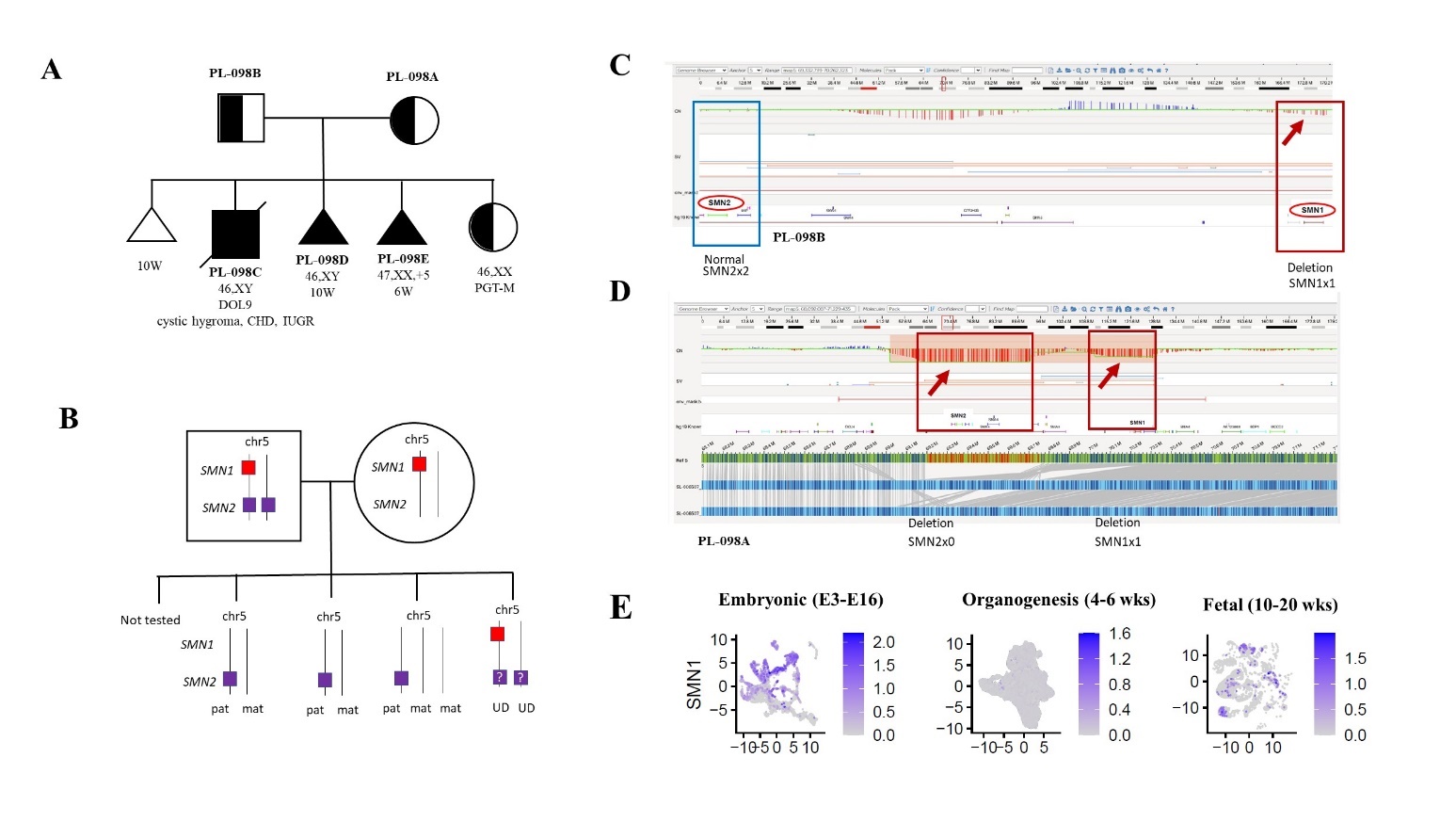

**Figure S3-25. Findings in the *SMN1* and *SMN2* gene.**

(**A**) Pedigree of the PL-098 family with 3 pregnancy losses and a neonatal death at 9 days of life (DOL). (**B**) In PL-098C sample, zero copies of the *SMN1* gene and one copy of *SMN2* was found, both located on chromosome 5. The same genotype (SMN1x0, SMN2x1) was detected in two consecutive pregnancy losses (PL-098D and PL-098E), despite trisomy 5 in PL-098E. (**C**) Optical Genome Mapping (OGM) revealed *SMN1* deletion (red arrow) in the father. **(D**) OGM of maternal sample revealed a heterozygous deletion of *SMN1* gene and a biallelic loss of the *SMN2* gene (red arrows). **(E)** Log-normalized expression of the *SMN1* gene in three stages: Embryonic E3-E16 (Embryonic day 3 to 16), organogenesis (4-6 weeks of gestation), and fetal development (10-20 weeks of gestation).

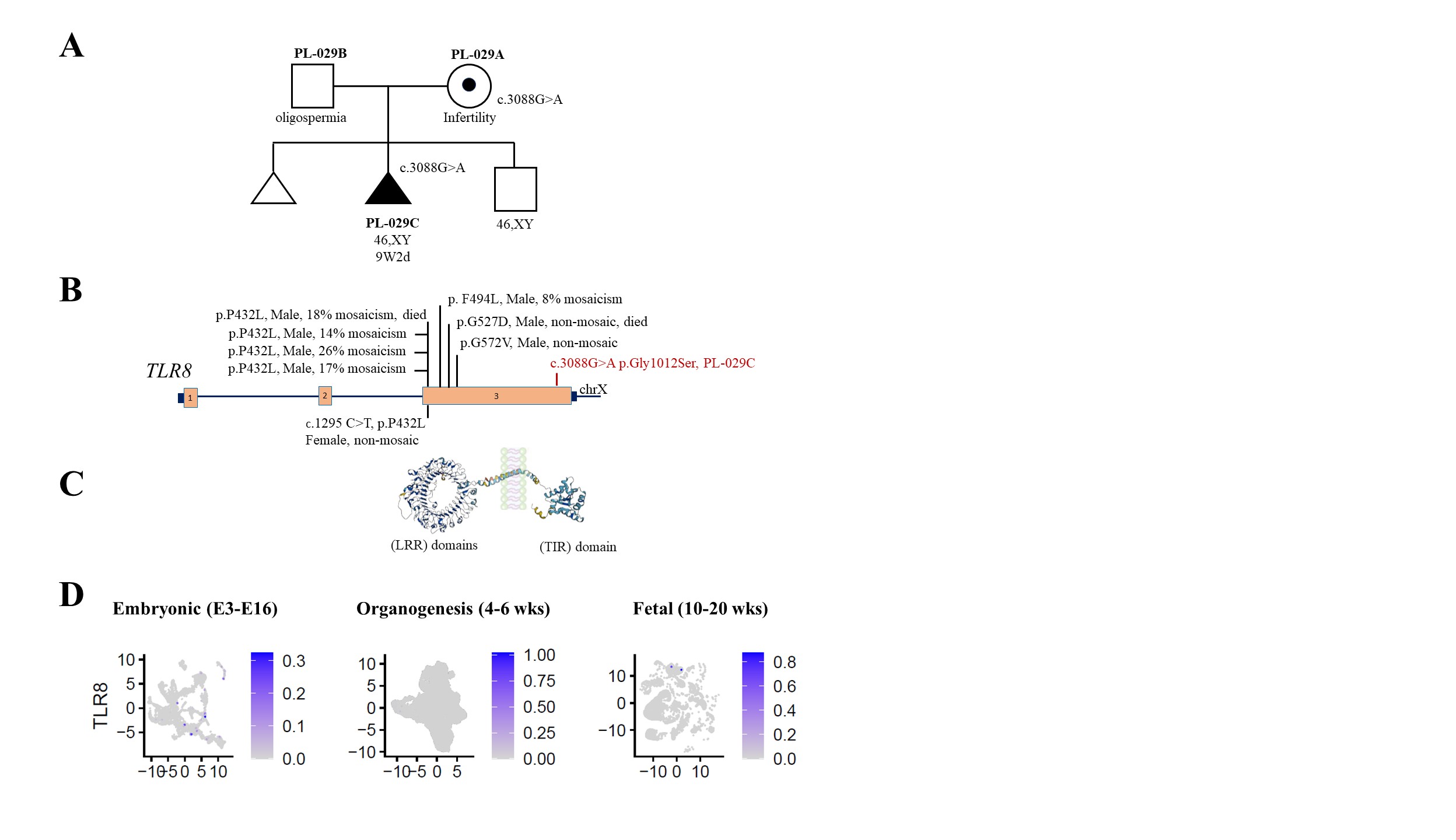

**Figure S3-26. Findings in the *TLR8* gene.**

(**A**) Pedigree of the PL-029 family. In pregnancy loss sample PL-029C, a maternally inherited missense hemizygous variant was found the *TLR8* gene. (**B**) Schematic representation of the *TLR8* gene located on chromosome X and variants detected in affected patients published previously. The *TLR8* variant in PL-029C (in red) is located within the TIR domain, responsible for a homodimer formation. (**C**) A 3D structure of a monomeric transmembrane toll-like receptor containing the leucine-rich repeats (LRR) intra-endosomal domain and a cytoplasmic (Toll/interleukin-1 receptor (TIR) domain. (**D**) Log-normalized expression of *TLR8* gene in three stages: Embryonic E3-E16 (Embryonic day 3 to 16), organogenesis (4-6 weeks of gestation), and fetal (10-20 weeks of gestation) development.

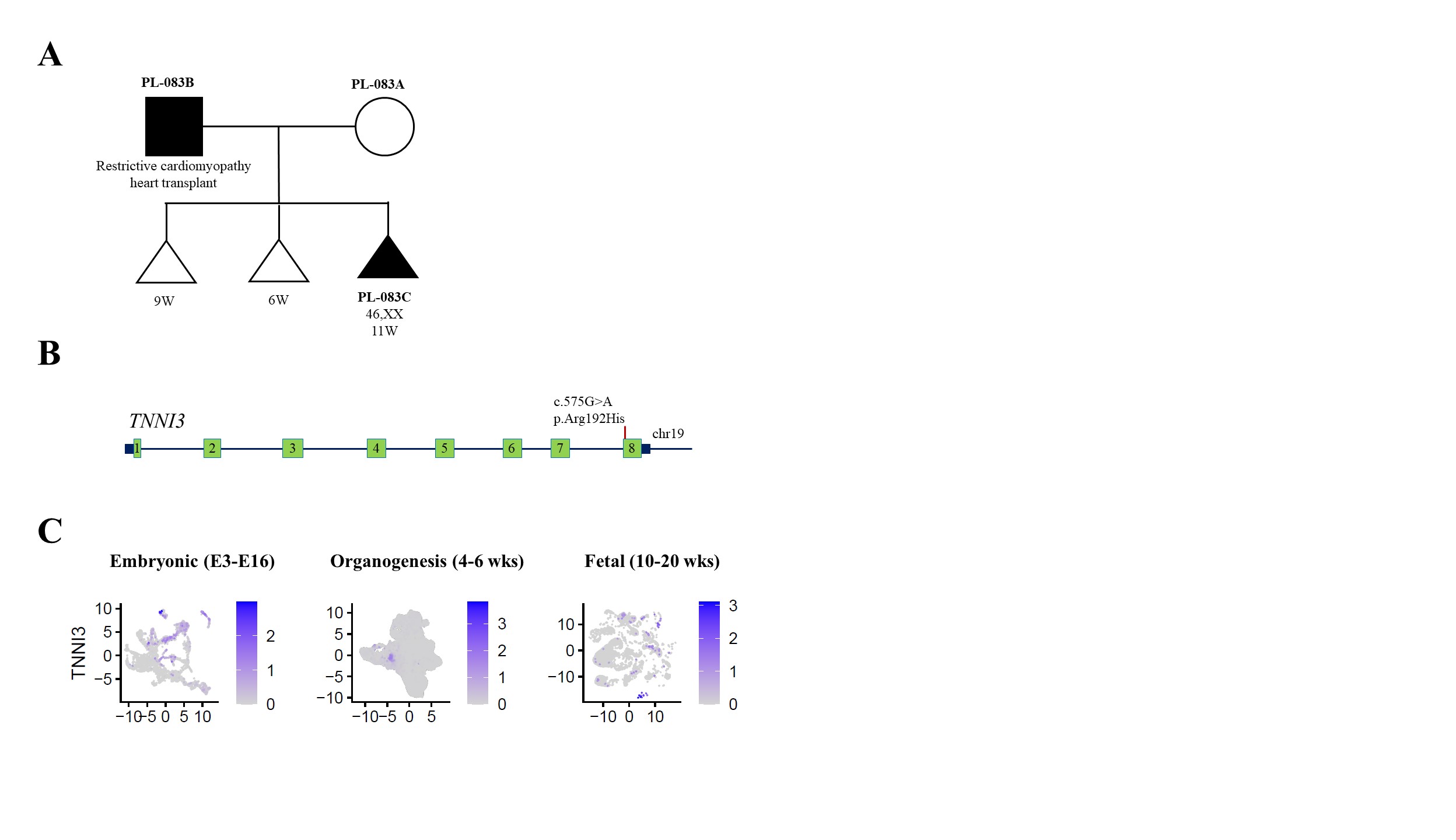

**Figure S3-27. Findings in the *TNNI3* gene.**

(**A**) Pedigree of the PL-083 family. In pregnancy loss sample PL-083C, a paternally inherited missense heterozygous variant was found and (**B**) confirmed by Sanger sequencing. (**C**) Schematic representation of the *TNNI3* gene located on chromosome 19. (**D**) Log-normalized expression of *TNNI3* gene in three stages: Embryonic E3-E16 (Embryonic day 3 to 16), organogenesis (4-6 weeks of gestation), and fetal (10-20 weeks of gestation) development.

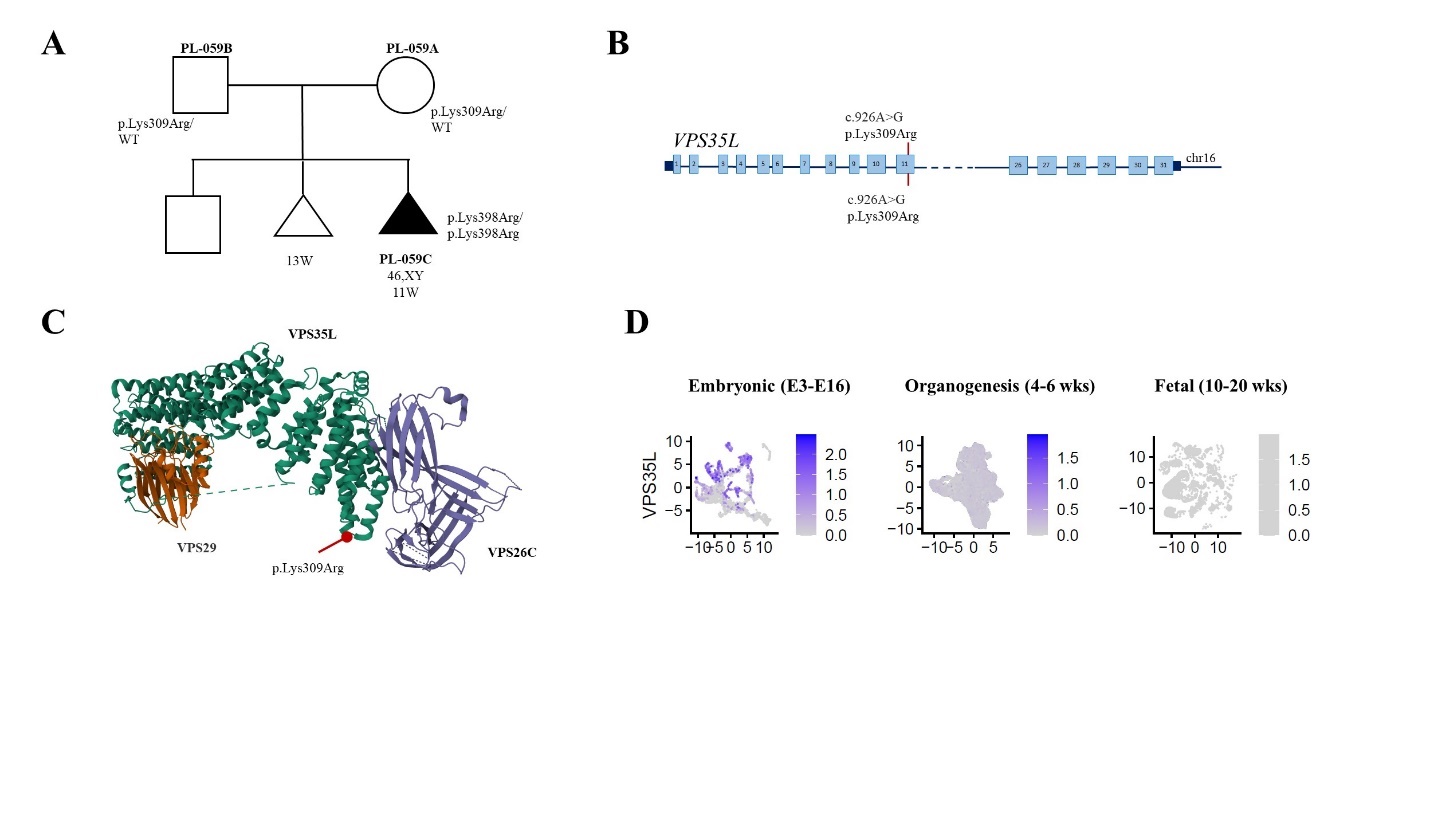

**Figure S3-28. Findings in the *VPS35L* gene.**

(**A**) Pedigree of the PL-059 family. In pregnancy loss sample PL-059C, a homozygous missense variant was found in exon 11 of *VPS35L* gene. Both parents carried a heterozygous variant. (**B**) Schematic representation of the *VPS35L* gene located on chromosome 16. (**C**) The 3D model of the VPS35L-VPS26-VPS29 trimer complex obtained from (<https://www.rcsb.org/structure/8SYN>). The substitution of a highly conserved Lysine by Arginine at 309 position (red dot) is proposed to affect interaction with VPS26 or other molecules within the Commander complex. (**D**) Log-normalized expression of *VPS35L* gene in three stages: Embryonic E3-E16 (Embryonic day 3 to 16), organogenesis (4-6 weeks of gestation), and fetal (10-20 weeks of gestation) development.

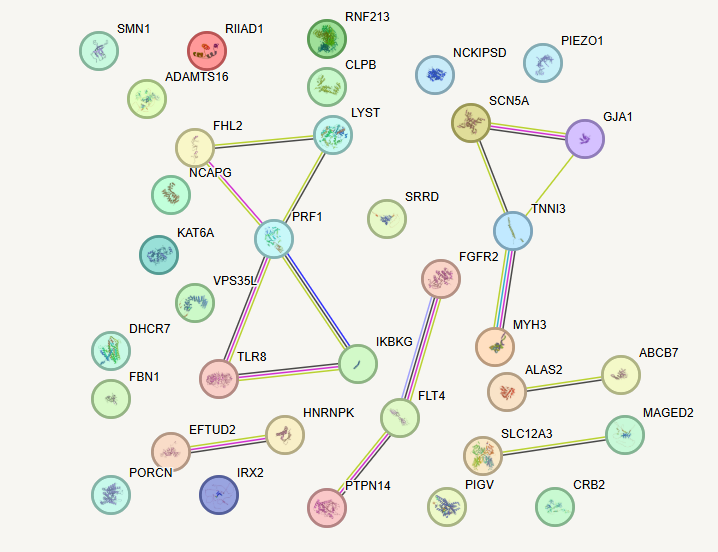

**Figure S4.** Protein-protein interaction between the genes found in a cohort of pregnancy loss samples. Data from curated interaction databases, protein interaction information, co-expression, and gene ontology were utilized to form the STRING (https://string-db.org/) gene functional network for 28 detected genes and 7 candidate genes with no disease association.

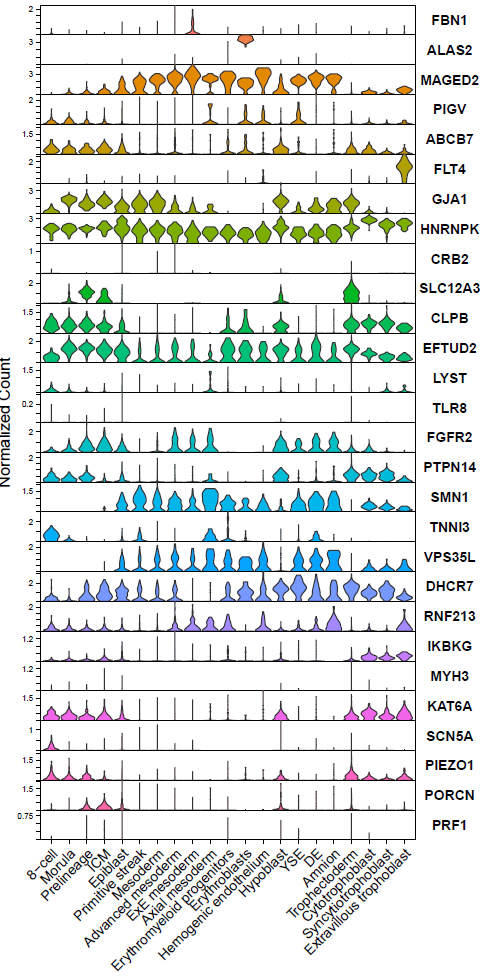

**Figure S5.** Transcriptomic profiles of identified genes from preimplantation development to gastrulation in human embryos. Violin plots show distribution of log normalized gene expression in single cells for each cell type. Data shown were integrated from published datasets.^5-8^ ICM, inner cell mass. ExE, extraembryonic. YSE, yolk sac endoderm. DE, definitive endoderm.

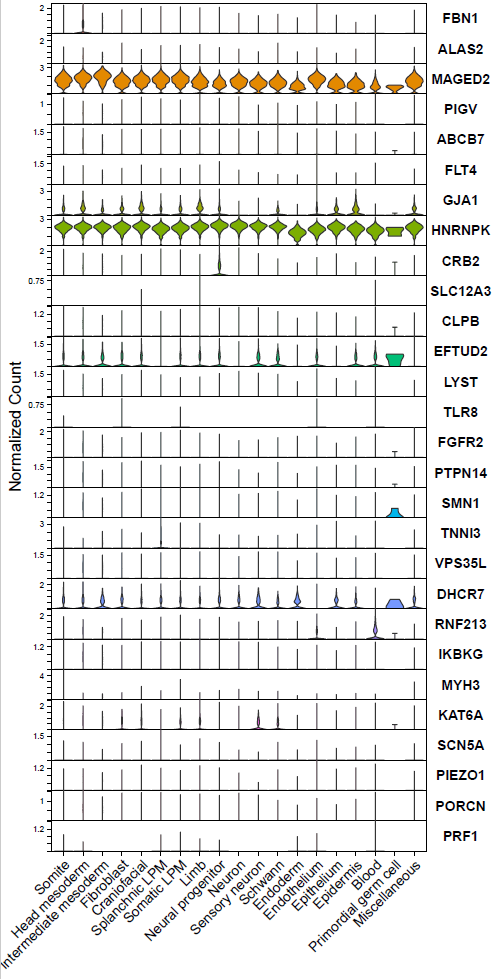

**Figure S6**. Transcriptomic profiles of identified genes during organogenesis in human embryos from about 4 to 6 weeks gestation. Violin plots show distribution of log normalized gene expression in single cells for each cell type. Data shown were derived from a published dataset.^9^ LPM, lateral plate mesoderm.

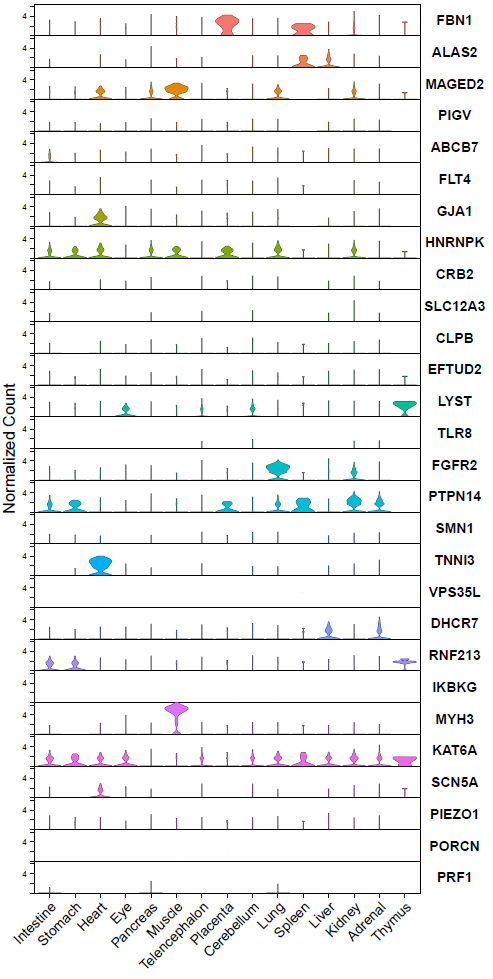

**Figure S7.** Transcriptomic profiles of identified genes in tissues from human fetuses from 10 to 20 weeks gestation. Violin plots show distribution of log normalized gene expression in single cells for each tissue. Data shown were derived from a published dataset.^10^ No counts shown for genes *VPS35L*, *IKBKG*, and *PORCN* as these genes were not included in the published dataset.
