## Supplementary material for "Genetic Variants in Recurrent Euploid Pregnancy Loss": Response to Medrxiv reviewers comments

September 30, 2025

Dear medRxiv Screening Team,

Thank you for your careful review of our submission, “Genetic Variants in Recurrent Euploid Pregnancy Loss” (MS ID: MEDRXIV/2025/335660). We appreciate the guidance and have revised the manuscript and supplement. Below we summarize the changes:

**Medrxiv Comment 1**
Precise ages should be removed or replaced with e.g. "in their 60’s" or an age range (5-year range minimum or an age range in quartiles based on the age range of your study). Age ranges cannot overlap. Example of non-overlapping age ranges: 21-25, 26-30, 31-35, 36-40, etc. Please address the following and any other instances in your manuscript:

- Gestational and parental ages in Table 1, page 29
- Gestational ages in the Figures (S3-1 to S3-28 (in file "SUPPLEMENTARY Appendix-TRIOS.docx"

**Response to comment 1**
*We have revised* ***parental ages*** *into non-overlapping 5-year bins (21–25, 26–30, 31–35, 36–40, 41–45, ≥46) in Table S1 and removed them from Table 1.*
*For gestational age (GA), we retained exact values because GA at pregnancy loss is a critical biological parameter in recurrent pregnancy loss research. Without GA, recurrence timing and outcomes cannot be interpreted, and the scientific validity of the study would be compromised. Similar genetic studies (Byrne et al. 2023; Workalemahu et al. 2023; Stanley et al. 2020; Wang et al. 2023; Xiang et al. 2023) have also reported GA in full, as it is integral to interpreting reproductive outcomes. We emphasize that this analysis involves a large cohort of 118 families, not individual case reports, which substantially reduces the risk of re-identification. All samples were fully de-identified, participants provided informed consent, and GA values are not presented alongside other identifiers (e.g., location, dates, years of loss, occupation, ethnicity, exact parental ages).*

**Medrxiv Comment 2**
Sample/patient IDs (e.g., Family and Individual IDs "PL-130 and PL-130C") should be removed if known to anyone (e.g., hospital staff or patients themselves) outside the research group. If removal of sample/patient IDs is not possible, they should be replaced with identifiers that cannot reveal the identity of the study subjects. If you wish to keep the sample/patient IDs and they were not known to anyone outside the research group, please confirm that the latter is the case.

**Response to comment 2:**
*The family IDs starting with “PL” are internal and de-identified study codes that are not known to participants, or clinicians, and staff outside the research team. They do not contain information about recruitment sites, dates, or personal identifiers. As such, they do not pose a risk of participant identification.*

**Medrxiv Comment 3**
Pedigrees (e.g., Figures S3-1 to S3-28 in file "SUPPLEMENTARY Appendix-TRIOS.docx") that can potentially make patients identifiable should be removed. You may edit them per the recommendations below or advise readers to contact the corresponding author to request access to those data.
- Full pedigrees can be substituted with partial pedigrees (i.e. not all family members are included or sex of relatives that are not relevant for the study is not shown)
- Complex identifying pedigrees can also be split into simple, disjoint two-generation parent/child pedigrees.
- Please note that we require that there be less than three indirect identifiers remaining in your manuscript and supplemental information (if present) for the study to be approved for posting on medRxiv. Examples of indirect identifiers are:
- Age range
- Ethnicity
- Religion
- Details about family and family history
- Potentially identifying medical conditions
- Occupation
- Professional affiliations
- Geographical area

**Response to comment 3**
*In our study, all pedigrees include parents and their pregnancies. We intentionally excluded unrelated or irrelevant relatives from other generations to minimize the information. Gestational ages are included in the pedigrees because they are essential for interpreting recurrence patterns and timing of losses, which is the primary focus of our study. Without this information, the scientific validity of the analysis would be compromised. Importantly, pedigrees contain no additional identifiers listed in your guidance (e.g. location, dates, ethnicity, religion, occupation, professional affiliations), and the data are presented across a cohort of 118 families, not as isolated case reports, which minimizes any risk of participant re-identification.*

We uploaded the files in Medrxiv portal and our core findings remain as originally reported. Please let us know if any further adjustments are needed. We appreciate your time and look forward to posting the revised preprint.

Best regards,

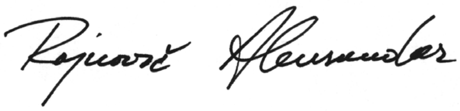

Aleksandar Rajkovic, MD, PhD
Stuart Lindsay Distinguished Professor in Experimental Pathology
Department of Pathology
University of California, San Francisco
